# Longitudinal analyses of depression and anxiety highlight greater prevalence during COVID-19 lockdowns in the Dutch general population and a continuing increase in suicidal ideation in young adults

**DOI:** 10.1101/2022.05.02.22273554

**Authors:** Anil P. S. Ori, Martijn Wieling, Lifelines Corona Research Initiative, Hanna M. van Loo

## Abstract

**Objective:** The pandemic of the coronavirus disease 2019 (COVID-19) has led to an increased burden on mental health. This study therefore investigated the development of major depressive disorder (MDD), generalized anxiety disorder (GAD), and suicidal ideation in the Netherlands during the first fifteen months of the pandemic and three nation-wide lockdowns.

**Methods:** Participants of the Lifelines Cohort Study –a Dutch population-based sample-reported current symptoms of MDD and GAD, including suicidal ideation, according to DSM-IV criteria using a digital structured questionnaire. Between March 2020 and June 2021, 36,106 participants (aged 18-96) filled out a total of 629,811 questionnaires across 23 time points. Trajectories over time were estimated using generalized additive models and analyzed in relation to age, sex, and lifetime history of MDD/GAD to identify groups at risk.

**Results:** We found non-linear trajectories for MDD and GAD with a higher number of symptoms and prevalence rates during periods of lockdown. The point prevalence of MDD and GAD peaked during the third hard lockdown at 2.88% (95% CI: 2.71%–3.06%) and 2.92% (95% CI: 2.76%-3.08%), respectively, in March 2021. Women, younger adults, and participants with a history of MDD/GAD reported significantly more symptoms. For suicidal ideation, we found a linear increase over time in younger participants which continued even after the lockdowns ended. For example, 4.63% (95% CI: 3.09%-6.96%) of 20-year-old participants reported suicidal ideation at our last measured time point in June 2021, which represents a 4.14x increase since the start of the pandemic.

**Conclusions:** Our study showed greater prevalence of MDD and GAD during COVID-19 lockdowns suggesting that the pandemic and government enacted restrictions impacted mental health in the population. We furthermore found a continuing increase in suicidal ideation in young adults. This warrants for alertness in clinical practice and prioritization of mental health in public health policy.

## Introduction

The ongoing pandemic of the coronavirus disease 2019 (COVID-19) has a major impact on societies and led to increases in major depressive disorder (MDD), generalized anxiety disorder (GAD), and suicidality across the world[1–3]. These conditions are severe and disabling and represent major contributors to the global burden of disease and mortality[4]. As most studies on depression, anxiety, and suicidality so far were conducted over the first months of the pandemic and were cross-sectional in their design, they do not provide temporal insights on mental health consequences over time nor on the impact of later COVID-19 lockdowns or other governmental restrictions. Continued longitudinal monitoring of changes in mental health in the population is therefore important as it can help identify groups who are at risk as well as inform government actions to reduce spread of the virus while also prioritizing the mental health of residents.

Here, we investigate the development of (symptoms of) MDD, GAD, and suicidal ideation during the COVID-19 pandemic in Lifelines, a large population-based cohort in the North of the Netherlands. Between April 2020 and July 2021, MDD, GAD and suicidal ideation were repeatedly assessed in >76,000 participants, which represents the largest longitudinal cohort on pandemic-related impact on mental health in the population[5]. Using detailed self-reported longitudinal data, we estimated the prevalence of these outcomes across the first fifteen months of the pandemic and three nation-wide lockdown periods. We furthermore investigated differences in risk across age, sex, and lifetime history of MDD/GAD.

## Methods

Full details on cohort information, digital questionnaires, and our analytical strategy can be found in the supplemental materials.

### The Lifelines COVID-19 Cohort

Lifelines is a multi-disciplinary prospective population-based cohort study examining in a unique three-generation design the health and health-related behaviours of 167,729 persons living in the North of the Netherlands. It employs a broad range of investigative procedures in assessing the biomedical, socio-demographic, behavioural, physical and psychological factors which contribute to the health and disease of the general population, with a special focus on multi-morbidity and complex genetics.

In March 2020, the Lifelines Corona Research Initiative was initiated to monitor the physical and mental health of residents in the three Northern provinces of the Netherlands during the COVID-19 pandemic through detailed digital questionnaires[5]. The Lifelines COVID-19 cohort is embedded in Lifelines, a large multi-generational prospective population-based study and biobank with extensive information collected on health, lifestyle and sociodemographic data[6, 7]. All participants provided written informed consent. The Lifelines Cohort Study was approved by the Medical Ethics Committee of the University Medical Center Groningen, The Netherlands.

### COVID-19 questionnaires and sample selection

In March 2020, the first digital questionnaire was sent to all 140,145 adult Lifelines participants with an e-mail address on file[5]. Follow-up questionnaires were initially sent on a weekly (questionnaires 1 to 6, Q1-Q6) and later on a biweekly and monthly basis (Q7-Q23). Up to July 2021, 23 questionnaires have been sent out with 76,376 study participants filling in at least one questionnaire (Figure S1). To minimize the impact of participation bias, i.e. participants with MDD and GAD were less likely to participate in the next questionnaire (see supplementary methods), we selected participants aged 18 years and older who filled out at least one questionnaire in Q1-Q3 and at least one questionnaire in Q21-Q23 to conduct our primary statistical analyses (N=36,106).

### Outcome measures

Current symptoms of MDD and GAD reflecting the DSM-IV criteria[8] were assessed using a digital self-report version of the Mini-International Neuropsychiatric Interview (MINI)[9], which has also been implemented in earlier assessments in Lifelines[10]. All items had a binary response option (yes/no)(Table S1, Figure S2). The first questionnaires (Q1-Q6) assessed current symptoms during the past seven days (as Q1-Q6 were sent out weekly), while the later questionnaires (Q7-Q23) assessed during the past 14 days (as Q7-Q23 were sent out biweekly or monthly). We extracted five outcomes for our analyses: sum scores for depressive (range 0-9) and anxiety symptoms (range 0-7), MDD, GAD according to the DSM-IV-TR criteria, and suicidal ideation. Suicidal ideation was assessed as present if a participant reported to have considered hurting themselves, wished they were dead, or had suicidal thoughts in the past seven/fourteen days.

### Predictors

We used four predictors in our analyses: time, age, biological sex, and lifetime history of MDD/GAD (see supplementary methods). Lifetime history of MDD and GAD were determined using an online assessment that is based on the Composite International Diagnostic Interview[11].

### Missing data and imputation

To handle missing data, we performed a single dataset imputation using a chained equation regression framework implemented in R-package mice_v3.13[12]. We imputed missing values if a participant filled out at least part of that questionnaire. Missingness within filled out questionnaires was overall limited (see supplemental methods). The missing data was imputed using information from other time points within the Lifelines COVID-19 study and from previous assessment waves in Lifelines.

### Statistical analyses

Generalized additive models (GAMs) were used to assess the population prevalence of MDD, GAD, and suicidality over time and their association with age, sex, and lifetime history of MDD/GAD. GAMs are regression models that can identify nonlinear patterns in longitudinal data[13, 14]. We modeled the prevalence of each of the five MDD/GAD outcomes as a (potentially) non-linear function of time and tested if there were significant interaction effects of time with age, sex, and lifetime history of MDD/GAD. Each outcome and predictor were fitted using a separate model. All analyses were performed in R_v4.0.3 using the packages mgcv_1.8.33[14] and itsadug_2.4[15]. Multiple testing correction was implemented by Bonferroni correction (alpha=0.0025).

### Sensitivity analyses

We conducted three sensitivity analyses (Figure S3). First, we implemented all GAMs without random effects in the full cohort (N=76,376) and compared the output with that obtained from our analyses on the main sample of 36,106 subjects. This allowed us to assess the impact of participant dropout on our findings. Second, because individuals were repeatedly assessed over time, we used a random intercept and linear random slope to account for the nested structure of the data within individuals and families. As including random effects for the full cohort was not possible due to computational constraints (see supplementary methods), we conducted the analysis on a subset of 5,000 participants (randomly drawn from the 36,106 subjects). While the GAMs with inclusion of random effects estimated lower prevalence (as only fixed effects were returned and the random effects were set to zero), it did allow us to evaluate how individual- and family-specific variation impact the observed effect of predictors and the trajectories over time in a random subset of our sample (see supplementary information). Third, as our main sample of 5,000 subjects included a low number of cases for rare phenotypes such as suicidal ideation, particularly for younger ages, we also performed a third sensitivity analysis in the youngest 5,000 participants using GAMs with random effects. This analysis included all participants from 18 to 45 years old with at least one assessment in Q1-Q3 and in Q21-Q23.

## Results

### Sample description

Our selected sample consisted of 36,106 study participants who completed a total of 629,811 questionnaires with at least 1 questionnaire in Q1-Q3 and at least one questionnaire in Q21-Q23 (Table 1 and Table S2). Participants had an average age of 57.4 years (SD= 11.9) and filled out a median number of 20 questionnaires. Women (61.9%) participated more often than men. An average of 1.9% and 2.3% of participants met the DSM-IV criteria for current MDD and GAD, respectively, during at least one assessment during the pandemic.

**Table 1.** Demographics and characteristics of the Lifelines COVID-19 study. Shown are the number of participants and their characteristics of our main analysis sample (36,106 participants). Number of MDD/GAD and suicidal ideation cases are presented as the number of participants who met the DSM-IV criteria for at least one questionnaire. The MDD/GAD prevalence and average symptom scores are presented by the mean and spread of their per-questionnaire average based on the imputed data. Demographics and characteristics of the full cohort and subsamples can be found in Table S2.

|  | **Main analysis sample** |
| --- | --- |
| Number of participants | 36,106 |
| Age | 57.4 (SD=11.9) |
| 18-30 years | 875 (2.4%) |
| 31-67 years | 27,394 (75.9%) |
| >68 years | 7,837 (21.7%) |
| Female (%) | 22,339 (61.9%) |
| Total questionnaires | 629,811 |
| Median questionnaire/person (IQR 25%-75%) | 20 (15-22) |
| Lifetime MDD (%) | 7,917 (21.9%) |
| Lifetime GAD (%) | 2,997 (8.3%) |
| Number of MDD cases | 3,675 |
| Average MDD prevalence | 1.9% |
| Average MDD symptom score | 0.50 (SD=1.14) |
| Number of GAD cases | 4,625 |
| Average GAD prevalence | 2.3% |
| Average GAD symptom score | 0.63 (SD=1.29) |
| Number of suicidal ideation cases | 2,007 |
| Average suicidal ideation prevalence | 0.71% |

From these participants, a subsample of 5,000 subjects was randomly drawn to perform sensitivity analyses to assess how individual- and family-specific variation impacts our analyses. The subsample was similar in terms of median number of questionnaires filled out, sex, age distribution, and internalizing disorder distribution to the original sample of 36,106 Lifelines participants (Table S2). Table S2 also shows the characteristics of the full sample and the subsample of the 5,000 youngest study participants that we used for sensitivity analyses.

### The COVID-19 pandemic and subsequent lockdowns in the Netherlands

During different phases of the pandemic, the Dutch government enacted a total of three nationwide lockdowns, each defined by specific measures and characteristics (Figure 1). Using data collected between March 2020 and June 2021, we next estimated the longitudinal trajectories of (symptoms of) MDD/GAD and suicidal ideation across the three lockdown periods.

**Figure 1.**
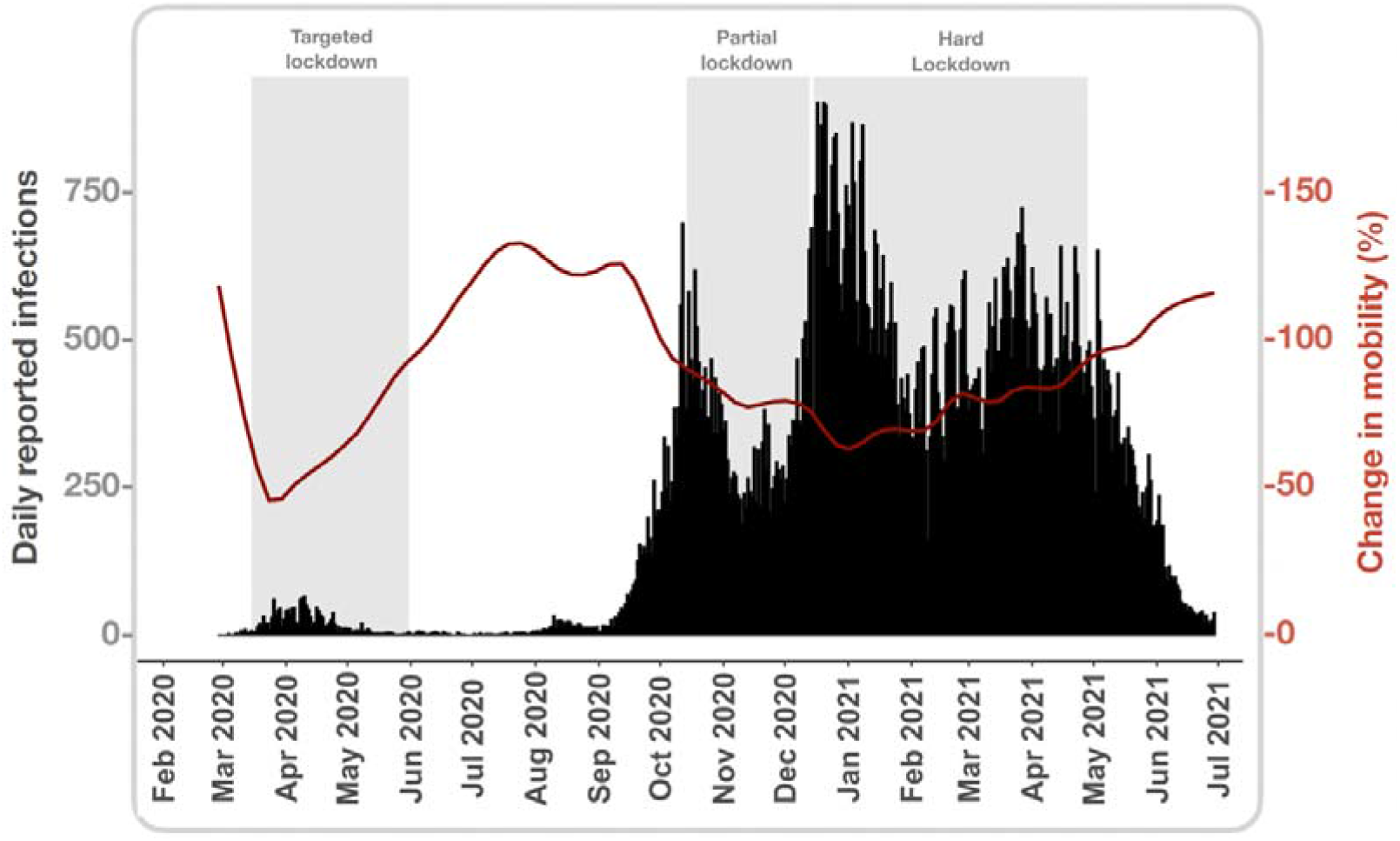
Timeline of the COVID-19 pandemic and subsequent government measures in the North of the Netherlands. Shown are the total number of daily reported infections (black bar graphs) and average change in mobility (red line) in the three Northern provinces in the Netherland (Groningen, Friesland, Drenthe) across three nation-wide lockdowns over time. The number of daily infections were downloaded from the website of the National Institute for Public Health and the Environment (RIVM) specifically for these three provinces. The change in mobility was downloaded from Apple Mobility Trends Reports also for these three provinces. The change in mobility is expressed as the percentage change compared to a baseline on January 13th, 2020. The three enacted nation-wide lockdowns are visualized by the grey vertical rectangles. The first known case of a SARS-CoV-2 infection was reported in the Netherlands on February 27 and the first reported death on March 06, 2020. The first lockdown, from March 12 - May 31, 2020, to reduce spreading of the virus was defined by targeted measures to restrict social interaction such as closing of public spaces, bars and restaurants, and work from home recommendations. The targeted lockdown reduced mobility in society by 80% but was not a hard lockdown[16]. After the subsequent summer, a new partial lockdown was announced that started on October 14 and lasted until December 14, 2020. During this period of partial lockdown, bars and restaurants were closed and limitations on social gatherings and house visits were recommended. The partial lockdown transitioned into a hard lockdown that lasted for four months which included mandatory closure of all non-essential stores and public space, closure of schools, and an evening curfew, among other measures. The national vaccination program started on January 6, 2021. Easing of restrictions of the hard lockdown were introduced on April 21 and set in motion starting April 28, 2021.

### Longitudinal trajectories of depression, anxiety, and suicidal ideation

#### Depressive symptoms

We found a significant non-linear trajectory for depressive symptoms over time (Figure 2, Table S3). Depressive symptoms were high at the start of the pandemic and declined as the first targeted lockdown progressed. Participants reported a lower number of symptoms during mid-summer, which then increased again in August 2020. Symptoms plateaued during the second partial lockdown in November and increased again during the hard lockdown after December 2020. During this third lockdown, reported symptoms reached their peak mid-March 2021 and declined again as the lockdown ended. Comparing the end of April 2021, when restrictions of the hard third lockdown started to be lifted, with the end of April 2020, participants reported more depressive symptoms a year later (0.54 compared to 0.46). The non-linear symptom trajectories were similar in sensitivity analyses in the full cohort as well as after taking into account individual- and family-specific effects (Figure S4 and Table S7, S11).

**Figure 2.**
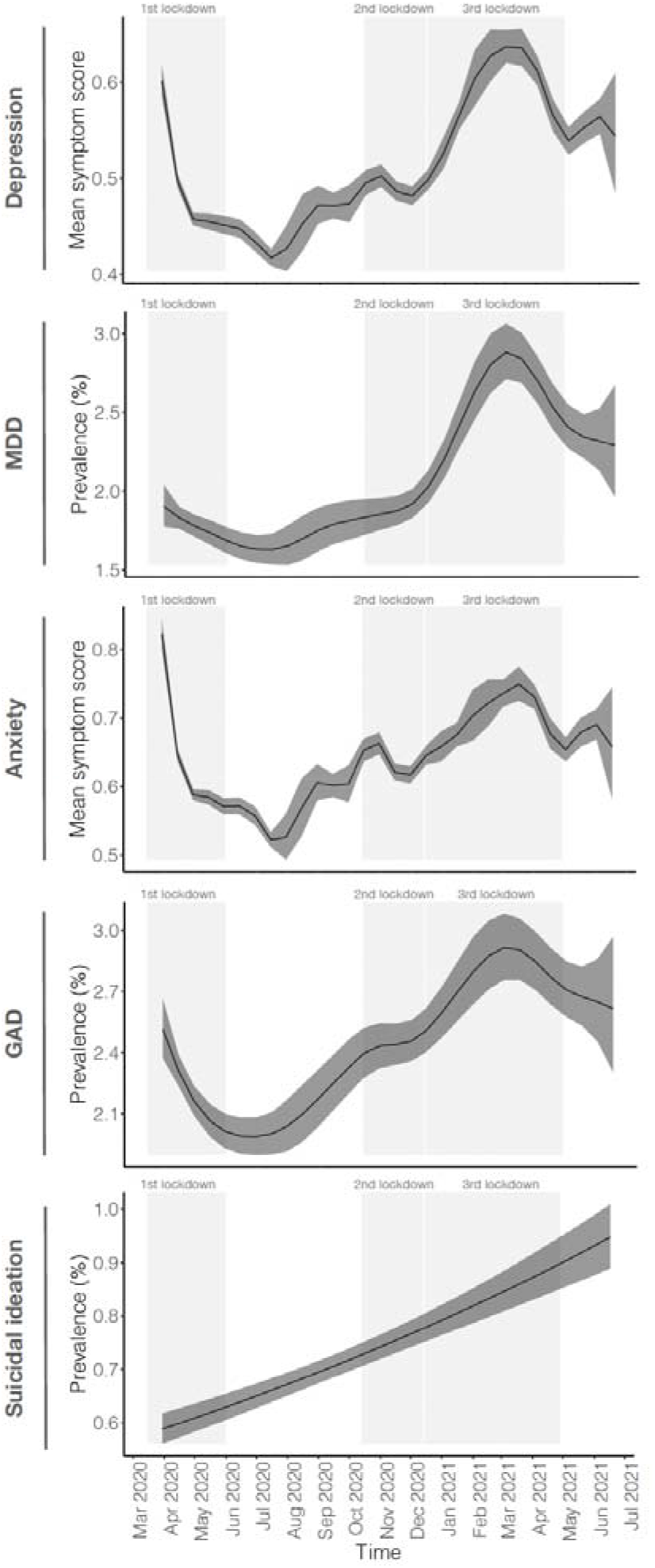
The longitudinal trajectory of MDD and GAD outcomes and suicidal ideation during the COVID-19 pandemic. Shown are the scores of reported depressive and anxiety symptoms and point prevalence of MDD, GAD, and suicidal ideation in the population over time. These trajectories were estimated by GAMs applied to 36,106 Lifelines participants. The x-axis denotes time with the corresponding month and year shown. The grey rectangles highlight the three different nationwide lockdowns in the Netherlands.

#### Major depressive disorder

We found a significant non-linear trajectory for the point prevalence of MDD (Figure 2, Table S3). The prevalence of MDD was relatively stable during the months of 2020 and increased rapidly at the start of the third hard lockdown. We observed a peak in MDD prevalence of 2.88% (95% CI: 2.71%–3.06%) at the beginning of March 2021, which then declined again the months after. Comparing the end of April 2021 with the end of April 2020, the prevalence of MDD was higher one year later (2.41% versus 1.78%;). The trajectory of MDD was similar across sensitivity analyses (Figure S5 and Table S7, S11).

#### Anxiety symptoms

Anxiety symptoms showed a significant non-linear trajectory with the highest number of symptoms reported at the immediate start of the pandemic (Figure 2, Table S3). Reported anxiety symptoms declined as the first lockdown progressed and were lower during mid-summer 2020 when government restrictions were eased. The prevalence of symptoms increased again from August and plateaued during the second lockdown. Anxiety symptoms moderately increased during the third lockdown but did not reach the level of symptoms reported at the start of the pandemic. The prevalence of symptoms declined as the end of the third lockdown approached in May 2021.

Comparing the end of April 2021 with the end of April 2020, the number of reported anxiety symptoms one year later was slightly higher (0.65 versus 0.59). Trajectories observed in our sensitivity analyses aligned with our findings from our main analyses (Figure S6 and Table S7, S11).

#### General anxiety disorder

For GAD, we found a significant non-linear trajectory. The prevalence of GAD was high at the beginning of the pandemic and declined as the first lockdown progressed, reaching its lowest prevalence at the start of July 2020. The months after, the prevalence had a roughly linear increase reaching its peak prevalence of 2.92% (95% CI: 2.76%-3.08%) at the beginning of March 2021 and declined again toward the end of the third lockdown. Comparing the end of April 2021 with the end of April 2020, the prevalence of GAD was higher one year later (2.71% versus 2.16%). The non-linear trajectories were similar in sensitivity analyses in the full cohort as well as after taking into account individual- and family-specific effects (Figure S5 and S7 and Table S2, S6, S10). The GAD trajectory did show a more smoothed pattern in the selected 5K subsample (Figure S5), which was likely due to lower statistical power. In the subsample of only younger participants, we observed a more similar trajectory as that of our main analysis.

#### Suicidal ideation

We observed a significant linear increase in the prevalence of suicidal ideation (Figure 2 and Table S3). At the beginning of April 2020, the prevalence of suicidal ideation was 0.59% (95% CI: 0.56%-0.62%) which increased to 0.95% (95% CI: 0.89%-1.01%) during mid-June 2021. This represents a 1.61x increase in reported suicidal ideation in the population. Sensitivity analyses in the full cohort yielded similar results. After taking into account individual-specific and family-specific variation the trajectory of prevalence was flat over time, indicating that most individuals in the population did not experience an increase in suicidal ideation and that specific individuals or subgroups may be more at risk (Figure S8 and Table S3, S7, S11).

### Longitudinal trajectories across age, sex and history of MDD/GAD

As (symptoms of) MDD and GAD are known to be more prevalent in younger adults, women, and individuals with a previous diagnosis, we next investigated how the observed longitudinal trajectories differed by age, sex, and lifetime history of MDD/GAD in our sample.

#### Longitudinal trajectories by age

Younger participants reported significantly more depressive and anxiety symptoms, as well as a higher prevalence of MDD and GAD than older participants across all time points (Table S4, Figure S9-S12). The relative risk of young subjects compared to older subjects did not change across time for these four outcomes nor did they in our sensitivity analyses (Tables S4, S8, S12).

#### Suicidal ideation

At the start of the pandemic, younger participants also reported significantly more suicidal thoughts than older participants (e.g. 1.12% versus 0.52% in 20-versus 60-year old participants). Moreover, younger participants also reported a steeper increase in suicidal ideations over time as indicated by a significant interaction effect between time and age (Figure 3 and Table S4). By mid-June 2021, 20-year-old participants had 4.14x more reports of suicidal ideation than in March 2020 (4.64% (CI: 3.09%-6.96%) versus 1.12% (CI: 0.76%-1.66%)), whereas this increase was lower for older participants (i.e. 1.98x, 1.17x, 0.59x, 1.08x for 30-, 40-, 60-, and 80-year old participants, respectively). This interaction effect between time and age was also significant in our sensitivity analyses of the youngest 5,000 study participants, but not in our randomly selected subsample (Table S8 and S12, Figure S14 and S15). As suicidal ideation is a rarer phenotype, including more young adults increased our statistical power to detect such an effect.

**Figure 3.**
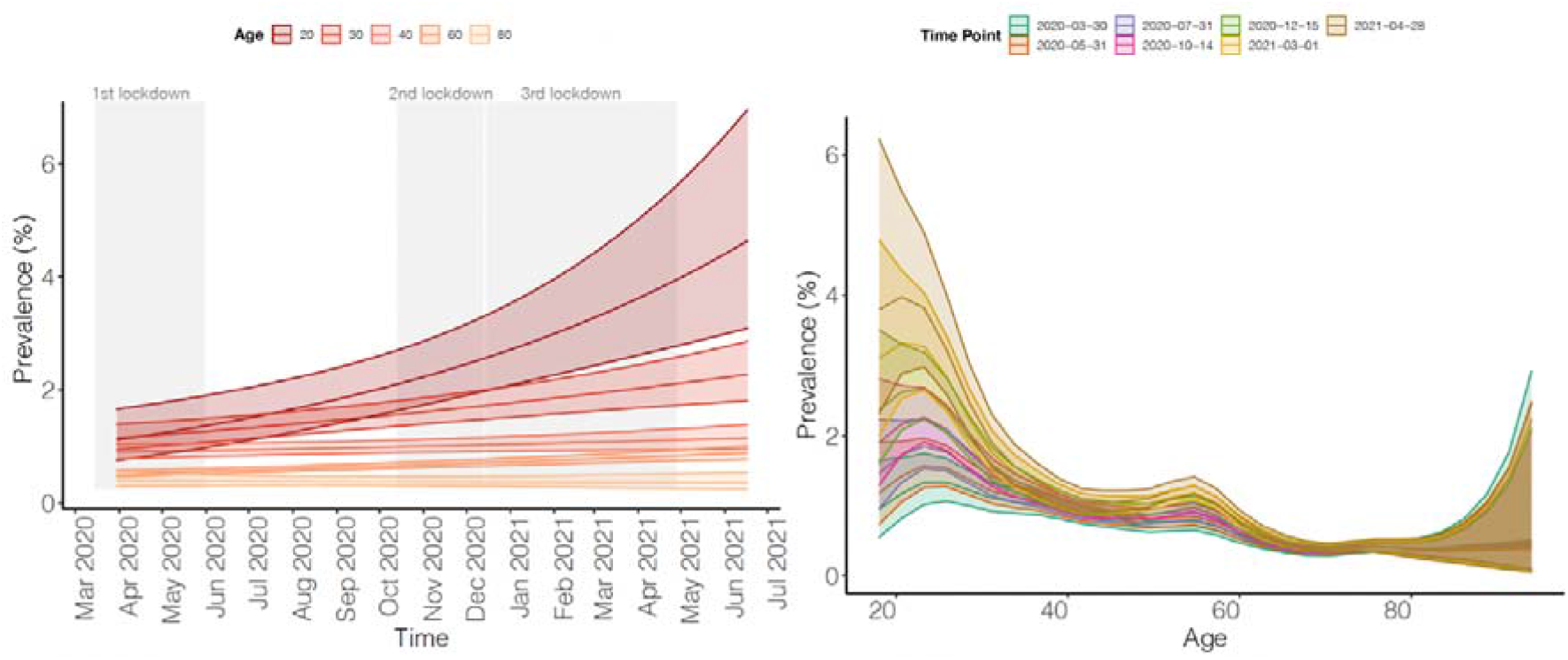
The trajectory of suicidal ideation by time and age during the COVID-19 pandemic. Shown are the results of interaction between time and age on reported suicidal ideation in our main sample of 36,106 participants (age 18-45). The left panels show the trajectory of suicidal ideation for specific ages over time. The right panels show the trajectory of specific time points across age. The legends at the top of the graph denoted color coding of groups. The gray rectangles highlight the three different nationwide lockdowns in the Netherlands.

#### Longitudinal trajectories by sex

As expected, the prevalence of MDD and GAD and their symptom scores were on average significantly higher in women than in men (similarly to findings in[10]), but the development of these outcomes over time did not differ between the sexes (Figure S16-S25 and Table S5, S9, S13). The prevalence and development of suicidal thoughts across time did not differ between women and men, which we also observed in our sensitivity analyses.

#### Longitudinal trajectories by lifetime history of MDD/GAD

Study participants with a lifetime history of MDD/GAD reported significantly more symptoms and a higher prevalence of MDD/GAD and suicidal ideation during the pandemic than participants without a previous diagnosis (Figure 4, Table S6). We found a significant difference in trajectories over time between participants with and without a history for MDD or GAD for depressive or anxiety symptom scores, respectively, while not for the other three outcomes (Table S6). For both depressive and anxiety symptom scores, the difference between participants with and without a lifetime diagnosis was greatest during periods out of lockdown when reported symptoms were lowest in the general population (Figure 4). Sensitivity analyses in the full cohort yielded similar findings, as did correcting for individual- and family-specific variation (Figure S26-S35 and Table S6, S10, S14).

**Figure 4.**
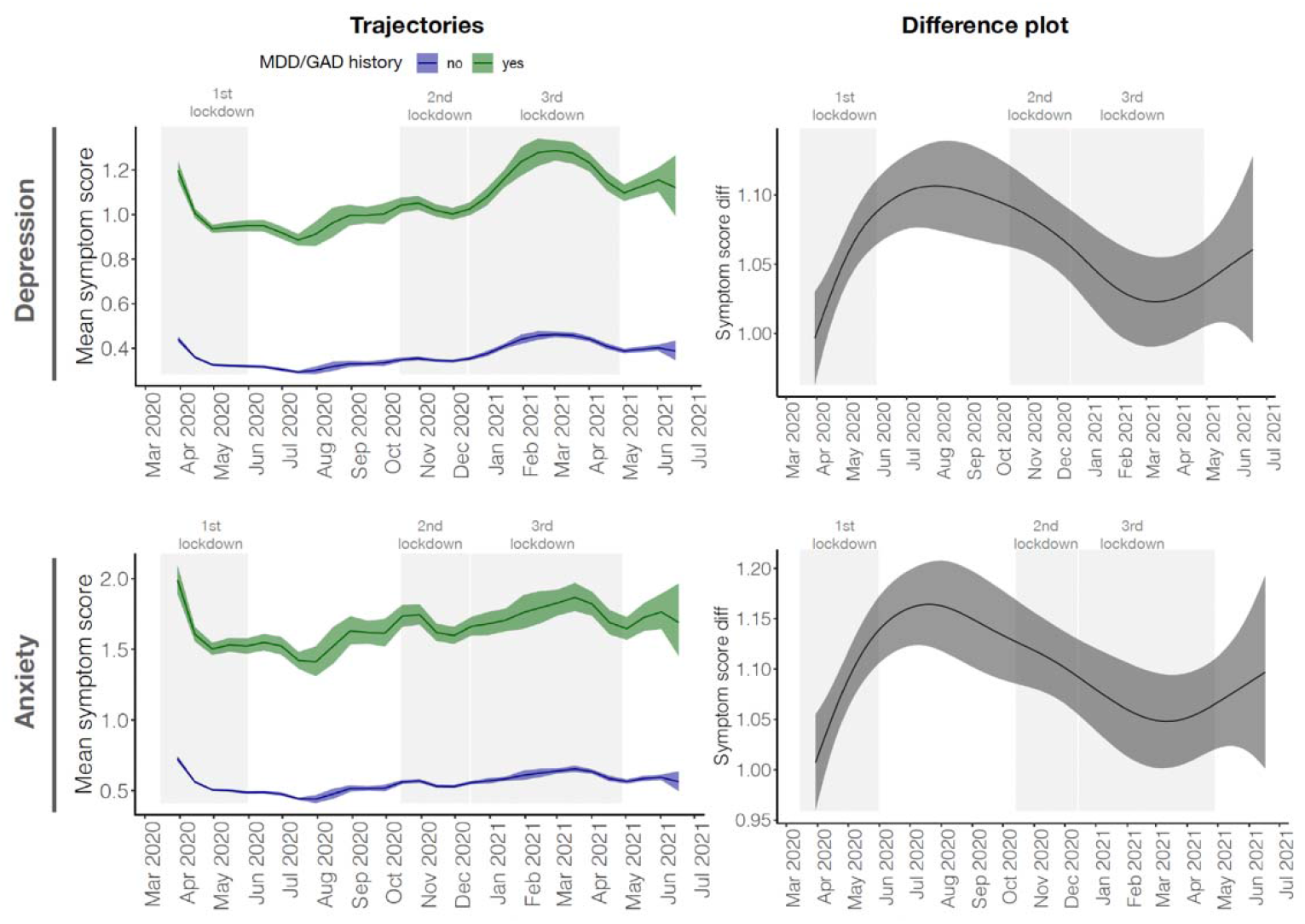
The trajectory of depressive and anxiety symptom scores between study participants with and without a history of MDD/GAD during the COVID-19 pandemic. Shown are the results of interaction between time and a lifetime history of MDD/GAD on symptom scores based on GAM analyses in our main analysis sample. The left panels show the trajectory of the mean score of reported depressive and anxiety symptoms between participants with (green) and without (blue) a history of MDD/GAD. The right panels show corresponding difference plots that visualize the trajectory of the difference between the two groups. The gray rectangles highlight the three different nationwide lockdowns in the Netherlands.

## Discussion

To investigate the impact of the COVID-19 pandemic on mental health, we described the development of current (symptoms of) MDD, GAD and suicidal thoughts in a longitudinal sample from the Northern Dutch general population who were followed for more than a year during the COVID-19 pandemic across three nation-wide lockdowns.

In general, we observed a greater prevalence of (symptoms of) MDD and GAD in the population during periods of lockdown and a declining prevalence during periods of eased or no restrictions, which suggests that the pandemic and government measures enacted to restrict spreading of the coronavirus indeed impacted the mental health of the general population. We observed a high prevalence of symptoms of MDD and GAD at the start of the pandemic followed by a rapid decline in the months after, which is in line with observations from other countries[3, 17–19]. A new finding of our study is that the prevalence of symptoms of MDD and GAD increased again during later lockdowns alongside a high prevalence of MDD and GAD during the third lockdown which peaked in March 2021. The third lockdown in the Netherlands, unlike the first and second lockdown, was a hard lockdown and characterized by stricter measures, including an evening curfew. Interestingly, we observed a plateauing of reported symptoms during the second lockdown, which was a shorter partial lockdown, where schools, sports, libraries and museums, unlike during the third lockdown, stayed open. Together our results suggest that lockdown measures impacted mental health in the population, although we submit that this is an observational study of multifactorial psychiatric conditions.

Women, younger adults, and subjects with pre-existing mental health conditions, were more at risk for developing MDD and GAD symptoms and disorders, which is in line with observations from previous studies as well[17–19]. However, apart from a difference in intercept, we observed no differences in the development of MDD/GAD over time across sex or age. We did find a difference in the trajectory of reported MDD/GAD symptoms between participants with a history of MDD/GAD compared to participants without a history, but this difference was not found for MDD and GAD. Participants with a history of MDD/GAD reported increased symptom severity during the end of the first lockdown and months after, which is in contrast to a smaller Dutch study that found no difference at the start of the pandemic[20] and highlights the importance of continued longitudinal measurements in large cohorts.

In contrast to the non-linear trajectories of (symptoms of) MDD and GAD, the prevalence of suicidal ideation showed an increasing linear trend among young adults while we did not observe this effect in the overall population. There were no sex differences in prevalence rates of suicidality, but subjects with a lifetime history of MDD reported more suicidal ideation. A meta-analysis of suicidality during the pandemic found increased rates of suicidal ideation and behaviors with a higher incidence in younger individuals as well[2]. As only 14% of studies on suicidality during the pandemic included children or young people[21], there is an urgent need for more epidemiological studies, like the Lifelines COVID-19 study, that includes these groups to investigate what factors are driving the observed increase. As psychological distress, low perceived social support, and loneliness are known factors that increase suicidality in adolescents and young adults[22, 23], government measures to reduce the spread of the virus by social distancing measures likely contributed to the increased prevalence. Adolescents and young adults were furthermore significantly impacted by unemployment at the start of the pandemic [24]. As unemployment increases risk of suicide[25], our observation of increased suicidal thoughts may translate to suicide attempts and mortality[2]. Indeed, the Dutch Suicide Prevention Center reported an increase in completed suicides among young adults in January and February of 2021 compared to previous years [26]. These alarming findings warrant for alertness in psychiatric care services and urge governments to consider the long-term impact of pandemic measurements on young people.

A key strength of our study is the high-quality and high-resolution longitudinal data collected using a validated structured diagnostic interview throughout the first fifteen months of the pandemic spanning three nationwide lockdowns. This allowed for the application of sophisticated nonlinear statistical models to investigate the development of MDD, GAD and suicidality across time. However, our findings should be interpreted considering several limitations. First, due to the large computational resources required to run GAMs with random effects, we were only able to account for individual- and family-specific variation in a subsample of our cohort. While this subsample was similar to our larger sample in its main characteristics, we cannot exclude that our analyses may have missed important insights due to limited statistical power, especially for more rare phenotypes. We did show that analyses on the youngest study participants is worthwhile to prioritize as a significant proportion of the variation in MDD/GAD outcomes lies in younger adults, which increases statistical power to identify group differences. Second, we fitted each outcome in a separate model and thus could only compare the prevalence within a single trajectory and not between trajectories of different outcomes. As depression and anxiety are known to have comorbidity, how changes in trajectories relate among outcomes is an important question to investigate in future research. Third, we did not have information on the prevalence of MDD, GAD and suicidality in the year before the start of the pandemic and thus could not account for that, nor for seasonal effects. Fourth, we assessed current symptoms of GAD within the past seven and fourteen days. We therefore did not assess GAD according to the DSM-IV criteria that requires symptoms to be present for at least six months. Fifth, we did not account for corona infection status of study participants. As the number of participants who reported to have tested positive for the coronavirus during the study was relatively low (11.2%), we expect infection status and its accompanying symptoms to have minimal impact on our findings, if any at all, given that the observed trajectories in prevalence of MDD/GAD and suicidal ideation also do not follow the number of reported infections over time (Figure 1). Sixth, Lifelines participants are more often female, middle aged, married, and Dutch native compared to the population in the North of the Netherlands [27]. We therefore cannot exclude that there are population sub-groups who may be at greater risk of declining mental health that are under-represented in the Lifelines COVID 19 study. Finally, our findings should be interpreted within the societal context of the study. The Netherlands had three lockdown periods with different characteristics and is furthermore on average a rich country with a social welfare system.

In summary, we investigated the development of (symptoms of) MDD and GAD and suicidal thoughts in the Northern Dutch population during the COVID-19 pandemic and observed higher prevalence during periods of lockdown, in particular the third hard lockdown. We furthermore found an alarming linear increase in suicidal thoughts among young adults that warrants for alertness in psychiatric care services. Further studies are needed to investigate mechanisms underlying these rising prevalence rates. Our findings provide important insights into the impact of the pandemic on the mental health of the population, which can help guide policy makers and clinical care during future lockdowns and epi/pandemics.

## Data Availability

All data were obtained from the Lifelines biobank under project application OV20_00021. Data can be obtained by submitting a request to the Lifelines Research Office. The data are not publicly available because they contain information that could compromise the study participants' privacy and consent.

https://www.lifelines.nl/

## Author contributions

All authors contributed to design of the study. The Lifelines Corona Research Initiative, AO and HvL were involved in data collection. AO, HvL and MW designed the study. AO performed the statistical analyses. AO drafted the manuscript; all other authors interpreted results and provided feedback on analyses and drafts of the manuscript.

## Acknowledgements

The Lifelines Biobank initiative has been made possible by funding from the Dutch Ministry of Health, Welfare and Sport, the Dutch Ministry of Economic Affairs, the University Medical Center Groningen (UMCG the Netherlands), the University of Groningen, the Northern Provinces of the Netherlands, FES (Fonds Economische Structuurversterking), SNN (Samenwerkingsverband Noord Nederland) and REP (Ruimtelijk Economisch Programma).

We acknowledge funding for the LifeLines Corona Research project from the University of Groningen and the University Medical Centre Groningen. The authors wish to acknowledge the efforts of the Lifelines Corona Research Initiative and the following initiatives participants:

HM Boezen† (1), Jochen O. Mierau (2,3), Lude H. Franke (4), Jackie Dekens (4,6), Patrick Deelen (4), Pauline Lanting (4), Judith M. Vonk (1), Ilja Nolte (1), Anil P.S. Ori (4,5), Annique Claringbould (4), Floranne Boulogne (4), Marjolein X.L. Dijkema (4), Henry H. Wiersma (4), Robert Warmerdam (4), Soesma A. Jankipersadsing (4), Irene van Blokland (4,7)

1. Department of Epidemiology, University of Groningen, University Medical Center Groningen, Groningen, The Netherlands
2. Faculty of Economics and Business, University of Groningen, Groningen, The Netherlands
3. Aletta Jacobs School of Public Health, Groningen, The Netherlands
4. Department of Genetics, University of Groningen, University Medical Center Groningen, Groningen, The Netherlands
5. Department of Psychiatry, University of Groningen, University Medical Center Groningen, Groningen, The Netherlands
6. Center of Development and Innovation, University of Groningen, University Medical Center Groningen, Groningen, The Netherlands
7. Department of Cardiology, University of Groningen, University Medical Center Groningen, Groningen, The Netherlands

We thank the UMCG Genomics Coordination Center, the UG Center for Information Technology and their sponsors BBMRI-NL & TarGet for storage and computing infrastructure.

