## Supplemental_Text for "Longitudinal analyses of depression and anxiety highlight greater prevalence during COVID-19 lockdowns in the Dutch general population and a continuing increase in suicidal ideation in young adults"

### Supplemental methods

#### *COVID-19 questionnaires and sample selection*

Lifelines includes 167,729 participants who are all residents of three northern provinces of the Netherlands. Lifelines, when adjusted for differences in demographic composition, is broadly representative for the adult population of the north of the Netherlands (Klijs et al. 2015). In March 2020, the first digital questionnaire was sent to all 140,145 adult Lifelines participants with an e-mail address on file (Intyre et al. 2021). Follow-up questionnaires were initially sent on a weekly (questionnaires 1 to 6, Q1-Q6) and later on a biweekly and monthly basis (Q7-Q23). From Q8, 111,239 participants who opened at least one email-invite were invited to fill out future questionnaires. From Q18, 65,504 participants were invited who filled in at least two previous questionnaires.

Digital questionnaires were designed by a multidisciplinary group of researchers and assessed a range of physical and mental health outcomes, including (symptoms of) MDD and GAD. A shortened version of the MINI was incorporated in the digital questionnaires. That is, three items that overlap between the MDD and GAD DSM-IV criteria (i.e., feelings of fatigue, difficulty concentrating, and problems with sleeping) were only assessed once to minimize questionnaire length and participant burden (we used a combined item for insomnia and hypersomnia). Questionnaire 12 (July 24 - Aug 08 2020) was excluded from our analyses as the MINI was not assessed in this questionnaire.

#### *Missing data and imputation*

To handle missing data, we performed a single dataset imputation using a chained equation approach and a flexible multiple regression framework as implemented in the mice_v3.13 package in R_v4.0.3 (van Buuren and Groothuis-Oudshoorn 2011). Missing data was either due to (1) a non-response for an entire questionnaire at one of the time points, (2) a non-response for a specific item while the participants did fill out the rest of that questionnaire, or due to (3) a specific item not being included in a questionnaire and therefore being missing because of the design of a questionnaire. Having suicidal thoughts was for example only assessed at monthly intervals to limit the burden on participants. If a participant did not fill out an entire questionnaire (1), the symptoms on this time point were not imputed. In case of a specific item being missing because a participant did not answer or because of the design of the questionnaire (2 or 3), we assumed that these data were *missing at random*, and we used imputation. Across all questionnaires, an average of 0.71% of participants had missing data across MDD and GAD symptoms (SD=1.11%, min=0.11%, max=6.41%). For suicidal ideation, the average percentage of participants with missing data was 0.95% (SD=1.73%). For lifetime history of MDD and GAD, 37.08% and 37.10% of participants had data missing. All participants had information on age and sex available. The missing data was subsequently imputed using both information from other time points within the Lifelines COVID-19 study as well as from previous assessment waves in Lifelines. Briefly, predictors for imputation were selected using the quickpred() function with a minimum threshold of proportion of usable cases of 0.25 and a minimum absolute correlation of 0.08. The mice() function was then used to generate a single imputed dataset using default settings.

#### *Participant dropout*

We investigated if participant dropouts impacted our analyses. A drop out was defined by being missing at the next questionnaire, which indicates that a participant who filled out a questionnaire at a specific time point “X” did not fill out a questionnaire at the next time point

“X+1”. If a participant was also missing at time point x+2, we define their dropout status at time point x+2 as NA. Thus, dropout status refers solely to missing at the next questionnaire. To investigate the relationship between MD/GAD outcomes and participant dropouts, we first implemented logistic regression models. Participant dropout was defined as missing at next questionnaires and used as the response variable. MDD diagnoses, MDD symptom sum score, suicidality, GAD diagnoses, GAD symptom sum score were used as predictors with age and sex as covariates. For each predictor, a separate logistic regression model was constructed as following; glm(missing_at_nextQ ~ predictor of interest + age + sex, family=”binomial”). In a second analysis, we estimated the prevalence of dropouts at the next questionnaire over time using a generalized additive model. Analyses were run in both the main sample (N=36,106) as well as the full cohort (N=76,016).

***Predictor measures***

Calendar age and sex were extracted from the Lifelines database and were available for all study participants. For age, we used participants’ age at the first COVID-19 questionnaire. To determine sex, Lifelines was linked to personal records of municipalities in the Netherlands that store sex assigned at birth based on physical sexual characteristics.

***Statistical analyses - generalized additive models***

Generalized additive models (GAMs) were used to assess the prevalence of MDD and GAD outcomes over time and their association with age, sex, and lifetime history of MDD/GAD. GAMs are regression models that allow for identification of complex non-linear trajectories in dynamically varying data (Hastie and Tibshirani 1986), such as time series data, by automatically determining the optimal combination of nonlinear basis functions (e.g. linear terms, polynomial terms, cubic terms, etc.) (Wood 2017; Wieling 2018). GAMs have a flexible mixed-effect regression framework that allows for the modelling of covariates and dependencies within participants. Overfitting is prevented by minimizing a combination of the error and a non-linearity penalty (Wieling 2018). All analyses were performed in R_v4.0.3 using the packages mgcv_1.8.33 (Wood 2017) and itsadug_2.4 (van Rij et al. 2016). We modeled the prevalence of each of the five MDD/GAD outcomes as a (potentially) non-linear function of time and tested if there were significant interaction effects of time with age, sex, and lifetime history of MDD/GAD. We analyzed each predictor separately as we did not want to correct our analyses for known risk factors of depression and anxiety (i.e. age, sex, and history of MDD/GAD). The prevalence of MDD, GAD, and suicidal ideation, which were binary outcomes, were modelled using a binomial link function (family=”binomial”), while MDD and GAD symptom scores, which are non-gaussian continuous outcomes, were modelled using a negative binomial link function (family=”nb”). In total, we ran 20 separate GAMs (5 dependent outcome variables x 4 independent variables) using the follow syntax:

Time model: MDD/GAD outcome variable ~ s(Time)
Age model: MDD/GAD outcome variable ~ s(Time) + s(Age) + ti(Time, Age)
Sex model: MDD/GAD outcome variable ~ Sex + s(Time) + s(Time, by=Sex)
LH model1: MDD outcome variable ~ MDD LH + s(Time) + s(Time, by=MDD LH)
LH model2: GAD outcome variable ~ GAD LH + s(Time) + s(Time, by=GAD LH)

The GAMs with sex and lifetime history (LH) as independent variables were implemented with an intercept term that estimated the constant difference between predictor groups and a single ordered factor difference smooth that estimates the differences in trajectory. The GAMs with age as predictor use a tensor product smooth, which can model two-dimensional patterns. Using this approach, we model a non-linear interaction between time and age by allowing the coefficients underlying the smooth for time to vary non-linearly depending on the value age. Mixed-effect models with the inclusion of a random intercept and a linear random slope for subject and family were implemented in a sub sample of the cohort using the following syntax: s(subject.id, bs='re') + s(family.id, bs='re') + s(subject.id, assessment_day, bs='re') + s(family.id, assessment_day, bs='re'). Smooth terms were implemented with k=20 and model parameters checked with the gam.check() function. For fast implementation, all models were executed using fast restricted maximum likelihood and discretization (method="fREML", discrete=TRUE).

***Sensitivity analyses and computational limits of mixed-effect GAM analyses***

As mixed-effect GAMs require significant computational resources we were unable to run such analyses using on our high computing cluster using our full sample. While we were able to run a fast implementation of GAMs with fixed-effects using discretization and fast REML (1GB RAM and 2 minutes computational time), GAMs with random-effects required significantly more computational memory and time. To determine the limits of our computing cluster given its available resources, we tested multiple model parameters to evaluate how to best run GAMs on the Lifeline dataset. We tested three important parameters: model complexity, sample size, and parallel computing. All GAMs were run using fREML and discretization. We found that analyses with a sample size of 5,000 individuals with the inclusion of linear random-effects (to correct for within-subject and within-family dependencies) completed within 24 hours using a maximum of 96GB of RAM. Analyses that included more individuals and/or more complex terms, such as non-linear random-effects and correction for autocorrelation, required significantly more RAM than available for a single job on the computing cluster. We were furthermore unable to speed up computational time by using multithreading, which seemed to be due to incompatibility between the mgcv package and parallel computing settings on the cluster. We therefore decided not to implement multithreading with our GAM analysis. Thus, based on our testing of model parameter and cluster computational settings, we chose to run GAMs with the inclusion of a random intercept and linear random slope in 5,000 randomly selected as a sensitivity analysis to assess how individual- and family-specific variation impacted our results. The mixed-effect GAMs do estimate lower prevalence as these models set the random effects to zero and only return fixed effects. They however do allow us to evaluate the significance of the predictor effects and compare the trajectories between models with and without random effects.

### Supplemental Results

***Participant dropout in main select sample***

For most study participants we did not have complete data for all twenty-three questionnaires. Our main selected sample had a total of 629,811 questionnaires across 36,106 participants. This means that about ~24% of questionnaires were not filled out and therefore missing. In the full cohort, 47% of questionnaires were missing. To investigate if our five phenotypic outcomes were associated with missingness in our sample, we assessed if they predicted a participant to be missing at the next questionnaire. The table below presents the results of the logistic regression analyses. Regression coefficient estimates and corresponding confidence intervals are exponentiated.

Results Dropout Analysis

| term | All samples | | | | Select subsample | | | |
|---|---|---|---|---|---|---|---|---|
| | estimate | conf.low | conf.high | p.value | estimate.1 | conf.low.1 | conf.high.1 | p.value.1 |
| **MDD diagnosis** | | | | | | | | |
| (Intercept) | 1.27 | 1.24 | 1.31 | 2.53e-71 | 0.49 | 0.47 | 0.50 | 3.32e-302 |
| MDD diagnoses | 1.32 | 1.28 | 1.37 | 4.18e-56 | 1.25 | 1.19 | 1.32 | 2.28e-19 |
| Age | 0.97 | 0.97 | 0.97 | 0.00e+00 | 0.98 | 0.98 | 0.98 | 0.00e+00 |
| Sex - Female | 0.93 | 0.92 | 0.94 | 2.17e-31 | 1.00 | 0.98 | 1.01 | 8.50e-01 |
| **MDD Sum Score** | | | | | | | | |
| (Intercept) | 1.20 | 1.17 | 1.24 | 1.25e-41 | 0.47 | 0.45 | 0.48 | 0.00e+00 |
| MDD sum score | 1.07 | 1.07 | 1.08 | 1.06e-227 | 1.06 | 1.05 | 1.06 | 2.84e-70 |
| Age | 0.97 | 0.97 | 0.97 | 0.00e+00 | 0.98 | 0.98 | 0.98 | 0.00e+00 |
| Sex - Female | 0.92 | 0.91 | 0.93 | 9.48e-43 | 0.99 | 0.97 | 1.01 | 1.98e-01 |
| **Suicidality** | | | | | | | | |
| (Intercept) | 1.29 | 1.26 | 1.33 | 8.39e-80 | 0.49 | 0.47 | 0.51 | 6.95e-295 |
| Suicidality | 1.11 | 1.05 | 1.18 | 6.11e-04 | 1.11 | 1.02 | 1.21 | 1.11e-02 |
| Age | 0.97 | 0.97 | 0.97 | 0.00e+00 | 0.98 | 0.98 | 0.98 | 0.00e+00 |
| Sex - Female | 0.94 | 0.93 | 0.95 | 5.41e-30 | 1.00 | 0.98 | 1.02 | 9.53e-01 |
| **GAD diagnosis** | | | | | | | | |
| (Intercept) | 1.27 | 1.24 | 1.31 | 4.46e-70 | 0.48 | 0.47 | 0.50 | 7.36e-305 |
| GAD diagnoses | 1.31 | 1.27 | 1.35 | 1.65e-62 | 1.27 | 1.22 | 1.33 | 2.29e-26 |
| Age | 0.97 | 0.97 | 0.97 | 0.00e+00 | 0.98 | 0.98 | 0.98 | 0.00e+00 |
| Sex - Female | 0.93 | 0.92 | 0.94 | 5.77e-32 | 1.00 | 0.98 | 1.01 | 7.87e-01 |
| **GAD Sum Score** | | | | | | | | |
| (Intercept) | 1.18 | 1.15 | 1.21 | 2.33e-33 | 0.46 | 0.44 | 0.48 | 0.00e+00 |
| GAD sum score | 1.07 | 1.06 | 1.07 | 1.08e-224 | 1.05 | 1.04 | 1.05 | 1.99e-66 |
| Age | 0.97 | 0.97 | 0.97 | 0.00e+00 | 0.98 | 0.98 | 0.98 | 0.00e+00 |
| Sex - Female | 0.92 | 0.91 | 0.93 | 7.58e-45 | 0.99 | 0.97 | 1.00 | 1.47e-01 |

The five MD/GAD outcomes are shown stacked by the rows. Association statistics are shown for analyses conducted in both the full lifelines sample (N=76,016) as well as the selected main analysis sample (N=36,106).

We observe that all five outcomes significantly predict a participant to be missing at the next questionnaire. Participants who meet the DSM-IV criteria of MDD and GAD at a specific questionnaire show the highest odds, i.e. 1.32 and 1.31, respectively, for being missing at the next questionnaire compared to participants who do not meet this criteria. Participants who report suicidal ideations have an odds of 1.11 to be missing at the next questionnaire compared to those who did not report these thoughts. Finally, a one-unit increase in depressive and anxiety sum score gives an odds of 1.07 to be missing at the next questionnaire. The odds ratios were slightly lower in our main analysis sample compared to the full cohort, which is expected as we selected our main analysis sample to have participants with more complete data across the twenty-three questionnaires. These participants were however still more likely to be missing at the next questionnaire when they would report (symptoms of) MDD and GAD but do return back to the study at later questionnaires.

We next estimated the prevalence of a participant being missing at the next questionnaire over the duration of the study using GAMs (see figure below). We found higher rates of missingness during the summer months of 2020 (Jul-Sep) and at the beginning of 2021 (Jan-Mar) during the hard lockdown.

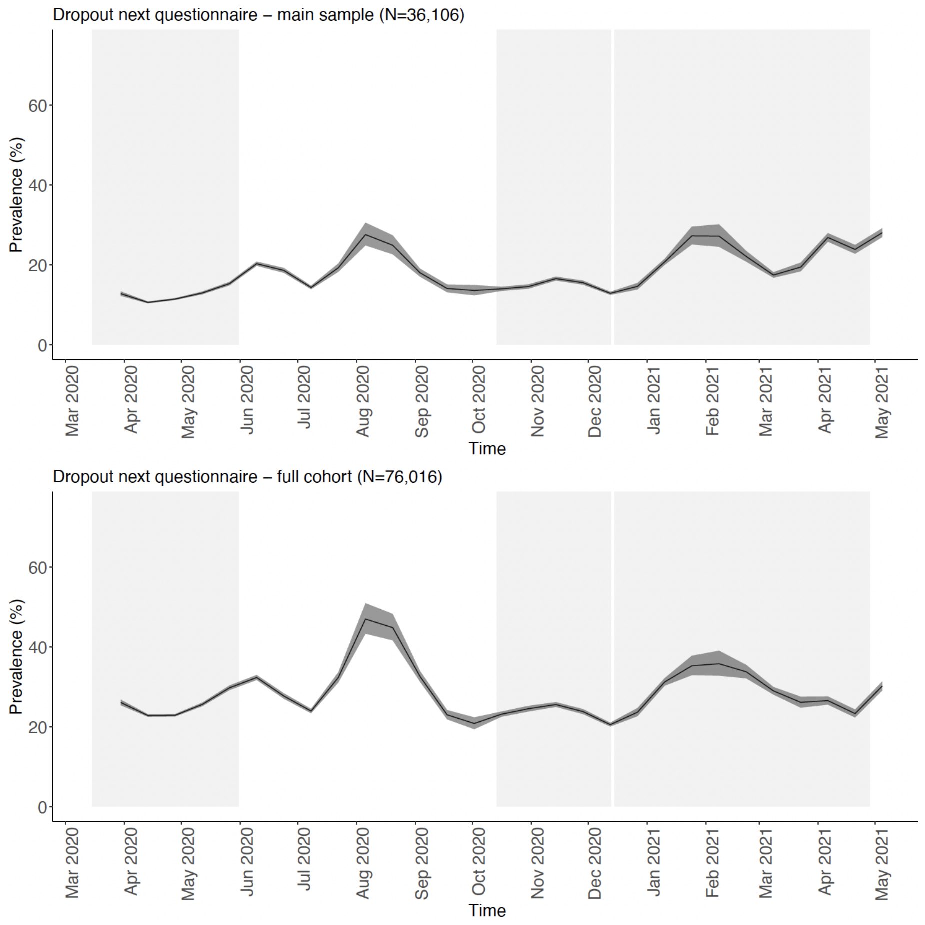

GAM analyses that model the effect of the tensor product between time and age on missingness furthermore showed that younger adults dropped out more than older adults (>60 years), except during the summer months of 2020 where participants across all ages dropped out. Younger adults dropped out more frequently during the final months of the study as well (see figure below).

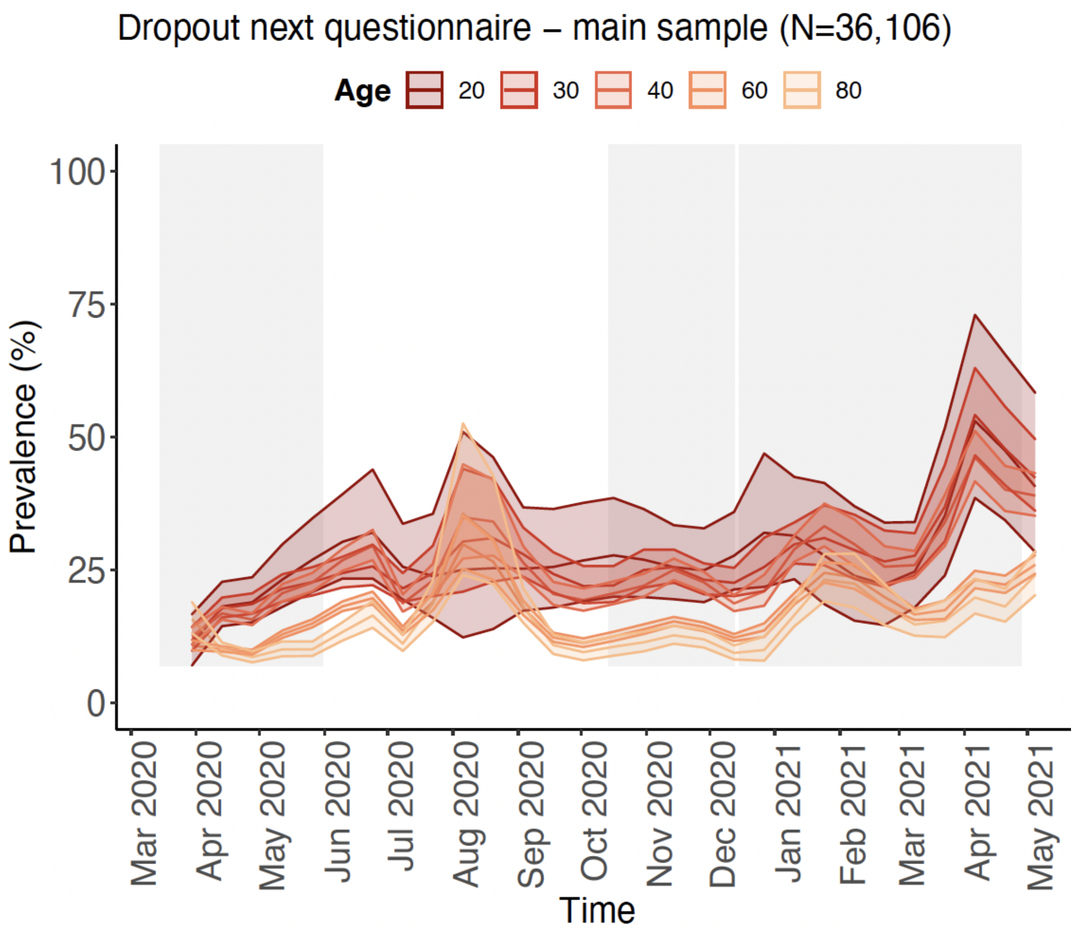

In summary, our findings show that participant dropout, defined by being missing at the next questionnaire, is relatively stable over time with two periods of higher prevalence during the months of Jul-Sep 2020 and Jan-Mar 2021. Our five phenotypic outcomes each significantly predicted being missing at the next questionnaire even in our main analyses sample that was selected for study participants with more complete data. This suggests that our estimated prevalence of (symptoms of) MDD/GAD and suicidal thoughts likely present a minimum observed prevalence and that the true prevalence might be higher. We expect this to be the case especially for our estimates during the third hard lockdown and the final months of our study, where young adults were significantly more often missing at the next questionnaire.
