## Supplemental_Tables for "Longitudinal analyses of depression and anxiety highlight greater prevalence during COVID-19 lockdowns in the Dutch general population and a continuing increase in suicidal ideation in young adults"

### Table S1. Questionnaire items

| Language | MINI questionnaire Item | Answer | Symptom | Module |
|---|---|---|---|---|
| Dutch | 1. Hebt u zich tijdens de afgelopen 7/14 dagen voortdurend somber of depressief gevoeld gedurende het grootste gedeelte van de dag, en dit bijna elke dag? | Ja/nee | Somberheid | MD |
| English | 1. In the last 7/14 days have you felt low or depressed for much of the day, every day? | Yes/no | Depressed | |
| Dutch | 2. Hebt u tijdens de afgelopen 7/14 dagen voortdurend het gevoel gehad nergens meer zin in te hebben of geen interesse meer te hebben voor dingen die u normaal wel interesseren? | Ja/nee | Anhedonie | MD |
| English | 2. In the last 7/14 days have you had the feeling that you've lost interest in or the will to do things you are normally interested in? | Yes/no | Anhedonia | |
| Dutch | 3. In de afgelopen 7/14 dagen, was uw eetlust merkbaar veranderd, of is uw gewicht toegenomen of afgenomen, zonder dat dit de bedoeling was? | Ja/nee | Eetlust/gewicht | MD |
| English | 3. In the last 7/14 days, did your appetite change noticeably, or did your weight increase or decrease without this being intended? | Yes/no | Appetite/weight | |
| Dutch | 4. In de afgelopen 7/14 dagen, hebt u bijna elke nacht slaapproblemen gehad (moeilijk inslapen, wakker worden tijdens de nacht of te vroeg in de ochtend, of juist te veel slapen)? | Ja/nee | Slaapproblemen | MD & GAD |
| English | 4. In the last 7/14 days, have you had problems sleeping almost every night (difficulty falling asleep, waking up in the night or too early in the morning, or actually sleeping too much)? | Yes/no | Difficulty sleeping | |
| Dutch | 5. In de afgelopen 7/14 dagen, praatte of bewoog u trager dan gewoonlijk, of voelde u zich juist rusteloos, gejaagd en kon u moeilijk stil blijven zitten? Bijna elke dag? | Ja/nee | Rusteloosheid | MD |
| English | 5. In the last 7/14 days, did you speak or move more slowly than normal? Or did you feel restless, jittery and could barely sit still? Nearly every day? | Yes/no | Restlessness | |
| Dutch | 6. In de afgelopen 7/14 dagen, voelde u zich bijna elke dag waardeloos of schuldig? | Ja/nee | Waardeloosheid | MD |
| English | 6. In the last 7/14 days, did you feel worthless or guilty almost every day? | Yes/no | Worthlessness | |
| Dutch | 7. In de afgelopen 7/14 dagen, kon u zich bijna elke dag moeilijk concentreren of moeilijk beslissingen nemen? | Ja/nee | Concentratieproblemen | MD & GAD |
| English | 7. In the last 7/14 days, was it difficult to concentrate or make decisions almost every day? | Yes/no | Difficulty concentrating | |
| Dutch | 8. In de afgelopen 7/14 dagen, hebt u overwogen zichzelf iets aan te doen, wenste u dat u dood was, of had u zelfmoordgedachten? | Ja/nee | Zelfmoordgedachten | MD |

| | | | | |
|---|---|---|---|---|
| English | 8. In the last 7/14 days, have you considered hurting yourself, wished you were dead, or had suicidal thoughts? | Yes/no | Suicideal ideation | |
| Dutch | 9. In de afgelopen 14 dagen, voelde u zich bijna elke dag moe of futloos? | Ja/nee | Vermoeidheid | MD & GAD |
| English | 9. In the last 14 days, did you feel tired or without energy almost every day? | Yes/no | Feelings of fatigue | |
| Dutch | 10. Hebt u in de afgelopen 7/14 dagen buitensporig gepiekerd en zich zorgen gemaakt over meerdere problemen van het dagelijks leven, op het werk, thuis, in uw naaste omgeving? | Ja/nee | Buitensporig piekeren | GAD |
| English | 10. In the last 7/14 days, have you been worrying excessively and worrying about multiple problems of every day life, at work, at home, in your immediate environment? | Yes/no | Excessive worrying | |
| Dutch | 11. Zijn deze zorgen bijna elke dag aanwezig in de afgelopen 7/14 dagen? | Ja/nee | - | GAD |
| English | 11. Were these worries present almost every day in the last 7/14 days? | Yes/no | - | |
| Dutch | 12. Vindt u het moeilijk om deze bezorgdheid in de hand te houden of belemmert dit u om zich te concentreren de afgelopen 7/14 dagen? | Ja/nee | - | GAD |
| English | 12. In the last 7/14 days did you find it hard to set these worries aside or did they prevent you from concentrating? | Yes/no | - | |
| Dutch | 13. U zich rusteloos, geladen of zenuwachtig voelde? / Gebeurde het in de afgelopen 7 dagen vaak dat | Ja/nee | Rusteloosheid/ nerveusheid | GAD |
| English | 14. You felt restless, jittery or nervous? / In the last 7/14 days did it often happen that | Yes/no | Restlessness/nervousness | |
| Dutch | 15. U zich gespannen voelde? / Gebeurde het in de afgelopen 7/14 dagen vaak dat | Ja/nee | Gespannenheid | GAD |
| English | 15. You felt tense? / In the last 7/14 days did it often happen that | Yes/no | Feeling tense | |
| Dutch | 16. U bijzonder prikkelbaar was? / Gebeurde het in de afgelopen 7/14 dagen vaak dat | Ja/nee | Prikkelbaar | GAD |
| English | 16. You were particularly irritable? / In the last 7/14 days did it often happen that | Yes/no | Irritable | |

| | Full sample | Selected sample | Random 5K sample | Youngest 5K sample |
|---|---|---|---|---|
| Number of participants | 76,016 | 36,106 | 5,000 | 5,000 |
| Age | 53.7 (SD=13.0) | 57.4 (SD=11.9) | 57.5 (SD=11.8) | 36.6 (SD=6.3) |
| 18-30 years | 3,884 (5.1%) | 875 (2.4%) | 114 (2.3%) | 875 (17.5%) |
| 31-67 years | 60,617 (79.7%) | 27,394 (75.9%) | 3,806 (76.1%) | 4,125 (82.5%) |
| >68 years | 11,515 (15.1%) | 7,837 (21.7%) | 1,080 (21.6%) | - |
| Female (%) | 46,253 (60.8%) | 22,339 (61.9%) | 3,067 (61.3%) | 3,508 (70.2) |
| Total questionnaires | 823,478 | 629,811 | 87,076 | 78,054 |
| Median questionnaire/person (IQR 25%-75%) | 10 (3-20) | 20 (15-22) | 20 (15-22) | 18 (12-22) |
| Lifetime MDD (%) | 17,814 (23.4%) | 7,917 (21.9%) | 1,095 (21.9%) | 1,468 (29.4%) |
| Lifetime GAD (%) | 7,011 (9.2%) | 2,997 (8.3%) | 399 (8.0%) | 547 (10.9%) |
| Number of MDD cases | 6,555 | 3,675 | 489 | 753 |
| Average MDD prevalence | 2.2% | 1.9% | 1.7% | 3.2% |
| Average MDD symptom score | 0.54 (SD=1.19) | 0.50 (SD=1.14) | 0.49 (SD=1.11) | 0.71 (SD=1.40) |
| Number of GAD cases | 8,004 | 4,625 | 642 | 970 |
| Average GAD prevalence | 2.6% | 2.3% | 2.2% | 3.9% |
| Average GAD symptom score | 0.67 (SD=1.34) | 0.63 (SD=1.29) | 0.61 (SD=1.26) | 0.99 (SD=1.61) |
| Number of suicidal ideation cases | 3,150 | 2,007 | 279 | 394 |
| Average suicidal ideation prevalence | 0.78% | 0.71% | 0.70% | 1.19% |

**Table S2. Demographics and characteristics of the full cohort and different sample subsets.** Shown are the number of participants and their characteristics for the full cohort, our selected sample, the random sample, and the youngest 5K sample. Number of MDD/GAD and suicidal ideation cases are presented as the number of participants who met the DSM-IV criteria during at least one questionnaire. The MDD/GAD prevalence and average symptom scores are presented by the mean and spread of their per-questionnaire average based on the imputed data.

**Table S3. GAM estimates of MD and GAD outcome trajectories in the main sample and full cohort over time.** Shown are the statistical output of the GAMs for each of the five outcome variables for both the analyses performed in the main sample (left columns) as well as the full cohort (right columns). The prevalence was modeled using a single smooth term of time (in days) as predictor without adjusting for dependencies within subject and family. Shown are the estimate and standard error (SE) of the intercept and the fitted smooth of the time variable with corresponding test statistics. The effective degree of freedom (edf) is a proxy for the degree of non-linearity of the smooth. An edf of 1.0 indicates a linear relationship, while an edf > 1.0 indicates non-linearity. P-values of the smooth terms of interest are adjusted by Bonferroni correction. Edf.df = reference degree of freedom, Z = z-score, Chi.sq= chi-square, P = p-value, t = t-value, F = F-value.

| | Main analysis sample (N=36K) | | | | Full sample (N=76K) | | | |
|---|---|---|---|---|---|---|---|---|
| **MDD** | **Estimate** | **SE** | **Z** | **P** | **Estimate** | **SE** | **Chi.sq** | **P** |
| (Intercept) | -3.90 | 0.01 | -390.6 | <2e-16 | -3.79 | 0.01 | -430.26 | <2e-16 |
| **Smooth terms** | **edf** | **edf.df** | **Chi.sq** | **P** | **edf** | **edf.df** | **Chi.sq** | **P** |
| s(Time) | 8.15 | 9.95 | 350.86 | <2e-16 | 9.42 | 11.36 | 268.26 | <2e-16 |
| **MDD score** | **Estimate** | **SE** | **t** | **P** | **Estimate** | **SE** | **t** | **P** |
| (Intercept) | -0.68 | 0.00 | -197.61 | <2e-16 | -0.61 | 0.00 | -195.58 | <2e-16 |
| **Smooth terms** | **edf** | **edf.df** | **F** | **P** | **edf** | **edf.df** | **F** | **P** |
| s(Time) | 15.10 | 16.95 | 93.63 | <2e-16 | 15.47 | 17.24 | 101.33 | <2e-16 |
| **Suicidal ideation** | **Estimate** | **SE** | **Z** | **P** | **Estimate** | **SE** | **Z** | **P** |
| (Intercept) | -4.90 | 0.02 | -314.36 | <2e-16 | -4.81 | 0.01 | -349.65 | <2e-16 |
| **Smooth terms** | **edf** | **edf.df** | **Chi.sq** | **P** | **edf** | **edf.df** | **Chi.sq** | **P** |
| s(Time) | 1.00 | 1.00 | 97.05 | <2e-16 | 1.00 | 1.00 | 89.64 | <2e-16 |
| **GAD** | **Estimate** | **SE** | **Z** | **P** | **Estimate** | **SE** | **Z** | **P** |
| (Intercept) | -3.73 | 0.01 | -407.06 | <2e-16 | -3.63 | 0.01 | -442.57 | <2e-16 |
| **Smooth terms** | **edf** | **edf.df** | **Chi.sq** | **P** | **edf** | **edf.df** | **Chi.sq** | **P** |
| s(Time) | 7.11 | 8.75 | 197.04 | <2e-16 | 9.28 | 11.2 | 205.28 | <2e-16 |
| **GAD score** | **Estimate** | **SE** | **t** | **P** | **Estimate** | **SE** | **t** | **P** |
| (Intercept) | -0.45 | 0.00 | -133.07 | <2e-16 | -0.40 | 0.00 | -128.64 | <2e-16 |
| **Smooth terms** | **edf** | **edf.df** | **F** | **P** | **edf** | **edf.df** | **F** | **P** |
| s(Time) | 16.30 | 17.89 | 78.61 | <2e-16 | 16.31 | 17.88 | 113.38 | <2e-16 |

**Table S4. GAM estimates of MD and GAD outcome trajectories by time and age in the main sample and full cohort.** Shown are the statistical output of the GAMs for each of the five outcome variables for both the analyses performed in the main sample (left columns) as well as the full cohort (right columns). The prevalence was modeled using a single smooth term of time (in days), a single smooth term of age (in years), and a tensor product interaction smooth term between time and age as predictors without adjusting for dependencies within subject and family. Shown are the estimate and standard error (SE) of the intercept and the fitted smooths for each term in the model. The effective degree of freedom (edf) is a proxy for the degree of non-linearity of the smooth. An edf of 1.0 indicates a linear relationship, while an edf > 1.0 indicates non-linearity. P-values of the smooth terms of interest are adjusted by Bonferroni correction. Edf.df = reference degree of freedom, Z = z-score, Chi.sq= chi-square, P = p-value, t = t-value, F = F-value.

| | Main analysis sample (N=36K) | | | | Full sample (N=76K) | | | |
|---|---|---|---|---|---|---|---|---|
| **MDD** | **Estimate** | **SE** | **Z** | **P** | **Estimate** | **SE** | **Z** | **P** |
| (Intercept) | -3.79 | 0.03 | -119.60 | <2e-16 | -3.72 | 0.04 | -91.81 | <2e-16 |
| **Smooth terms** | **edf** | **edf.df** | **Chi.sq** | **P** | **edf** | **edf.df** | **Chi.sq** | **P** |
| s(Time) | 8.09 | 9.88 | 355.04 | <2e-16 | 9.13 | 11.04 | 270.47 | <2e-16 |
| s(Age) | 15.54 | 17.15 | 1631.81 | <2e-16 | 13.45 | 15.43 | 2113.77 | <2e-16 |
| ti(Time, Age) | 3.56 | 4.43 | 8.19 | 1.00 | 4.59 | 5.72 | 20.27 | 0.04 |
| **MDD score** | **Estimate** | **SE** | **t** | **P** | **Estimate** | **SE** | **t** | **P** |
| (Intercept) | -0.59 | 0.01 | -57.69 | <2e-16 | -0.54 | 0.01 | -38.24 | <2e-16 |
| **Smooth terms** | **edf** | **edf.df** | **F** | **P** | **edf** | **edf.df** | **F** | **P** |
| s(Time) | 15.25 | 17.08 | 92.56 | <2e-16 | 15.51 | 17.27 | 45.17 | <2e-16 |
| s(Age) | 15.12 | 16.82 | 443.95 | <2e-16 | 14.79 | 16.47 | 564.46 | <2e-16 |
| ti(Time, Age) | 4.18 | 5.20 | 6.34 | 9.42e-05 | 20.05 | 29.89 | 1.91 | 0.04 |
| **Suicidal ideation** | **Estimate** | **SE** | **Z** | **P** | **Estimate** | **SE** | **Z** | **P** |
| (Intercept) | -4.87 | 0.05 | -93.36 | <2e-16 | -4.76 | 0.06 | -82.57 | <2e-16 |
| **Smooth terms** | **edf** | **edf.df** | **Chi.sq** | **P** | **edf** | **edf.df** | **Chi.sq** | **P** |
| s(Time) | 1.00 | 1.00 | 12.57 | 7.88e-03 | 1.00 | 1.00 | 4.80 | 0.03 |
| s(Age) | 10.34 | 12.48 | 741.24 | <2e-16 | 10.92 | 13.07 | 970.91 | <2e-16 |
| ti(Time, Age) | 5.59 | 7.42 | 27.00 | 9.72e-03 | 6.11 | 8.56 | 26.19 | 0.04 |
| **GAD** | **Estimate** | **SE** | **Z** | **P** | **Estimate** | **SE** | **Z** | **P** |
| (Intercept) | -3.79 | 0.04 | -93.48 | <2e-16 | -3.77 | 0.05 | -70.84 | <2e-16 |
| **Smooth terms** | **edf** | **edf.df** | **Chi.sq** | **P** | **edf** | **edf.df** | **Chi.sq** | **P** |
| s(Time) | 7.12 | 8.76 | 125.19 | <2e-16 | 9.23 | 11.15 | 125.1 | <2e-16 |
| s(Age) | 13.84 | 15.73 | 2265.94 | <2e-16 | 13.81 | 15.62 | 2726.16 | <2e-16 |
| ti(Time, Age) | 1.70 | 2.18 | 0.70 | 1.00 | 11.66 | 16.81 | 31.67 | 0.32 |
| **GAD score** | **Estimate** | **SE** | **t** | **P** | **Estimate** | **SE** | **t** | **P** |
| (Intercept) | -0.4 | 0.01 | -40.9 | <2e-16 | -0.37 | 0.01 | -25.67 | <2e-16 |
| **Smooth terms** | **edf** | **edf.df** | **F** | **P** | **edf** | **edf.df** | **F** | **P** |
| s(Time) | 16.39 | 17.95 | 76.70 | <2e-16 | 16.33 | 17.9 | 60.18 | <2e-16 |
| s(Age) | 14.44 | 16.29 | 697.65 | <2e-16 | 14.78 | 16.44 | 842.48 | <2e-16 |
| ti(Time, Age) | 3.23 | 4.03 | 2.75 | 0.51 | 19.87 | 29.32 | 1.84 | 0.07 |

**Table S5. GAM estimates of MD and GAD outcome trajectories by time and sex in the main sample and full cohort.** Shown are the statistical output of the GAMs for each of the five outcome variables for both the analyses performed in the main sample (left columns) as well as the full cohort (right columns). The prevalence was modeled using an ordered factor difference smooth of sex across time (in days) as predictor without adjusting for dependencies within subject and family. Shown are the estimate and standard error (SE) of the intercept and the fitted smooths for each term in the model. The effective degree of freedom (edf) is a proxy for the degree of non-linearity of the smooth. An edf of 1.0 indicates a linear relationship, while an edf > 1.0 indicates non-linearity. P-values of the smooth terms of interest are adjusted by Bonferroni correction. Edf.df = reference degree of freedom, Z = z-score, Chi.sq= chi-square, P = p-value, t = t-value, F = F-value.

| | Main analysis sample (N=36K) | | | | Full sample (N=76K) | | | |
|---|---|---|---|---|---|---|---|---|
| **MDD** | **Estimate** | **SE** | **Z** | **P** | **Estimate** | **SE** | **Z** | **P** |
| (Intercept) | -4.11 | 0.02 | -236.93 | <2e-16 | -4.00 | 0.02 | -262.87 | <2e-16 |
| Sex - women | 0.32 | 0.02 | 15.46 | <2e-16 | 0.34 | 0.02 | 18.56 | <2e-16 |
| **Smooth terms** | **edf** | **edf.df** | **Chi.sq** | **P** | **edf** | **edf.df** | **Chi.sq** | **P** |
| s(Time) | 8.06 | 9.84 | 160.33 | <2e-16 | 9.4 | 11.33 | 162.69 | <2e-16 |
| s(Time):sex - women | 2.32 | 2.86 | 5.72 | 1.00 | 2.15 | 2.66 | 7.86 | 0.75 |
| **MDD score** | **Estimate** | **SE** | **t** | **P** | **Estimate** | **SE** | **t** | **P** |
| (Intercept) | -0.97 | 0.01 | -177.45 | <2e-16 | -0.90 | 0.00 | -182.01 | <2e-16 |
| Sex - women | 0.43 | 0.01 | 67.37 | <2e-16 | 0.43 | 0.01 | 73.78 | <2e-16 |
| **Smooth terms** | **edf** | **edf.df** | **F** | **P** | **edf** | **edf.df** | **F** | **P** |
| s(Time) | 15.25 | 17.07 | 49.85 | <2e-16 | 15.59 | 17.33 | 59.24 | <2e-16 |
| s(Time):sex - women | 4.20 | 5.22 | 2.67 | 0.44 | 4.05 | 5.03 | 2.72 | 0.38 |
| **Suicidal ideation** | **Estimate** | **SE** | **Z** | **P** | **Estimate** | **SE** | **Z** | **P** |
| (Intercept) | -4.94 | 0.03 | -193.67 | <2e-16 | -4.86 | 0.02 | -215.94 | <2e-16 |
| Sex - women | 0.06 | 0.03 | 1.77 | 0.08 | 0.08 | 0.03 | 2.69 | 7.18e-03 |
| **Smooth terms** | **edf** | **edf.df** | **Chi.sq** | **P** | **edf** | **edf.df** | **Chi.sq** | **P** |
| s(Time) | 1.00 | 1.00 | 52.15 | <2e-16 | 1.00 | 1.00 | 61.47 | <2e-16 |
| s(Time):sex - women | 1.00 | 1.00 | 2.37 | 1.00 | 1.00 | 1.00 | 6.86 | 0.18 |
| **GAD** | **Estimate** | **SE** | **Z** | **P** | **Estimate** | **SE** | **Z** | **P** |
| (Intercept) | -3.99 | 0.02 | -243.99 | <2e-16 | -3.9 | 0.01 | -267.44 | <2e-16 |
| Sex - women | 0.40 | 0.02 | 20.71 | <2e-16 | 0.41 | 0.02 | 24.04 | <2e-16 |
| **Smooth terms** | **edf** | **edf.df** | **Chi.sq** | **P** | **edf** | **edf.df** | **Chi.sq** | **P** |
| s(Time) | 7.13 | 8.77 | 103.58 | <2e-16 | 9.32 | 11.25 | 136.95 | <2e-16 |
| s(Time):sex - women | 1.79 | 2.21 | 2.28 | 1.00 | 1.98 | 2.45 | 4.19 | 1.00 |
| **GAD score** | **Estimate** | **SE** | **t** | **P** | **Estimate** | **SE** | **t** | **P** |
| (Intercept) | -0.76 | 0.01 | -144.85 | <2e-16 | -0.69 | 0.00 | -147.13 | <2e-16 |
| Sex - women | 0.45 | 0.01 | 74.78 | <2e-16 | 0.45 | 0.01 | 81.44 | <2e-16 |
| **Smooth terms** | **edf** | **edf.df** | **F** | **P** | **edf** | **edf.df** | **F** | **P** |
| s(Time) | 16.38 | 17.94 | 73.23 | <2e-16 | 16.36 | 17.92 | 115.57 | <2e-16 |
| s(Time):sex - women | 1.01 | 1.01 | 1.40 | 1.00 | 1.02 | 1.03 | 0.21 | 1.00 |

**Table S6. GAM estimates of MD and GAD outcome trajectories by time and lifetime history of MD/GAD in the main sample and full cohort.** Shown are the statistical output of the GAMs for each of the five outcome variables for both the analyses performed in the main sample (left columns) as well as the full cohort (right columns). The prevalence was modeled using an ordered factor difference smooth of MDD or GAD lifetime history (LT) across time (in days) as predictor without adjusting for dependencies within subject and family. Shown are the estimate and standard error (SE) of the intercept and the fixed effect of MDD/GAD LT as well as the fitted smooth of the time variable and the ordered factor difference smooth of MDD/GAD LT with corresponding test statistics. The effective degree of freedom (edf) is a proxy for the degree of non-linearity of the smooth. An edf of 1.0 indicates a linear relationship, while an edf > 1.0 indicates non-linearity. P-values of the smooth terms of interest are adjusted by Bonferroni correction. Edf.df = reference degree of freedom, Z = z-score, Chi.sq= chi-square, P = p-value, t = t-value, F = F-value.

| | **Main analysis sample (N=36K)** | | | | **Full sample (N=76K)** | | | |
|---|---|---|---|---|---|---|---|---|
| **MDD** | **Estimate** | **SE** | **Z** | **P** | **Estimate** | **SE** | **Z** | **P** |
| (Intercept) | -4.69 | 0.02 | -292.21 | <2e-16 | -4.56 | 0.01 | -322.04 | <2e-16 |
| MDD_LT - yes | 1.93 | 0.02 | 96.34 | <2e-16 | 1.90 | 0.02 | 107.07 | <2e-16 |
| **Smooth terms** | **edf** | **edf.df** | **Chi.sq** | **P** | **edf** | **edf.df** | **Chi.sq** | **P** |
| s(Time) | 8.04 | 9.81 | 213.8 | <2e-16 | 9.26 | 11.17 | 166.95 | <2e-16 |
| s(Time):MDD_LT-yes | 1.94 | 2.40 | 4.56 | 1.00 | 3.31 | 4.11 | 11.04 | 0.62 |
| **MDD score** | **Estimate** | **SE** | **t** | **P** | **Estimate** | **SE** | **t** | **P** |
| (Intercept) | -1.02 | 0.00 | -265.21 | <2e-16 | -0.96 | 0 | -272.03 | <2e-16 |
| MDD_LT - yes | 1.06 | 0.01 | 162.69 | <2e-16 | 1.06 | 0.01 | 177.56 | <2e-16 |
| **Smooth terms** | **edf** | **edf.df** | **F** | **P** | **edf** | **edf.df** | **F** | **P** |
| s(Time) | 15.14 | 16.98 | 89.22 | <2e-16 | 15.59 | 17.33 | 101.59 | <2e-16 |
| s(Time):MDD_LT-yes | 4.68 | 5.81 | 5.04 | 1.40e-03 | 6.71 | 8.26 | 9.09 | <2e-16 |
| **Suicidal ideation** | **Estimate** | **SE** | **Z** | **P** | **Estimate** | **SE** | **Z** | **P** |
| (Intercept) | -5.70 | 0.03 | -217.75 | <2e-16 | -5.63 | 0.02 | -241.08 | <2e-16 |
| MDD_LT - yes | 1.93 | 0.03 | 58.94 | <2e-16 | 1.94 | 0.03 | 66.95 | <2e-16 |
| **Smooth terms** | **edf** | **edf.df** | **Chi.sq** | **P** | **edf** | **edf.df** | **Chi.sq** | **P** |
| s(Time) | 1.00 | 1.00 | 20.84 | 1.17e-04 | 1.00 | 1.00 | 23.07 | 4.28e-05 |
| s(Time):MDD_LT-yes | 1.00 | 1.00 | 2.88 | 1.00 | 1.00 | 1.01 | 1.86 | 1.00 |
| **GAD** | **Estimate** | **SE** | **Z** | **P** | **Estimate** | **SE** | **Z** | **P** |
| (Intercept) | -4.07 | 0.01 | -364.5 | <2e-16 | -3.99 | 0.01 | -398.1 | <2e-16 |
| GAD_LT - yes | 1.89 | 0.02 | 97.5 | <2e-16 | 1.9 | 0.02 | 110.48 | <2e-16 |
| **Smooth terms** | **edf** | **edf.df** | **Chi.sq** | **P** | **edf** | **edf.df** | **Chi.sq** | **P** |
| s(Time) | 7.19 | 8.83 | 174.88 | <2e-16 | 9.19 | 11.1 | 185.49 | <2e-16 |
| s(Time):GAD_LT-yes | 2.95 | 3.67 | 9.47 | 1.00 | 3.20 | 3.97 | 11.57 | 0.45 |
| **GAD score** | **Estimate** | **SE** | **t** | **P** | **Estimate** | **SE** | **t** | **P** |
| (Intercept) | -0.6 | 0.00 | -172.21 | <2e-16 | -0.55 | 0.00 | -173.14 | <2e-16 |
| GAD_LT - yes | 1.10 | 0.01 | 116.88 | <2e-16 | 1.09 | 0.01 | 129.10 | <2e-16 |
| **Smooth terms** | **edf** | **edf.df** | **F** | **P** | **edf** | **edf.df** | **F** | **P** |
| s(Time) | 16.51 | 18.03 | 86.75 | <2e-16 | 16.55 | 18.05 | 124.58 | <2e-16 |
| s(Time):GAD_LT-yes | 4.55 | 5.66 | 4.74 | 3.42e-03 | 6.26 | 7.74 | 6.46 | <2e-16 |

**Table S7. GAM estimates of MD and GAD outcome trajectories with and without correction for within-subject and -family dependencies.** Shown are the statistical output of the GAMs for each of the five outcome variables for analyses performed in a random sample of 5,000 study participants selected from the main analyses sample of 36,106 individuals. The prevalence was modeled using a single smooth term of time (in days) as predictor without (left columns) and with (right columns) correction for dependencies within subject and family. Shown are the estimate and standard error (SE) of the intercept and the fitted smooth of the time variable with corresponding test statistics. The effective degree of freedom (edf) is a proxy for the degree of non-linearity of the smooth. An edf of 1.0 indicates a linear relationship, while an edf > 1.0 indicates non-linearity. P-values of the smooth terms of interest are adjusted by Bonferroni correction. Edf.df = reference degree of freedom, Z = z-score, Chi.sq= chi-square, P = p-value, t = t-value, F = F-value.

| | No correction for subject and family dependencies (N=5K) | | | | With correction for subject and family dependencies (N=5K) | | | |
|---|---|---|---|---|---|---|---|---|
| **MDD** | **Estimate** | **SE** | **Z** | **P** | **Estimate** | **SE** | **Z** | **P** |
| (Intercept) | -4.04 | 0.03 | -142.68 | <2e-16 | -5.04 | 0.05 | -93.92 | <2e-16 |
| **Smooth terms** | **edf** | **edf.df** | **Chi.sq** | **P** | **edf** | **edf.df** | **Chi.sq** | **P** |
| s(Time) | 4.89 | 6.06 | 63.33 | <2e-16 | 5.52 | 6.84 | 52.62 | 8.46e-08 |
| s(Subject.id) | - | - | - | - | 927.80 | 5000 | 5835.33 | <2e-16 |
| s(Family.id) | - | - | - | - | 365.79 | 4609 | 1466.89 | 5.31e-03 |
| s(Time, Subject.id) | - | - | - | - | 201.46 | 4999 | 838.56 | 8.62e-15 |
| s(Time, Family.id) | - | - | - | - | 0.05 | 4608 | 0.05 | 0.50 |
| **MDD score** | **Estimate** | **SE** | **t** | **P** | **Estimate** | **SE** | **t** | **P** |
| (Intercept) | -0.71 | 0.01 | -83.46 | <2e-16 | -1.67 | 0.03 | -65.01 | <2e-16 |
| **Smooth terms** | **edf** | **edf.df** | **F** | **P** | **edf** | **edf.df** | **F** | **P** |
| s(Time) | 10.22 | 12.23 | 18.08 | <2e-16 | 12.41 | 14.4 | 19.69 | <2e-16 |
| s(Subject.id) | - | - | - | - | 3186.49 | 5000 | 99.5 | 3.48e-12 |
| s(Family.id) | - | - | - | - | 688.67 | 4609 | 16.69 | 1.00 |
| s(Time, Subject.id) | - | - | - | - | 1873.56 | 4999 | 25.3 | 1.00E-10 |
| s(Time, Family.id) | - | - | - | - | 0.02 | 4608 | 0 | 0.786 |
| **Suicidal ideation** | **Estimate** | **SE** | **Z** | **P** | **Estimate** | **SE** | **Z** | **P** |
| (Intercept) | -4.90 | 0.04 | -117.25 | <2e-16 | -5.66 | 0.07 | -86.68 | <2e-16 |
| **Smooth terms** | **edf** | **edf.df** | **Chi.sq** | **P** | **edf** | **edf.df** | **Chi.sq** | **P** |
| s(Time) | 1.00 | 1.00 | 19.85 | 1.78e-04 | 1.00 | 1.00 | 0.02 | 1.00 |
| s(Subject.id) | - | - | - | - | 665.82 | 5000 | 3391.99 | <2e-16 |
| s(Family.id) | - | - | - | - | 0.02 | 4609 | 0.02 | 0.54 |
| s(Time, Subject.id) | - | - | - | - | 177.25 | 4999 | 677.88 | <2e-16 |
| s(Time, Family.id) | - | - | - | - | 0.02 | 4608 | 0.02 | 0.50 |
| **GAD** | **Estimate** | **SE** | **Z** | **P** | **Estimate** | **SE** | **Z** | **P** |
| (Intercept) | -3.78 | 0.02 | -152.56 | <2e-16 | -4.75 | 0.05 | -98.00 | <2e-16 |
| **Smooth terms** | **edf** | **edf.df** | **Chi.sq** | **P** | **edf** | **edf.df** | **Chi.sq** | **P** |
| s(Time) | 2.37 | 2.93 | 14.03 | 0.08 | 2.26 | 2.79 | 6.34 | 1.00 |
| s(Subject.id) | - | - | - | - | 1472.61 | 5000 | 7988.08 | <2e-16 |
| s(Family.id) | - | - | - | - | 0.06 | 4609 | 0.06 | 0.53 |
| s(Time, Subject.id) | - | - | - | - | 296.05 | 4999 | 1420.35 | <2e-16 |
| s(Time, Family.id) | - | - | - | - | 0.03 | 4608 | 0.03 | 0.55 |
| **GAD score** | **Estimate** | **SE** | **t** | **P** | **Estimate** | **SE** | **t** | **P** |
| (Intercept) | -0.49 | 0.01 | -59.71 | <2e-16 | -1.52 | 0.03 | -57.33 | <2e-16 |
| **Smooth terms** | **edf** | **edf.df** | **F** | **P** | **edf** | **edf.df** | **F** | **P** |
| s(Time) | 10.02 | 12.01 | 16.20 | <2e-16 | 12.65 | 14.63 | 30.00 | <2e-16 |
| s(Subject.id) | - | - | - | - | 3616.56 | 5000 | 105.47 | <2e-16 |

| | | | | | | | | |
|---|---|---|---|---|---|---|---|---|
| s(Family.id) | - | - | - | - | 417.65 | 4609 | 8.09 | 1.00 |
| s(Time, Subject.id) | - | - | - | - | 1985.19 | 4999 | 34.03 | 5.93e-09 |
| s(Time, Family.id) | - | - | - | - | 0.70 | 4608 | 0.00 | 1.00 |

**Table S8. GAM estimates of MD and GAD outcome trajectories by time and age and without correction for within-subject and -family dependencies.** Shown are the statistical output of the GAMs for each of the five outcome variables for analyses performed in a random sample of 5,000 study participants selected from the main analyses sample of 36,106 individuals. The prevalence was modeled using a single smooth term of time (in days), a single smooth term of age (in years), and a tensor product interaction smooth term between time and age as predictors without (left columns) and with (right columns) correction for dependencies within subject and family. Shown are the estimate and standard error (SE) of the intercept and the fitted smooths for each term in the model. The effective degree of freedom (edf) is a proxy for the degree of non-linearity of the smooth. An edf of 1.0 indicates a linear relationship, while an edf > 1.0 indicates non-linearity. P-values of the smooth terms of interest are adjusted by Bonferroni correction. Edf.df = reference degree of freedom, Z = z-score, Chi.sq= chi-square, P = p-value, t = t-value, F = F-value.

| | Sub sample without correction for subject and family dependencies (N=5K) | | | | Sub sample with correction for subject and family dependencies (N=5K) | | | |
|---|---|---|---|---|---|---|---|---|
| **MDD** | **Estimate** | **SE** | **Z** | **P** | **Estimate** | **SE** | **Z** | **P** |
| (Intercept) | -4.71 | 0.33 | -14.17 | <2e-16 | -4.97 | 0.05 | -92.26 | <2e-16 |
| **Smooth terms** | **edf** | **edf.df** | **Chi.sq** | **P** | **edf** | **edf.df** | **Chi.sq** | **P** |
| s(Time) | 4.87 | 6.04 | 62.37 | <2e-16 | 5.51 | 6.82 | 51.73 | 1.25e-07 |
| s(Age) | 15.07 | 16.4 | 334.72 | <2e-16 | 1.00 | 1.00 | 50.6 | 2.28e-11 |
| ti(Time, Age) | 1.00 | 1.00 | 6.06 | 0.28 | 1.00 | 1.00 | 2.19 | 1.00 |
| s(Subject.id) | - | - | - | - | 986.59 | 5000 | 6178.22 | <2e-16 |
| s(Family.id) | - | - | - | - | 278.01 | 4609 | 947.54 | 0.04 |
| s(Time, Subject.id) | - | - | - | - | 200.04 | 4999 | 817.16 | 1.38e-14 |
| s(Time, Family.id) | - | - | - | - | 0.08 | 4608 | 0.08 | 0.50 |
| **MDD score** | **Estimate** | **SE** | **t** | **P** | **Estimate** | **SE** | **t** | **P** |
| (Intercept) | -0.64 | 0.02 | -27.94 | <2e-16 | -4.9 | 0.05 | -99.82 | <2e-16 |
| **Smooth terms** | **edf** | **edf.df** | **F** | **P** | **edf** | **edf.df** | **F** | **P** |
| s(Time) | 10.57 | 12.59 | 10.91 | <2e-16 | 4.67 | 5.79 | 9.53 | 1.18e-08 |
| s(Age) | 16.59 | 17.98 | 66.91 | <2e-16 | 1.00 | 1.00 | 53.29 | 5.78e-12 |
| ti(Time, Age) | 15.25 | 21.92 | 1.73 | 0.34 | 1.00 | 1.00 | 3.69 | 1.00 |
| s(Subject.id) | - | - | - | - | 924.82 | 5000 | 1.52 | 3.5e-08 |
| s(Family.id) | - | - | - | - | 306.75 | 4609 | 0.31 | 1.00 |
| s(Time, Subject.id) | - | - | - | - | 23.62 | 4999 | 0.01 | 0.94 |
| s(Time, Family.id) | - | - | - | - | 0.03 | 4608 | 0.00 | 0.50 |
| **Suicidal ideation** | **Estimate** | **SE** | **Z** | **P** | **Estimate** | **SE** | **Z** | **P** |
| (Intercept) | -4.83 | 0.04 | -115.32 | <2e-16 | -5.61 | 0.07 | -85.31 | <2e-16 |
| **Smooth terms** | **edf** | **edf.df** | **Chi.sq** | **P** | **edf** | **edf.df** | **Chi.sq** | **P** |
| s(Time) | 1.00 | 1.00 | 20.44 | 1.41e-04 | 1.00 | 1.00 | 0.01 | 1.00 |
| s(Age) | 1.00 | 1.00 | 62.00 | <2e-16 | 1.00 | 1.00 | 24.79 | 1.29e-05 |
| ti(Time, Age) | 1.98 | 2.47 | 3.44 | 1.00 | 2.13 | 2.69 | 3.53 | 1.00 |
| s(Subject.id) | - | - | - | - | 651.77 | 5000 | 3299.7 | <2e-16 |
| s(Family.id) | - | - | - | - | 0.02 | 4609 | 0.02 | 0.55 |
| s(Time, Subject.id) | - | - | - | - | 177.34 | 4999 | 672.04 | 5.44e-11 |
| s(Time, Family.id) | - | - | - | - | 0.03 | 4608 | 0.03 | 0.50 |
| **GAD** | **Estimate** | **SE** | **Z** | **P** | **Estimate** | **SE** | **Z** | **P** |
| (Intercept) | -4.00 | 0.14 | -27.64 | <2e-16 | -4.73 | 0.10 | -47.71 | <2e-16 |
| **Smooth terms** | **edf** | **edf.df** | **Chi.sq** | **P** | **edf** | **edf.df** | **Chi.sq** | **P** |
| s(Time) | 2.20 | 2.72 | 13.54 | 1.00 | 2.17 | 2.67 | 5.23 | 1.00 |
| s(Age) | 13.76 | 15.64 | 400.12 | <2e-16 | 5.08 | 5.85 | 90.16 | <2e-16 |
| ti(Time, Age) | 1.01 | 1.01 | 1.07 | 1.00 | 1.00 | 1.00 | 0.20 | 1.00 |
| s(Subject.id) | - | - | - | - | 1418.27 | 5000 | 7514.83 | <2e-16 |

| | | | | | | | | |
|---|---|---|---|---|---|---|---|---|
| s(Family.id) | - | - | - | - | 0.04 | 4609 | 0.04 | 0.56 |
| s(Time, Subject.id) | - | - | - | - | 294.24 | 4999 | 1363.86 | 5.22e-19 |
| s(Time, Family.id) | - | - | - | - | 0.02 | 4608 | 0.02 | 0.57 |
| **GAD score** | **Estimate** | **SE** | **t** | **P** | **Estimate** | **SE** | **t** | **P** |
| (Intercept) | -0.50 | 0.02 | -20.37 | <2e-16 | -1.41 | 0.05 | -26.91 | <2e-16 |
| **Smooth terms** | **edf** | **edf.df** | **F** | **P** | **edf** | **edf.df** | **F** | **P** |
| s(Time) | 10.40 | 12.41 | 15.11 | <2e-16 | 12.56 | 14.53 | 19.35 | <2e-16 |
| s(Age) | 14.97 | 16.80 | 101.51 | <2e-16 | 5.94 | 6.16 | 46.04 | <2e-16 |
| ti(Time, Age) | 2.88 | 3.58 | 1.34 | 1.00 | 7.06 | 10.55 | 1.26 | 1.00 |
| s(Subject.id) | - | - | - | - | 3527.38 | 5000 | 96.8 | <2e-16 |
| s(Family.id) | - | - | - | - | 454.4 | 4609 | 8.09 | 1.00 |
| s(Time, Subject.id) | - | - | - | - | 1982.6 | 4999 | 31.2 | 9.63e-10 |
| s(Time, Family.id) | - | - | - | - | 1.24 | 4608 | 0.00 | 1.00 |

**Table S9. GAM estimates of MD and GAD outcome trajectories by time and sex and without correction for within-subject and -family dependencies.** Shown are the statistical output of the GAMs for each of the five outcome variables for analyses performed in a random sample of 5,000 study participants selected from the main analyses sample of 36,106 individuals. The prevalence was modeled using an ordered factor difference smooth of sex across time (in days) as predictor without (left columns) and with (right columns) correction for dependencies within subject and family. Shown are the estimate and standard error (SE) of the intercept and the fitted smooths for each term in the model. The effective degree of freedom (edf) is a proxy for the degree of non-linearity of the smooth. An edf of 1.0 indicates a linear relationship, while an edf > 1.0 indicates non-linearity. P-values of the smooth terms of interest are adjusted by Bonferroni correction. Edf.df = reference degree of freedom, Z = z-score, Chi.sq= chi-square, P = p-value, t = t-value, F = F-value.

| | **Sub sample without correction for subject and family dependencies (N=5K)** | | | | **Sub sample with correction for subject and family dependencies (N=5K)** | | | |
|---|---|---|---|---|---|---|---|---|
| **MDD** | **Estimate** | **SE** | **Z** | **P** | **Estimate** | **SE** | **Z** | **P** |
| (Intercept) | -4.16 | 0.05 | -88.31 | <2e-16 | -5.21 | 0.09 | -57.66 | <2e-16 |
| Sex - women | 0.18 | 0.06 | 3.14 | 1.67e-03 | 0.27 | 0.11 | 2.39 | 0.02 |
| **Smooth terms** | **edf** | **edf.df** | **Chi.sq** | **P** | **edf** | **edf.df** | **Chi.sq** | **P** |
| s(Time) | 4.17 | 5.18 | 35.41 | 4.66e-05 | 4.82 | 5.97 | 25.56 | 4.82e-03 |
| s(Time):sex - women | 3.29 | 4.08 | 11.03 | 0.55 | 3.85 | 4.78 | 11.43 | 0.86 |
| s(Subject.id) | - | - | - | - | 972.36 | 5000 | 6132.87 | <2e-16 |
| s(Family.id) | - | - | - | - | 318.56 | 4609 | 1169.36 | 0.020 |
| s(Time, Subject.id) | - | - | - | - | 195.42 | 4998 | 789.86 | 2.68e-14 |
| s(Time, Family.id) | - | - | - | - | 0.05 | 4608 | 0.05 | 0.50 |
| **MDD score** | **Estimate** | **SE** | **t** | **P** | **Estimate** | **SE** | **t** | **P** |
| (Intercept) | -0.96 | 0.01 | -68.91 | <2e-16 | -2.05 | 0.04 | -48.96 | <2e-16 |
| Sex - women | 0.39 | 0.02 | 22.61 | <2e-16 | 0.61 | 0.05 | 11.74 | <2e-16 |
| **Smooth terms** | **edf** | **edf.df** | **F** | **P** | **edf** | **edf.df** | **F** | **P** |
| s(Time) | 10.29 | 12.29 | 12.72 | <2e-16 | 12.29 | 14.25 | 12.97 | <2e-16 |
| s(Time):sex - women | 3.16 | 3.92 | 2.65 | 0.52 | 5.35 | 6.6 | 2.61 | 0.21 |
| s(Subject.id) | - | - | - | - | 3218.60 | 5000 | 96.71 | <2e-16 |
| s(Family.id) | - | - | - | - | 628.54 | 4609 | 13.63 | 1.00 |
| s(Time, Subject.id) | - | - | - | - | 1872.01 | 4998 | 24.57 | 4.51e-11 |
| s(Time, Family.id) | - | - | - | - | 0.03 | 4608 | 0.00 | 0.79 |
| **Suicidal ideation** | **Estimate** | **SE** | **Z** | **P** | **Estimate** | **SE** | **Z** | **P** |
| (Intercept) | -4.84 | 0.07 | -74.22 | <2e-16 | -5.65 | 0.10 | -53.82 | <2e-16 |
| Sex - women | -0.10 | 0.08 | -1.17 | 0.24 | -0.01 | 0.13 | -0.10 | 0.92 |
| **Smooth terms** | **edf** | **edf.df** | **Chi.sq** | **P** | **edf** | **edf.df** | **Chi.sq** | **P** |
| s(Time) | 1.00 | 1.00 | 6.93 | 0.17 | 1.00 | 1.00 | 0.32 | 1.00 |
| s(Time):sex - women | 1.00 | 1.00 | 0.08 | 1.00 | 1.00 | 1.00 | 0.37 | 1.00 |
| s(Subject.id) | - | - | - | - | 665.17 | 5000 | 3364.39 | <2e-16 |
| s(Family.id) | - | - | - | - | 0.02 | 4609 | 0.02 | 0.54 |
| s(Time, Subject.id) | - | - | - | - | 177.82 | 4998 | 679.27 | 5.21e-11 |
| s(Time, Family.id) | - | - | - | - | 0.02 | 4608 | 0.02 | 0.50 |
| **GAD** | **Estimate** | **SE** | **Z** | **P** | **Estimate** | **SE** | **Z** | **P** |
| (Intercept) | -4.10 | 0.05 | -89.58 | <2e-16 | -5.15 | 0.09 | -58.4 | <2e-16 |
| Sex - women | 0.48 | 0.05 | 8.78 | <2e-16 | 0.60 | 0.11 | 5.63 | <2e-16 |
| **Smooth terms** | **edf** | **edf.df** | **Chi.sq** | **P** | **edf** | **edf.df** | **Chi.sq** | **P** |
| s(Time) | 1.92 | 2.35 | 9.74 | 0.30 | 1.48 | 1.75 | 0.34 | 1.00 |
| s(Time):sex - women | 1.62 | 1.95 | 2.74 | 1.00 | 1.86 | 2.27 | 2.82 | 1.00 |
| s(Subject.id) | - | - | - | - | 1451.65 | 5000 | 7925.70 | <2e-16 |
| s(Family.id) | - | - | - | - | 0.04 | 4609 | 0.04 | 0.53 |

| | | | | | | | | |
|---|---|---|---|---|---|---|---|---|
| s(Time, Subject.id) | - | - | - | - | 295.05 | 4998 | 1417 | <2e-16 |
| s(Time, Family.id) | - | - | - | - | 0.03 | 4608 | 0.03 | 0.56 |
| **GAD score** | **Estimate** | **SE** | **t** | **P** | **Estimate** | **SE** | **t** | **P** |
| (Intercept) | -0.76 | 0.01 | -57.36 | <2e-16 | -1.94 | 0.04 | -44.87 | <2e-16 |
| Sex - women | 0.42 | 0.02 | 25.61 | <2e-16 | 0.67 | 0.05 | 12.53 | <2e-16 |
| **Smooth terms** | **edf** | **edf.df** | **F** | **P** | **edf** | **edf.df** | **F** | **P** |
| s(Time) | 10.08 | 12.07 | 17.26 | <2e-16 | 12.65 | 14.63 | 27.01 | <2e-16 |
| s(Time):sex - women | 1.00 | 1.00 | 6.90 | 0.17 | 1.25 | 1.45 | 0.14 | 1.00 |
| s(Subject.id) | - | - | - | - | 3567.51 | 5000 | 104.92 | <2e-16 |
| s(Family.id) | - | - | - | - | 437.01 | 4609 | 8.45 | 1 |
| s(Time, Subject.id) | - | - | - | - | 1984.12 | 4998 | 33.09 | 3.44e-09 |
| s(Time, Family.id) | - | - | - | - | 1.36 | 4608 | 0.00 | 1 |

**Table S10. GAM estimates of MD and GAD outcome trajectories by time and lifetime history of MD/GAD and without correction for within-subject and -family dependencies.** Shown are the statistical output of the GAMs for each of the five outcome variables for analyses performed in a random sample of 5,000 study participants selected from the main analyses sample of 36,106 individuals. The prevalence was modeled using an ordered factor difference smooth of MD/GAD lifetime history (LT) across time (in days) as predictor without (left columns) and with (right columns) correction for dependencies within subject and family. Shown are the estimate and standard error (SE) of the intercept and the fitted smooths for each term in the model. The effective degree of freedom (edf) is a proxy for the degree of non-linearity of the smooth. An edf of 1.0 indicates a linear relationship, while an edf > 1.0 indicates non-linearity. P-values of the smooth terms of interest are adjusted by Bonferroni correction. Edf.df = reference degree of freedom, Z = z-score, Chi.sq= chi-square, P = p-value, t = t-value, F = F-value.

| | Sub sample without correction for subject and family dependencies (N=5K) | | | | Sub sample with correction for subject and family dependencies (N=5K) | | | |
|---|---|---|---|---|---|---|---|---|
| **MDD** | **Estimate** | **SE** | **Z** | **P** | **Estimate** | **SE** | **Z** | **P** |
| (Intercept) | -4.85 | 0.05 | -105.84 | <2e-16 | -5.52 | 0.07 | -82.62 | <2e-16 |
| MDD_LT - yes | 1.97 | 0.06 | 34.45 | <2e-16 | 1.53 | 0.09 | 16.84 | <2e-16 |
| **Smooth terms** | **edf** | **edf.df** | **Chi.sq** | **P** | **edf** | **edf.df** | **Chi.sq** | **P** |
| s(Time) | 4.96 | 6.15 | 59.28 | <2e-16 | 5.48 | 6.78 | 53.49 | 5.66e-08 |
| s(Time):MDD_LT-yes | 1.35 | 1.61 | 6.94 | 0.99 | 1.00 | 1.00 | 1.36 | 1.00 |
| s(Subject.id) | - | - | - | - | 982.34 | 5000 | 5593.33 | <2e-16 |
| s(Family.id) | - | - | - | - | 179.90 | 4609 | 473.01 | 0.10 |
| s(Time, Subject.id) | - | - | - | - | 198.57 | 4999 | 749.49 | 2.02e-14 |
| s(Time, Family.id) | - | - | - | - | 0.08 | 4608 | 0.08 | 0.50 |
| **MDD score** | **Estimate** | **SE** | **t** | **P** | **Estimate** | **SE** | **t** | **P** |
| (Intercept) | -1.03 | 0.01 | -106.98 | <2e-16 | -1.73 | 0.03 | -67.72 | <2e-16 |
| MDD_LT - yes | 1.04 | 0.02 | 59.75 | <2e-16 | 0.31 | 0.02 | 15.06 | <2e-16 |
| **Smooth terms** | **edf** | **edf.df** | **F** | **P** | **edf** | **edf.df** | **F** | **P** |
| s(Time) | 10.59 | 12.62 | 18.08 | <2e-16 | 12.44 | 14.42 | 17.89 | <2e-16 |
| s(Time):MDD_LT-yes | 1.74 | 2.16 | 0.50 | 1.00 | 4.01 | 4.98 | 2.41 | 0.74 |
| s(Subject.id) | - | - | - | - | 3169.21 | 5000 | 78.58 | 8.98e-09 |
| s(Family.id) | - | - | - | - | 670.95 | 4609 | 12.91 | 1.00 |
| s(Time, Subject.id) | - | - | - | - | 1867.88 | 4999 | 23.87 | 1.85e-11 |
| s(Time, Family.id) | - | - | - | - | 0.04 | 4608 | 0.00 | 0.81 |
| **Suicidal ideation** | **Estimate** | **SE** | **Z** | **P** | **Estimate** | **SE** | **Z** | **P** |
| (Intercept) | -5.68 | 0.07 | -82.59 | <2e-16 | -6.12 | 0.09 | -70.42 | <2e-16 |
| MDD_LT - yes | 1.92 | 0.09 | 22.06 | <2e-16 | 1.41 | 0.12 | 11.45 | <2e-16 |
| **Smooth terms** | **edf** | **edf.df** | **Chi.sq** | **P** | **edf** | **edf.df** | **Chi.sq** | **P** |
| s(Time) | 1.00 | 1.00 | 14.66 | 2.58e-03 | 1 | 1 | 0.84 | 1.00 |
| s(Time):MDD_LT-yes | 1.00 | 1.00 | 1.87 | 1.00 | 1 | 1 | 1.96 | 1.00 |
| s(Subject.id) | - | - | - | - | 564.34 | 5000 | 2482.57 | <2e-16 |
| s(Family.id) | - | - | - | - | 0.01 | 4609 | 0.01 | 0.53 |
| s(Time, Subject.id) | - | - | - | - | 193.61 | 4999 | 742.95 | 5.09e-14 |
| s(Time, Family.id) | - | - | - | - | 0.02 | 4608 | 0.02 | 0.50 |
| **GAD** | **Estimate** | **SE** | **Z** | **P** | **Estimate** | **SE** | **Z** | **P** |
| (Intercept) | -4.14 | 0.03 | -135.87 | <2e-16 | -4.89 | 0.05 | -96.17 | <2e-16 |
| GAD_LT - yes | 1.96 | 0.05 | 36.87 | <2e-16 | 1.37 | 0.10 | 13.76 | <2e-16 |
| **Smooth terms** | **edf** | **edf.df** | **Chi.sq** | **P** | **edf** | **edf.df** | **Chi.sq** | **P** |
| s(Time) | 2.33 | 2.89 | 13.06 | 0.12 | 2.25 | 2.78 | 5.07 | 1.00 |
| s(Time):GAD_LT-yes | 1.00 | 1.00 | 0.33 | 1.00 | 1.00 | 1.00 | 0.79 | 1.00 |
| s(Subject.id) | - | - | - | - | 1379.07 | 5000 | 6899.30 | <2e-16 |

| | | | | | | | | |
|---|---|---|---|---|---|---|---|---|
| s(Family.id) | - | - | - | - | 0.02 | 4609 | 0.02 | 0.56 |
| s(Time, Subject.id) | - | - | - | - | 292.2 | 4999 | 1285.02 | <2e-16 |
| s(Time, Family.id) | - | - | - | - | 0.01 | 4608 | 0.01 | 0.57 |
| **GAD score** | **Estimate** | **SE** | **t** | **P** | **Estimate** | **SE** | **t** | **P** |
| (Intercept) | -0.64 | 0.01 | -75.77 | <2e-16 | -1.53 | 0.03 | -58.39 | <2e-16 |
| GAD_LT - yes | 1.14 | 0.03 | 45.04 | <2e-16 | 0.25 | 0.02 | 10.99 | <2e-16 |
| **Smooth terms** | **edf** | **edf.df** | **F** | **P** | **edf** | **edf.df** | **F** | **P** |
| s(Time) | 10.46 | 12.48 | 18.18 | <2e-16 | 12.71 | 14.68 | 30.43 | <2e-16 |
| s(Time):GAD_LT-yes | 2.13 | 2.63 | 0.75 | 1.00 | 4.95 | 6.13 | 4.44 | 3.02e-03 |
| s(Subject.id) | - | - | - | - | 3609.81 | 5000 | 93.03 | <2e-16 |
| s(Family.id) | - | - | - | - | 411.53 | 4609 | 7.06 | 1.00 |
| s(Time, Subject.id) | - | - | - | - | 1980.36 | 4999 | 32.65 | 5.84e-09 |
| s(Time, Family.id) | - | - | - | - | 1.02 | 4608 | 0.00 | 1.00 |

**Table S11. GAM estimates of MD and GAD outcome trajectories with and without correction for within-subject and -family dependencies in the youngest 5K participants.** Shown are the statistical output of the GAMs for each of the five outcome variables for analyses performed in the youngest 5,000 study participants selected from the main analyses sample of 36,106 individuals. The prevalence was modeled using a single smooth term of time (in days) as predictor without (left columns) and with (right columns) correction for dependencies within subject and family. Shown are the estimate and standard error (SE) of the intercept and the fitted smooth of the time variable with corresponding test statistics. The effective degree of freedom (edf) is a proxy for the degree of non-linearity of the smooth. An edf of 1.0 indicates a linear relationship, while an edf > 1.0 indicates non-linearity. P-values of the smooth terms of interest are adjusted by Bonferroni correction. Edf.df = reference degree of freedom, Z = z-score, Chi.sq= chi-square, P = p-value, t = t-value, F = F-value.

| | Sub sample without correction for subject and family dependencies (N= youngest 5K) | | | | Sub sample with correction for subject and family dependencies (N= youngest 5K) | | | |
|---|---|---|---|---|---|---|---|---|
| **MDD** | **Estimate** | **SE** | **Z** | **P** | **Estimate** | **SE** | **Z** | **P** |
| (Intercept) | -3.38 | 0.02 | -152.56 | <2e-16 | -4.41 | 0.05 | -92.36 | <2e-16 |
| **Smooth terms** | **edf** | **edf.df** | **Chi.sq** | **P** | **edf** | **edf.df** | **Chi.sq** | **P** |
| s(Time) | 5.43 | 6.73 | 74.56 | <2e-16 | 5.51 | 6.82 | 43.54 | 5.10e-06 |
| s(Subject.id) | - | - | - | - | 1080.56 | 5000 | 7063.6 | 3.17e-21 |
| s(Family.id) | - | - | - | - | 602.76 | 4501 | 3153.57 | 0.02 |
| s(Time, Subject.id) | - | - | - | - | 392.57 | 4999 | 2400.45 | 4.35e-25 |
| s(Time, Family.id) | - | - | - | - | 0.01 | 4500 | 0.01 | 0.60 |
| **MDD score** | **Estimate** | **SE** | **t** | **P** | **Estimate** | **SE** | **t** | **P** |
| (Intercept) | -0.34 | 0.01 | -41.28 | <2e-16 | -1.19 | 0.02 | -49.64 | <2e-16 |
| **Smooth terms** | **edf** | **edf.df** | **F** | **P** | **edf** | **edf.df** | **F** | **P** |
| s(Time) | 10.14 | 12.15 | 21.00 | <2e-16 | 12.40 | 14.42 | 30.18 | 2.50e-81 |
| s(Subject.id) | - | - | - | - | 3248.67 | 5000 | 117.83 | 7.95e-11 |
| s(Family.id) | - | - | - | - | 770.69 | 4501 | 25.53 | 1.00 |
| s(Time, Subject.id) | - | - | - | - | 2073.28 | 4999 | 33.76 | 9.73e-05 |
| s(Time, Family.id) | - | - | - | - | 126.57 | 4500 | 0.33 | 1.00 |
| **Suicidal ideation** | **Estimate** | **SE** | **Z** | **P** | **Estimate** | **SE** | **Z** | **P** |
| (Intercept) | -4.37 | 0.03 | -126.66 | <2e-16 | -5.16 | 0.06 | -90.12 | <2e-16 |
| **Smooth terms** | **edf** | **edf.df** | **Chi.sq** | **P** | **edf** | **edf.df** | **Chi.sq** | **P** |
| s(Time) | 1.00 | 1.00 | 17.63 | 5.50e-04 | 1.00 | 1.00 | 0.08 | 1.00 |
| s(Subject.id) | - | - | - | - | 451.19 | 5000 | 2073.37 | 2.04e-14 |
| s(Family.id) | - | - | - | - | 441.17 | 4501 | 2019.64 | 1.56e-13 |
| s(Time, Subject.id) | - | - | - | - | 210.28 | 4999 | 819.62 | 6.35e-12 |
| s(Time, Family.id) | - | - | - | - | 0.03 | 4500 | 0.03 | 0.51 |
| **GAD** | **Estimate** | **SE** | **Z** | **P** | **Estimate** | **SE** | **Z** | **P** |
| (Intercept) | -3.19 | 0.02 | -158 | <2e-16 | -4.12 | 0.04 | -94.82 | <2e-16 |
| **Smooth terms** | **edf** | **edf.df** | **Chi.sq** | **P** | **edf** | **edf.df** | **Chi.sq** | **P** |
| s(Time) | 4.20 | 5.23 | 28.84 | 6.92e-04 | 4.28 | 5.31 | 21.38 | 1.92e-02 |
| s(Subject.id) | - | - | - | - | 1304.61 | 5000 | 9009.46 | 7.94e-44 |
| s(Family.id) | - | - | - | - | 576.66 | 4501 | 2872.96 | 0.02 |
| s(Time, Subject.id) | - | - | - | - | 129.06 | 4999 | 357.58 | 2.35e-08 |
| s(Time, Family.id) | - | - | - | - | 298.66 | 4500 | 1490.15 | 5.48e-17 |
| **GAD score** | **Estimate** | **SE** | **t** | **P** | **Estimate** | **SE** | **t** | **P** |
| (Intercept) | -0.01 | 0.01 | -1.79 | 0.074 | -0.80 | 0.02 | -34.39 | <2e-16 |
| **Smooth terms** | **edf** | **edf.df** | **F** | **P** | **edf** | **edf.df** | **F** | **P** |
| s(Time) | 10.89 | 12.94 | 24.09 | <2e-16 | 14.50 | 16.39 | 42.10 | 3.32e-133 |
| s(Subject.id) | - | - | - | - | 3341.76 | 5000 | 127.72 | 0.02 |

| | | | | | | | | |
|---|---|---|---|---|---|---|---|---|
| s(Family.id) | - | - | - | - | 886.26 | 4501 | 34.90 | 1.00 |
| s(Time, Subject.id) | - | - | - | - | 2391.08 | 4999 | 45.37 | 1.95e-25 |
| s(Time, Family.id) | - | - | - | - | 0.91 | 4500 | 0.00 | 1.00 |

**Table S12. GAM estimates of MD and GAD outcome trajectories by time and age and without correction for within-subject and -family dependencies in the youngest 5K participants.** Shown are the statistical output of the GAMs for each of the five outcome variables for analyses performed in the youngest 5,000 study participants selected from the main analyses sample of 36,106 individuals. The prevalence was modeled using a single smooth term of time (in days), a single smooth term of age (in years), and a tensor product interaction smooth term between time and age as predictors without (left columns) and with (right columns) correction for dependencies within subject and family. Shown are the estimate and standard error (SE) of the intercept and the fitted smooths for each term in the model. The effective degree of freedom (edf) is a proxy for the degree of non-linearity of the smooth. An edf of 1.0 indicates a linear relationship, while an edf > 1.0 indicates non-linearity. P-values of the smooth terms of interest are adjusted by Bonferroni correction. Edf.df = reference degree of freedom, Z = z-score, Chi.sq= chi-square, P = p-value, t = t-value, F = F-value.

| | Sub sample without correction for subject and family dependencies (N= youngest 5K) | | | | Sub sample with correction for subject and family dependencies (N= youngest 5K) | | | |
|---|---|---|---|---|---|---|---|---|
| **MDD** | **Estimate** | **SE** | **Z** | **P** | **Estimate** | **SE** | **Z** | **P** |
| (Intercept) | -3.22 | 0.03 | -122.91 | <2e-16 | -4.23 | 0.06 | -72.18 | <2e-16 |
| **Smooth terms** | **edf** | **edf.df** | **Chi.sq** | **P** | **edf** | **edf.df** | **Chi.sq** | **P** |
| s(Time) | 5.39 | 6.68 | 73.89 | <2e-16 | 5.50 | 6.8 | 47.14 | 5.15e-08 |
| s(Age) | 17.20 | 18.54 | 279.34 | <2e-16 | 2.25 | 2.53 | 24.98 | 1.20e-05 |
| ti(Time, Age) | 2.58 | 3.24 | 6.23 | 1.00 | 3.59 | 4.87 | 9.21 | 1.00 |
| s(Subject.id) | - | - | - | - | 1048.88 | 5000 | 6759.44 | 2.00e-19 |
| s(Family.id) | - | - | - | - | 624.27 | 4501 | 3330.17 | 4.80e-03 |
| s(Time, Subject.id) | - | - | - | - | 385.5 | 4999 | 2324.37 | 3.61e-25 |
| s(Time, Family.id) | - | - | - | - | 0.02 | 4500 | 0.02 | 0.57 |
| **MDD score** | **Estimate** | **SE** | **t** | **P** | **Estimate** | **SE** | **t** | **P** |
| (Intercept) | -0.24 | 0.01 | -22.91 | <2e-16 | -4.17 | 0.05 | -80.99 | <2e-16 |
| **Smooth terms** | **edf** | **edf.df** | **F** | **P** | **edf** | **edf.df** | **F** | **P** |
| s(Time) | 10.27 | 12.29 | 20.22 | <2e-16 | 4.82 | 5.97 | 7.48 | 5.48e-08 |
| s(Age) | 17.54 | 18.67 | 19.8 | <2e-16 | 2.25 | 2.55 | 10.22 | 7.37e-06 |
| ti(Time, Age) | 1.00 | 1.01 | 0.02 | 1.00 | 2.67 | 3.24 | 2.44 | 1.00 |
| s(Subject.id) | - | - | - | - | 999.19 | 5000 | 1.61 | 0.11 |
| s(Family.id) | - | - | - | - | 619.97 | 4501 | 0.92 | 1.00 |
| s(Time, Subject.id) | - | - | - | - | 136.02 | 4999 | 0.12 | 0.77 |
| s(Time, Family.id) | - | - | - | - | 0.01 | 4500 | 0.00 | 0.56 |
| **Suicidal ideation** | **Estimate** | **SE** | **Z** | **P** | **Estimate** | **SE** | **Z** | **P** |
| (Intercept) | -4.20 | 0.04 | -104.11 | <2e-16 | -4.99 | 0.07 | -76 | <2e-16 |
| **Smooth terms** | **edf** | **edf.df** | **Chi.sq** | **P** | **edf** | **edf.df** | **Chi.sq** | **P** |
| s(Time) | 1.00 | 1.00 | 31.24 | <2e-16 | 1.00 | 1.00 | 4.04 | 0.04 |
| s(Age) | 12.33 | 14.75 | 138.59 | <2e-16 | 1.00 | 1.00 | 24.61 | 7.03e-07 |
| ti(Time, Age) | 1.00 | 1.00 | 17.91 | 4.74e-04 | 1.00 | 1.00 | 18.03 | 4.38-04 |
| s(Subject.id) | - | - | - | - | 440.28 | 5000 | 1983.97 | 1.06e-13 |
| s(Family.id) | - | - | - | - | 454.67 | 4501 | 2096.8 | 2.63e-14 |
| s(Time, Subject.id) | - | - | - | - | 167.64 | 4999 | 565.44 | 2.07e-08 |
| s(Time, Family.id) | - | - | - | - | 25.44 | 4500 | 36.15 | 0.03 |
| **GAD** | **Estimate** | **SE** | **Z** | **P** | **Estimate** | **SE** | **Z** | **P** |
| (Intercept) | -3.13 | 0.03 | -124.71 | <2e-16 | -4.04 | 0.05 | -75.43 | <2e-16 |
| **Smooth terms** | **edf** | **edf.df** | **Chi.sq** | **P** | **edf** | **edf.df** | **Chi.sq** | **P** |
| s(Time) | 4.21 | 5.23 | 28.75 | 3.55e-05 | 4.29 | 5.32 | 18.40 | 3.38e-03 |
| s(Age) | 3.03 | 3.79 | 20.97 | 3.11e-04 | 1.00 | 1.01 | 6.85 | 9.04e-04 |
| ti(Time, Age) | 7.88 | 11.52 | 16.31 | 1.00 | 9.80 | 14.22 | 19.66 | 1.00 |
| s(Subject.id) | - | - | - | - | 1279.81 | 5000 | 8793.19 | 7.62e-41 |

| | | | | | | | | |
|---|---|---|---|---|---|---|---|---|
| s(Family.id) | - | - | - | - | 598.49 | 4501 | 3039.99 | 8.68e-03 |
| s(Time, Subject.id) | - | - | - | - | 110.29 | 4999 | 278.44 | 9.14e-08 |
| s(Time, Family.id) | - | - | - | - | 312.79 | 4500 | 1612.3 | 1.03e-18 |
| **GAD score** | **Estimate** | **SE** | **t** | **P** | **Estimate** | **SE** | **t** | **P** |
| (Intercept) | 0.05 | 0.01 | 5.18 | 2.17e-07 | -0.7 | 0.03 | -23.98 | <2e-16 |
| **Smooth terms** | **edf** | **edf.df** | **F** | **P** | **edf** | **edf.df** | **F** | **P** |
| s(Time) | 10.99 | 13.05 | 22.43 | <2e-16 | 14.46 | 16.36 | 26.44 | 6.21e-81 |
| s(Age) | 17.19 | 18.51 | 11.94 | <2e-16 | 1.00 | 1.00 | 32.94 | 9.44e-09 |
| ti(Time, Age) | 4.61 | 6.75 | 0.67 | 1.00 | 25.83 | 37.81 | 1.62 | 0.18 |
| s(Subject.id) | - | - | - | - | 3347.06 | 5000 | 125.48 | 6.89e-03 |
| s(Family.id) | - | - | - | - | 876.19 | 4501 | 33.45 | 1.00 |
| s(Time, Subject.id) | - | - | - | - | 2381.56 | 4999 | 43.39 | 2.24e-23 |
| s(Time, Family.id) | - | - | - | - | 5.19 | 4500 | 0.00 | 1.00 |

**Table S13. GAM estimates of MD and GAD outcome trajectories by time and sex and without correction for within-subject and -family dependencies in youngest 5K participants.** Shown are the statistical output of the GAMs for each of the five outcome variables for analyses performed in the youngest 5,000 study participants selected from the main analyses sample of 36,106 individuals. The prevalence was modeled using an ordered factor difference smooth of sex across time (in days) as predictor without (left columns) and with (right columns) correction for dependencies within subject and family. Shown are the estimate and standard error (SE) of the intercept and the fitted smooths for each term in the model. The effective degree of freedom (edf) is a proxy for the degree of non-linearity of the smooth. An edf of 1.0 indicates a linear relationship, while an edf > 1.0 indicates non-linearity. P-values of the smooth terms of interest are adjusted by Bonferroni correction. Edf.df = reference degree of freedom, Z = z-score, Chi.sq= chi-square, P = p-value, t = t-value, F = F-value.

| | **Sub sample without correction for subject and family dependencies (N= youngest 5K)** | | | | **Sub sample with correction for subject and family dependencies (N= youngest 5K)** | | | |
|---|---|---|---|---|---|---|---|---|
| **MDD** | **Estimate** | **SE** | **Z** | **P** | **Estimate** | **SE** | **Z** | **P** |
| (Intercept) | -3.57 | 0.04 | -83.70 | <2e-16 | -4.60 | 0.09 | -50.76 | <2e-16 |
| Sex - women | 0.25 | 0.05 | 5.14 | 2.73e-07 | 0.28 | 0.11 | 2.62 | 8.84e-03 |
| **Smooth terms** | **edf** | **edf.df** | **Chi.sq** | **P** | **edf** | **edf.df** | **Chi.sq** | **P** |
| s(Time) | 5.43 | 6.73 | 59.60 | <2e-16 | 5.51 | 6.82 | 43.48 | 2.63e-07 |
| s(Time):sex - women | 1.00 | 1.00 | 2.52 | 1.00 | 1.00 | 1.00 | 0.58 | 1.00 |
| s(Subject.id) | - | - | - | - | 1107.73 | 5000 | 7289.81 | 3.39e-24 |
| s(Family.id) | - | - | - | - | 571.71 | 4501 | 2907.70 | 0.03 |
| s(Time, Subject.id) | - | - | - | - | 391.22 | 4998 | 2376.81 | 6.99e-25 |
| s(Time, Family.id) | - | - | - | - | 0.01 | 4500 | 0.01 | 0.60 |
| **MDD score** | **Estimate** | **SE** | **t** | **P** | **Estimate** | **SE** | **t** | **P** |
| (Intercept) | -0.58 | 0.02 | -37.96 | <2e-16 | -1.55 | 0.04 | -35.11 | <2e-16 |
| Sex - women | 0.33 | 0.02 | 18.44 | <2e-16 | 0.51 | 0.05 | 9.90 | <2e-16 |
| **Smooth terms** | **edf** | **edf.df** | **F** | **P** | **edf** | **edf.df** | **F** | **P** |
| s(Time) | 10.13 | 12.12 | 14.03 | <2e-16 | 12.33 | 14.32 | 17.63 | 4.41e-45 |
| s(Time):sex - women | 3.73 | 4.62 | 3.36 | 0.12 | 4.86 | 5.99 | 4.68 | 1.84e-03 |
| s(Subject.id) | - | - | - | - | 3288.22 | 5000 | 115.53 | 2.99e-14 |
| s(Family.id) | - | - | - | - | 713.73 | 4501 | 21.87 | 1.00 |
| s(Time, Subject.id) | - | - | - | - | 2069.7 | 4998 | 32.9 | 1.12e-04 |
| s(Time, Family.id) | - | - | - | - | 128.45 | 4500 | 0.34 | 1.00 |
| **Suicidal ideation** | **Estimate** | **SE** | **Z** | **P** | **Estimate** | **SE** | **Z** | **P** |
| (Intercept) | -4.48 | 0.07 | -67.8 | <2e-16 | -5.21 | 0.11 | -49.19 | <2e-16 |
| Sex - women | 0.14 | 0.08 | 1.86 | 0.06 | 0.07 | 0.12 | 0.55 | 0.58 |
| **Smooth terms** | **edf** | **edf.df** | **Chi.sq** | **P** | **edf** | **edf.df** | **Chi.sq** | **P** |
| s(Time) | 1.00 | 1.00 | 8.72 | 3.16e-03 | 1.00 | 1.00 | 0.14 | 0.71 |
| s(Time):sex - women | 1.00 | 1.00 | 0.81 | 1.00 | 1.00 | 1.00 | 0.38 | 1.00 |
| s(Subject.id) | - | - | - | - | 461.8 | 5000 | 2144.90 | 2.31e-15 |
| s(Family.id) | - | - | - | - | 429.82 | 4501 | 1937.38 | 4.81e-13 |
| s(Time, Subject.id) | - | - | - | - | 210.37 | 4998 | 818.89 | 3.93e-12 |
| s(Time, Family.id) | - | - | - | - | 0.05 | 4500 | 0.05 | 0.51 |
| **GAD** | **Estimate** | **SE** | **Z** | **P** | **Estimate** | **SE** | **Z** | **P** |
| (Intercept) | -3.35 | 0.04 | -86.56 | <2e-16 | -4.35 | 0.08 | -52.47 | <2e-16 |
| Sex - women | 0.22 | 0.04 | 4.89 | 1.02e-06 | 0.31 | 0.1 | 3.22 | 1.20e-03 |
| **Smooth terms** | **edf** | **edf.df** | **Chi.sq** | **P** | **edf** | **edf.df** | **Chi.sq** | **P** |
| s(Time) | 4.20 | 5.22 | 33.28 | 4.33e-06 | 4.28 | 5.31 | 21.76 | 8.22e-04 |
| s(Time):sex - women | 1.00 | 1.01 | 6.21 | 0.26 | 1.00 | 1.00 | 5.36 | 0.41 |
| s(Subject.id) | - | - | - | - | 1351.71 | 5000 | 9351.96 | 1.62e-52 |
| s(Family.id) | - | - | - | - | 523.4 | 4501 | 2461.65 | 0.07 |

| | | | | | | | | |
|---|---|---|---|---|---|---|---|---|
| s(Time, Subject.id) | - | - | - | - | 113.4 | 4998 | 290.63 | 9.83e-08 |
| s(Time, Family.id) | - | - | - | - | 310.8 | 4500 | 1588.18 | 3.09e-18 |

| **GAD score** | **Estimate** | **SE** | **t** | **P** | **Estimate** | **SE** | **t** | **P** |
|---|---|---|---|---|---|---|---|---|
| (Intercept) | -0.23 | 0.01 | -17.30 | <2e-16 | -1.17 | 0.04 | -27.47 | <2e-16 |
| Sex - women | 0.30 | 0.02 | 19.19 | <2e-16 | 0.52 | 0.05 | 10.46 | <2e-16 |
| **Smooth terms** | **edf** | **edf.df** | **F** | **P** | **edf** | **edf.df** | **F** | **P** |
| s(Time) | 10.81 | 12.86 | 24.81 | <2e-16 | 14.45 | 16.34 | 20.71 | 1.54e-61 |
| s(Time):sex - women | 1.00 | 1.00 | 7.35 | 0.13 | 3.96 | 4.91 | 1.88 | 1.00 |
| s(Subject.id) | - | - | - | - | 3352.9 | 5000 | 125.62 | 1.16e-03 |
| s(Family.id) | - | - | - | - | 857.55 | 4501 | 32.72 | 1.00 |
| s(Time, Subject.id) | - | - | - | - | 2390.3 | 4998 | 44.33 | 5.16e-26 |
| s(Time, Family.id) | - | - | - | - | 0.65 | 4500 | 0.00 | 1.00 |

**Table S14. GAM estimates of MD and GAD outcome trajectories by time and lifetime history of MD/GAD and without correction for within-subject and -family dependencies in youngest 5K participants.** Shown are the statistical output of the GAMs for each of the five outcome variables for analyses performed in the youngest 5,000 study participants selected from the main analyses sample of 36,106 individuals. The prevalence was modeled using an ordered factor difference smooth of MD/GAD lifetime history (LT) across time (in days) as predictor without (left columns) and with (right columns) correction for dependencies within subject and family. Shown are the estimate and standard error (SE) of the intercept and the fitted smooths for each term in the model. The effective degree of freedom (edf) is a proxy for the degree of non-linearity of the smooth. An edf of 1.0 indicates a linear relationship, while an edf > 1.0 indicates non-linearity. P-values of the smooth terms of interest are adjusted by Bonferroni correction. Edf.df = reference degree of freedom, Z = z-score, Chi.sq= chi-square, P = p-value, t = t-value, F = F-value.

| | Sub sample without correction for subject and family dependencies (N= youngest 5K) | | | | Sub sample with correction for subject and family dependencies (N= youngest 5K) | | | |
|---|---|---|---|---|---|---|---|---|
| **MDD** | **Estimate** | **SE** | **Z** | **P** | **Estimate** | **SE** | **Z** | **P** |
| (Intercept) | -4.16 | 0.04 | -113.2 | <2e-16 | -4.87 | 0.06 | -82.7 | <2e-16 |
| MDD_LT - yes | 1.68 | 0.05 | 37.26 | <2e-16 | 1.26 | 0.07 | 17.25 | <2e-16 |
| **Smooth terms** | **edf** | **edf.df** | **Chi.sq** | **P** | **edf** | **edf.df** | **Chi.sq** | **P** |
| s(Time) | 5.21 | 6.46 | 54.09 | <2e-16 | 5.36 | 6.63 | 41.2 | 6.49e-07 |
| s(Time):MDD_LT-yes | 1.00 | 1.00 | 1.18 | 1.00 | 1.00 | 1.00 | 0.80 | 1.00 |
| s(Subject.id) | - | - | - | - | 1010.6 | 5000 | 6151.53 | 1.39e-24 |
| s(Family.id) | - | - | - | - | 547.81 | 4501 | 2633.37 | 4.00e-03 |
| s(Time, Subject.id) | - | - | - | - | 379.76 | 4999 | 2089.65 | 2.3e-25 |
| s(Time, Family.id) | - | - | - | - | 0.01 | 4500 | 0.01 | 0.61 |
| **MDD score** | **Estimate** | **SE** | **t** | **P** | **Estimate** | **SE** | **t** | **P** |
| (Intercept) | -0.72 | 0.01 | -73.44 | <2e-16 | -1.28 | 0.02 | -53.47 | <2e-16 |
| MDD_LT - yes | 0.97 | 0.02 | 61.02 | <2e-16 | 0.33 | 0.02 | 19.63 | <2e-16 |
| **Smooth terms** | **edf** | **edf.df** | **F** | **P** | **edf** | **edf.df** | **F** | **P** |
| s(Time) | 10.33 | 12.35 | 18.92 | <2e-16 | 12.45 | 14.46 | 24.66 | 2.68e-66 |
| s(Time):MDD_LT-yes | 2.00 | 2.48 | 1.20 | 1.00 | 4.10 | 5.08 | 3.79 | 0.04 |
| s(Subject.id) | - | - | - | - | 3245.58 | 5000 | 95.01 | 2.74e-09 |
| s(Family.id) | - | - | - | - | 733.92 | 4501 | 19.58 | 1.00 |
| s(Time, Subject.id) | - | - | - | - | 2044.07 | 4999 | 30.35 | 4.49e-04 |
| s(Time, Family.id) | - | - | - | - | 136.52 | 4500 | 0.35 | 1.00 |
| **Suicidal ideation** | **Estimate** | **SE** | **Z** | **P** | **Estimate** | **SE** | **Z** | **P** |
| (Intercept) | -5.21 | 0.06 | -84.66 | <2e-16 | -5.74 | 0.08 | -69.5 | <2e-16 |
| MDD_LT - yes | 1.75 | 0.07 | 23.5 | <2e-16 | 1.40 | 0.11 | 13.27 | <2e-16 |
| **Smooth terms** | **edf** | **edf.df** | **Chi.sq** | **P** | **edf** | **edf.df** | **Chi.sq** | **P** |
| s(Time) | 1.00 | 1.00 | 5.23 | 0.02 | 1.00 | 1.00 | 0.06 | 0.80 |
| s(Time):MDD_LT-yes | 1.00 | 1.00 | 0.00 | 1.00 | 1.00 | 1.00 | 0.01 | 1.00 |
| s(Subject.id) | - | - | - | - | 341.9 | 5000 | 1369.14 | 7.35e-12 |
| s(Family.id) | - | - | - | - | 443.22 | 4501 | 2025.99 | 8.41e-19 |
| s(Time, Subject.id) | - | - | - | - | 215.68 | 4999 | 825.53 | 7.09e-14 |
| s(Time, Family.id) | - | - | - | - | 0.01 | 4500 | 0.01 | 0.54 |
| **GAD** | **Estimate** | **SE** | **Z** | **P** | **Estimate** | **SE** | **Z** | **P** |
| (Intercept) | -3.52 | 0.02 | -142.29 | <2e-16 | -4.28 | 0.05 | -94.1 | <2e-16 |
| GAD_LT - yes | 1.60 | 0.04 | 36.76 | <2e-16 | 1.15 | 0.08 | 15.04 | <2e-16 |
| **Smooth terms** | **edf** | **edf.df** | **Chi.sq** | **P** | **edf** | **edf.df** | **Chi.sq** | **P** |
| s(Time) | 4.44 | 5.51 | 34.94 | 3.31e-06 | 4.40 | 5.46 | 22.69 | 6.36e-04 |
| s(Time):GAD_LT-yes | 2.54 | 3.14 | 7.88 | 1.00 | 1.96 | 2.42 | 5.65 | 1.00 |
| s(Subject.id) | - | - | - | - | 1216.94 | 5000 | 7784.15 | 7.01e-43 |

| | | | | | | | | |
|---|---|---|---|---|---|---|---|---|
| s(Family.id) | - | - | - | - | 575.49 | 4501 | 2729.19 | 1.43e-03 |
| s(Time, Subject.id) | - | - | - | - | 168.56 | 4999 | 513.79 | 1.41e-10 |
| s(Time, Family.id) | - | - | - | - | 251.91 | 4500 | 1028.49 | 5.05e-14 |
| **GAD score** | **Estimate** | **SE** | **t** | **P** | **Estimate** | **SE** | **t** | **P** |
| (Intercept) | -0.16 | 0.01 | -20.94 | <2e-16 | -0.82 | 0.02 | -35.8 | <2e-16 |
| GAD_LT - yes | 0.91 | 0.02 | 44.92 | <2e-16 | 0.24 | 0.02 | 14.45 | <2e-16 |
| **Smooth terms** | **edf** | **edf.df** | **F** | **P** | **edf** | **edf.df** | **F** | **P** |
| s(Time) | 11.36 | 13.43 | 27.5 | <2e-16 | 14.51 | 16.4 | 42.38 | 1.52e-135 |
| s(Time):GAD_LT-yes | 3.78 | 4.7 | 2.87 | 0.36 | 5.34 | 6.61 | 6.05 | 2.18e-05 |
| s(Subject.id) | - | - | - | - | 3340.57 | 5000 | 112.87 | 0.11 |
| s(Family.id) | - | - | - | - | 874.28 | 4501 | 30.45 | 1.00 |
| s(Time, Subject.id) | - | - | - | - | 2381.12 | 4999 | 42.84 | 4.63e-24 |
| s(Time, Family.id) | - | - | - | - | 0.65 | 4500 | 0.00 | 1.00 |
