## Supplemental_Figures for "Longitudinal analyses of depression and anxiety highlight greater prevalence during COVID-19 lockdowns in the Dutch general population and a continuing increase in suicidal ideation in young adults"

### 1. Figure S1-3: Lifelines COVID-19 study questionnaire design and analysis plan

**Figure S1**. **Distribution of questionnaires over time and study participant response rate.** Shown are the assessment time points of the Lifelines COVID-19 study. Individual questionnaires are color-coded with the density distribution of the number of participants shown. The table below denotes the response rate as a percentage of the invited adult Lifelines cohort (N~140,000).

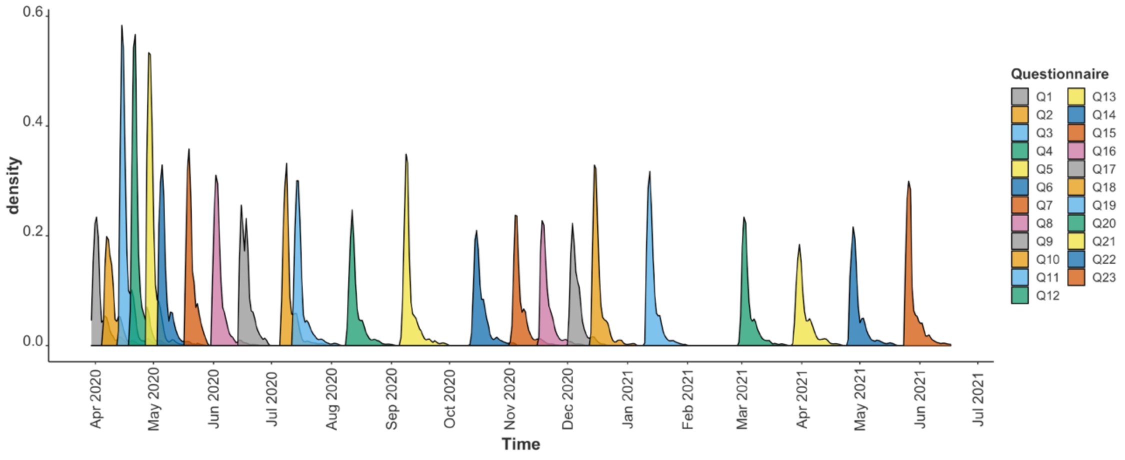

| | Q1 | Q2 | Q3 | Q4 | Q5 | Q6 | Q7 | Q8 | Q9 | Q10 | Q11 | Q12 | Q13 | Q14 | Q15 | Q16 | Q17 | Q18 | Q19 | Q20 | Q21 | Q22 | Q23 |
|---|---|---|---|---|---|---|---|---|---|---|---|---|---|---|---|---|---|---|---|---|---|---|---|
| **Response** | 53K | 51K | 50K | 47K | 45K | 43K | 43K | 38K | 35K | 33K | 35K | 36K | 35K | 34K | 34K | 32K | 31K | 32K | 34K | 30K | 29K | 29K | 29K |
| **Response rate** | 41% | 39% | 38% | 36% | 35% | 33% | 33% | 29% | 27% | 25% | 27% | 28% | 27% | 26% | 26% | 25% | 24% | 25% | 26% | 23% | 22% | 22% | 22% |

**Figure S2**. **An overview of the MINI depression and anxiety items that are included in the Lifelines COVID-19 questionnaires.** Shown are the twenty-three questionnaires in the columns and the nine depression and six anxiety items in the rows. The dates at the bottom of the graph show when the digital questionnaire was sent out to the study participants. For the first six questionnaires (Q1-Q6) symptoms were assessed over the past seven days, while for Q7 and beyond, symptoms were assessed over the past 14 days. Items from the full MINI depression domain were included in the questionnaire. For items of the MINI anxiety domain, O3C (feelings of fatigue), O3D (lack of concentration), and O3F (sleeping problems), were not included, as these were already included in the MINI MD items. The color-coding highlights if an item was included in a questionnaire (green) or not (grey). Missing data items were imputed. Questionnaire 12 was excluded from the study.

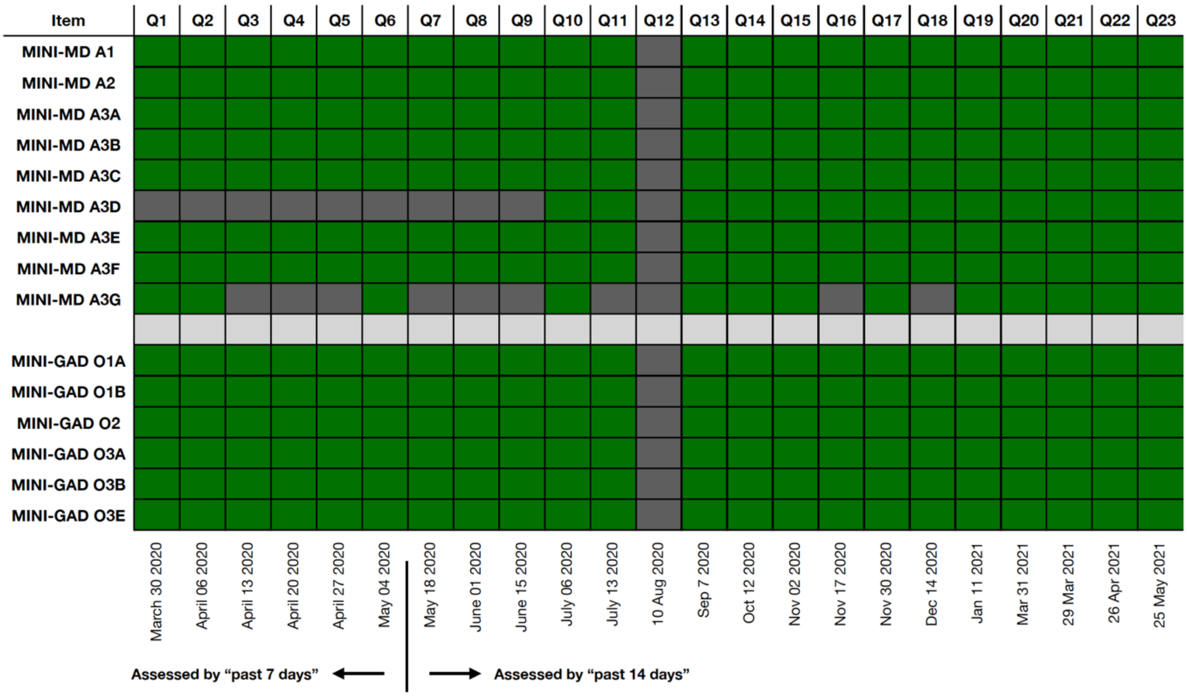

**Figure S3. An overview of the study analysis plan.** Shown is a simplified overview of the analytical strategy of the study. The distinct between the selected sub sample that was used for the primary analyses is shown together with information of the full cohort. The analyses workflow with subsequent steps, such as data imputation and different sensitivity analyses, are shown as well.

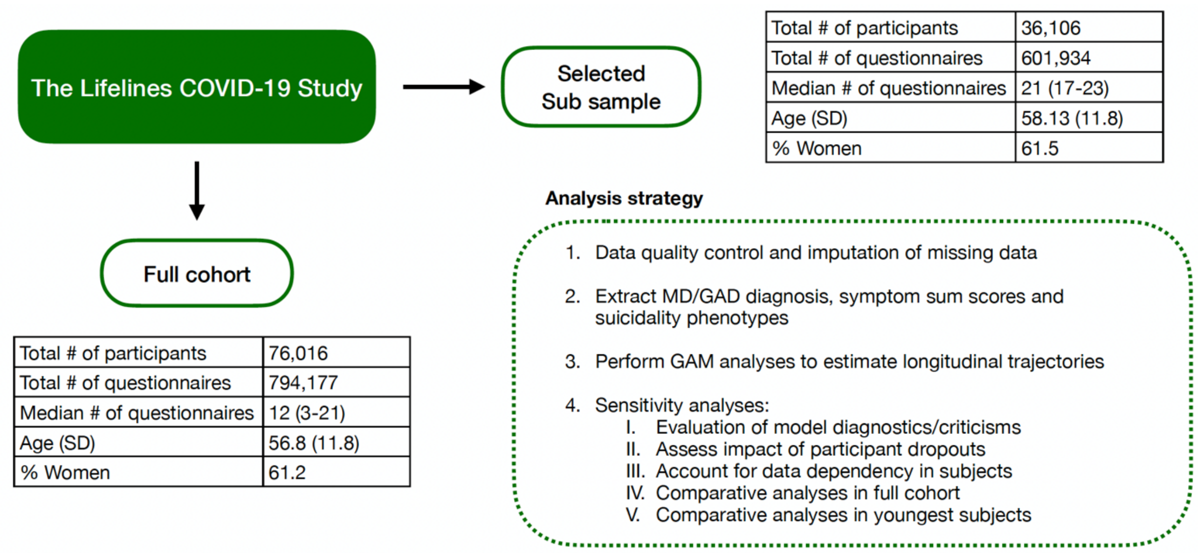

#### 2. Figure S4-8: longitudinal trajectories of phenotypic outcomes

**Figure S4. The longitudinal trajectory of depressive symptoms during the COVID-19 pandemic in the Lifelines cohort.** Shown are the fitted smooths with standard errors of mean depressive symptom score over time. Results of GAM analyses are shown for the following analyses: 1) main analysis sample of N=36,106 participants (left upper panel), 2) full cohort of N=76,016 participants (right upper panel), 3) a sub sample of 5,000 participants selected from analysis (1) without correction for random effects (left mid panel), 4) the same sub sample of 5,000 participants as (3) with correction for random effects (right mid panel), 5) a sub sample of the youngest 5,000 participants without correction for random effects (left lower panel), and 6) a sub sample of the youngest 5,000 participants with correction for random effects (right lower panel). The y-axis shows the mean symptom sum score and the x-axis the time in months. For the mixed-effect models, the random effects were removed before plotting and thus yielded lower prevalence estimates. The grey rectangles denoted the three lockdowns in the Netherlands.

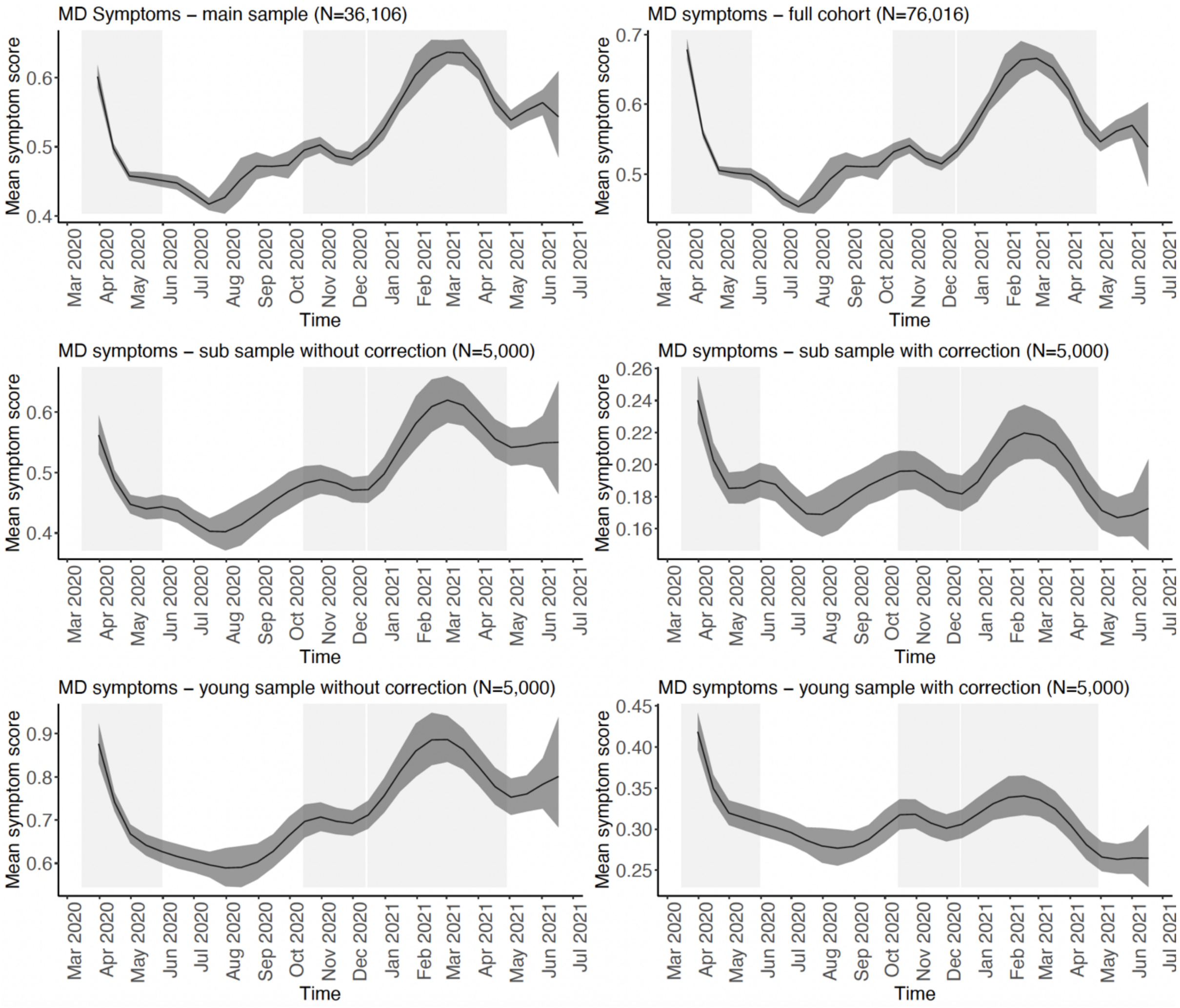

**Figure S5. The longitudinal trajectory of MDD during the COVID-19 pandemic in the Lifelines cohort.** Shown are the fitted smooths with standard errors of MDD prevalence over time. Results of GAM analyses are shown for the following analyses: 1) main analysis sample of N=36,106 participants (left upper panel), 2) full cohort of N=76,016 participants (right upper panel), 3) a sub sample of 5,000 participants selected from analysis (1) without correction for random effects (left mid panel), 4) the same sub sample of 5,000 participants as (3) with correction for random effects (right mid panel), 5) a sub sample of the youngest 5,000 participants without correction for random effects (left lower panel), and 6) a sub sample of the youngest 5,000 participants with correction for random effects (right lower panel). The y-axis shows the prevalence in % and the x-axis the time in months. For the mixed-effect models, the random effects were removed before plotting and thus yielded lower prevalence estimates. The grey rectangles denoted the three lockdowns in the Netherlands.

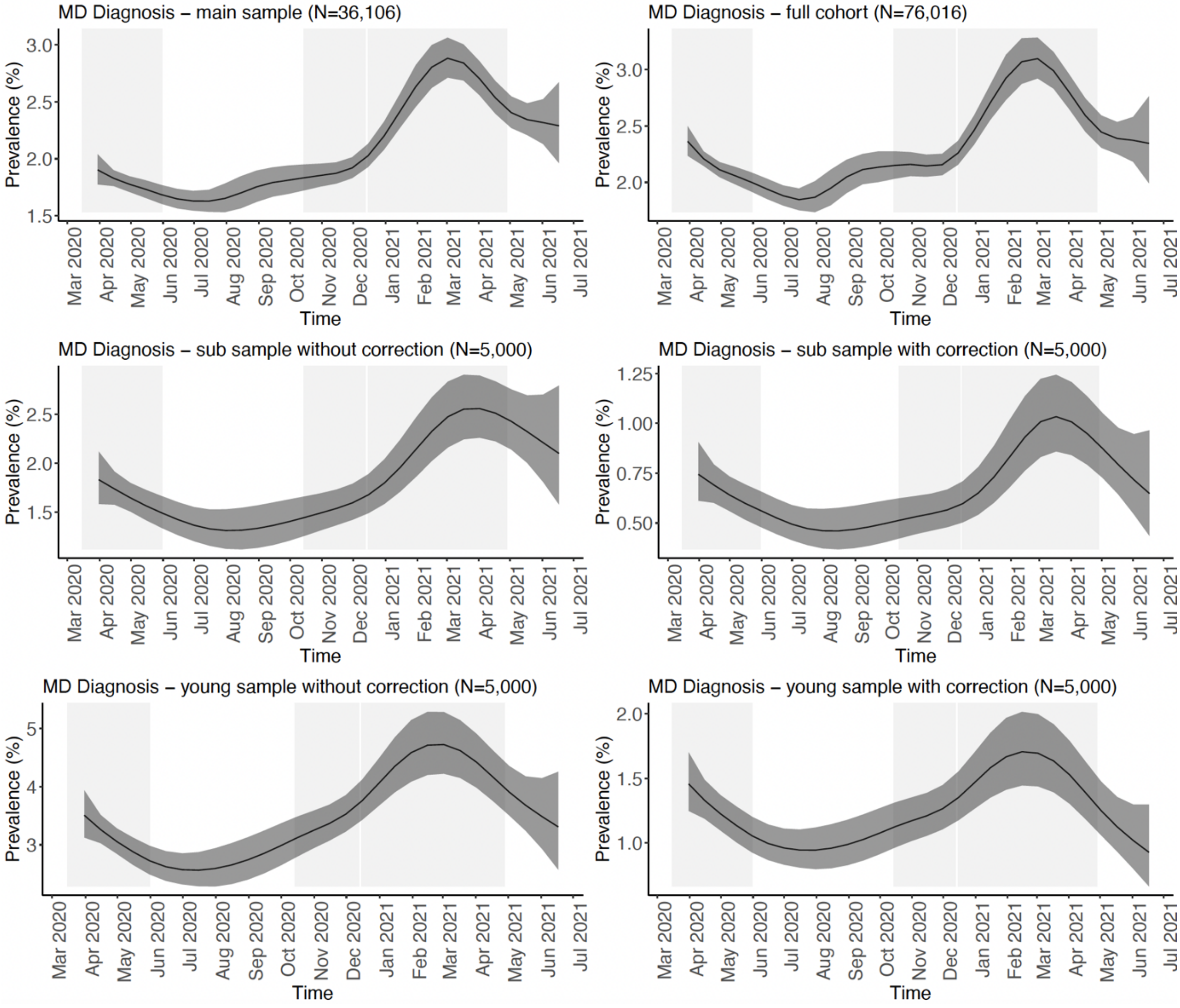

**Figure S6. The longitudinal trajectory of anxiety symptoms during the COVID-19 pandemic in the Lifelines cohort.** Shown are the fitted smooths with standard errors of mean anxiety symptom score over time. Results of GAM analyses are shown for the following analyses: 1) main analysis sample of N=36,106 participants (left upper panel), 2) full cohort of N=76,016 participants (right upper panel), 3) a sub sample of 5,000 participants selected from analysis (1) without correction for random effects (left mid panel), 4) the same sub sample of 5,000 participants as (3) with correction for random effects (right mid panel), 5) a sub sample of the youngest 5,000 participants without correction for random effects (left lower panel), and 6) a sub sample of the youngest 5,000 participants with correction for random effects (right lower panel). The y-axis shows the mean anxiety symptom sum score and the x-axis the time in months. For the mixed-effect models, the random effects were removed before plotting and thus yielded lower prevalence estimates. The grey rectangles denoted the three lockdowns in the Netherlands.

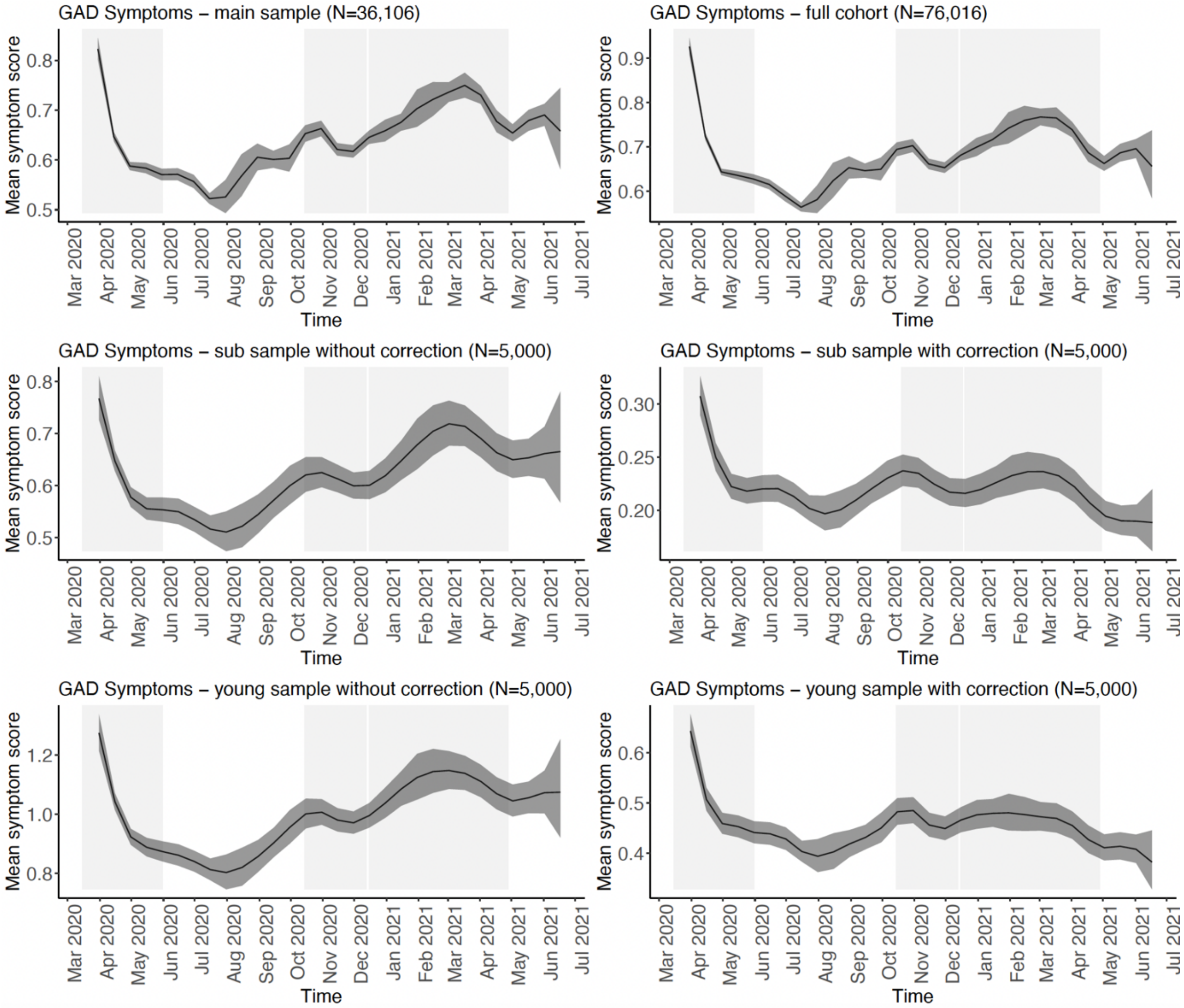

**Figure S7. The longitudinal trajectory of GAD during the COVID-19 pandemic in the Lifelines cohort.** Shown are the fitted smooths with standard errors of GAD prevalence over time. Results of GAM analyses are shown for the following analyses: 1) main analysis sample of N=36,106 participants (left upper panel), 2) full cohort of N=76,016 participants (right upper panel), 3) a sub sample of 5,000 participants selected from analysis (1) without correction for random effects (left mid panel), 4) the same sub sample of 5,000 participants as (3) with correction for random effects (right mid panel), 5) a sub sample of the youngest 5,000 participants without correction for random effects (left lower panel), and 6) a sub sample of the youngest 5,000 participants with correction for random effects (right lower panel). The y-axis shows the prevalence in % and the x-axis the time in months. For the mixed-effect models, the random effects were removed before plotting and thus yielded lower prevalence estimates. The grey rectangles denoted the three lockdowns in the Netherlands.

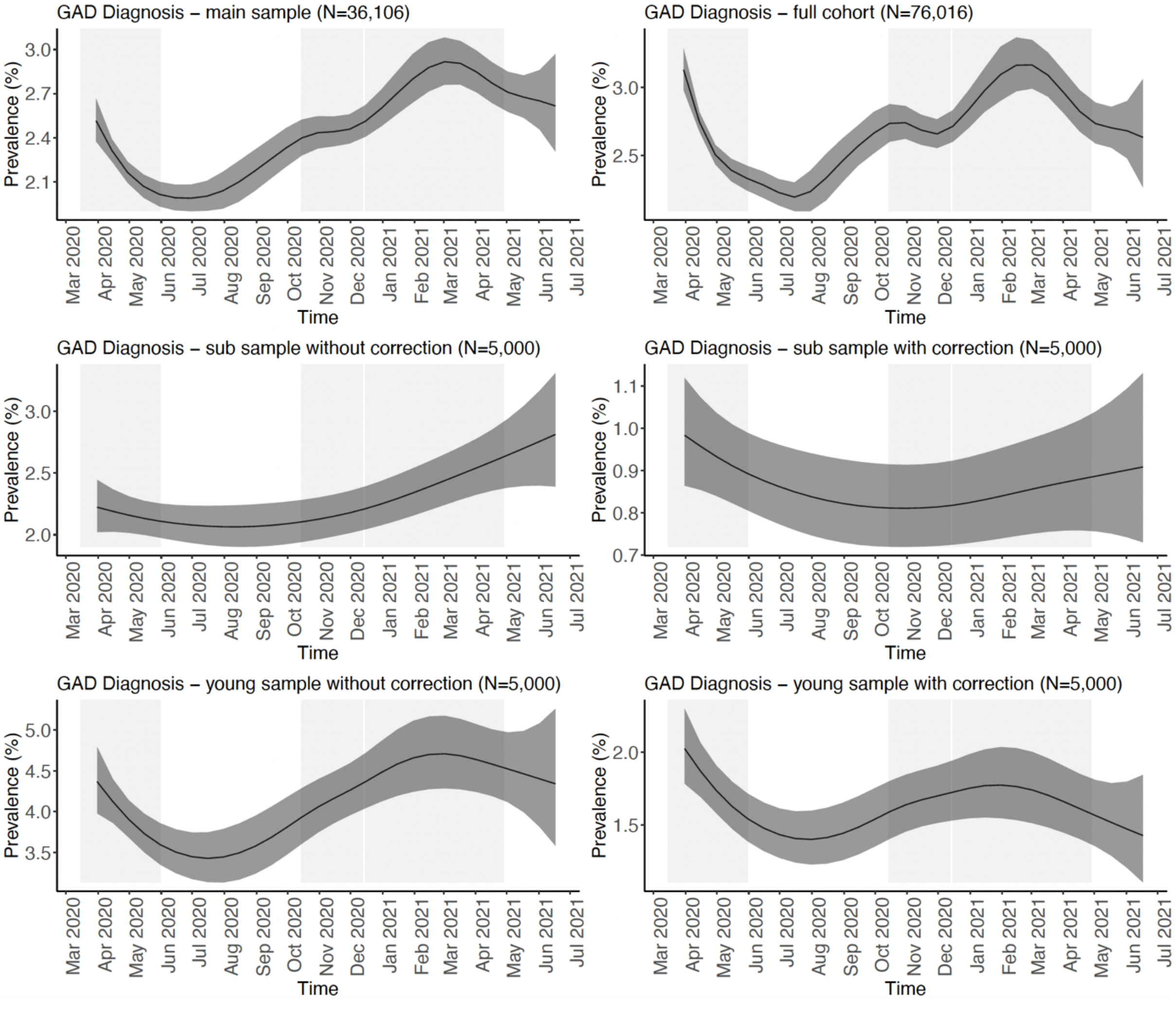

**Figure S8. The longitudinal trajectory of suicidal ideations during the COVID-19 pandemic in the Lifelines cohort.** Shown are the fitted smooths with standard errors of suicidal ideation prevalence over time. Results of GAM analyses are shown for the following analyses: 1) main analysis sample of N=36,106 participants (left upper panel), 2) full cohort of N=76,016 participants (right upper panel), 3) a sub sample of 5,000 participants selected from analysis (1) without correction for random effects (left mid panel), 4) the same sub sample of 5,000 participants as (3) with correction for random effects (right mid panel), 5) a sub sample of the youngest 5,000 participants without correction for random effects (left lower panel), and 6) a sub sample of the youngest 5,000 participants with correction for random effects (right lower panel). The y-axis shows the prevalence in % and the x-axis the time in months. For the mixed-effect models, the random effects were removed before plotting and thus yielded lower prevalence estimates. The grey rectangles denoted the three lockdowns in the Netherlands.

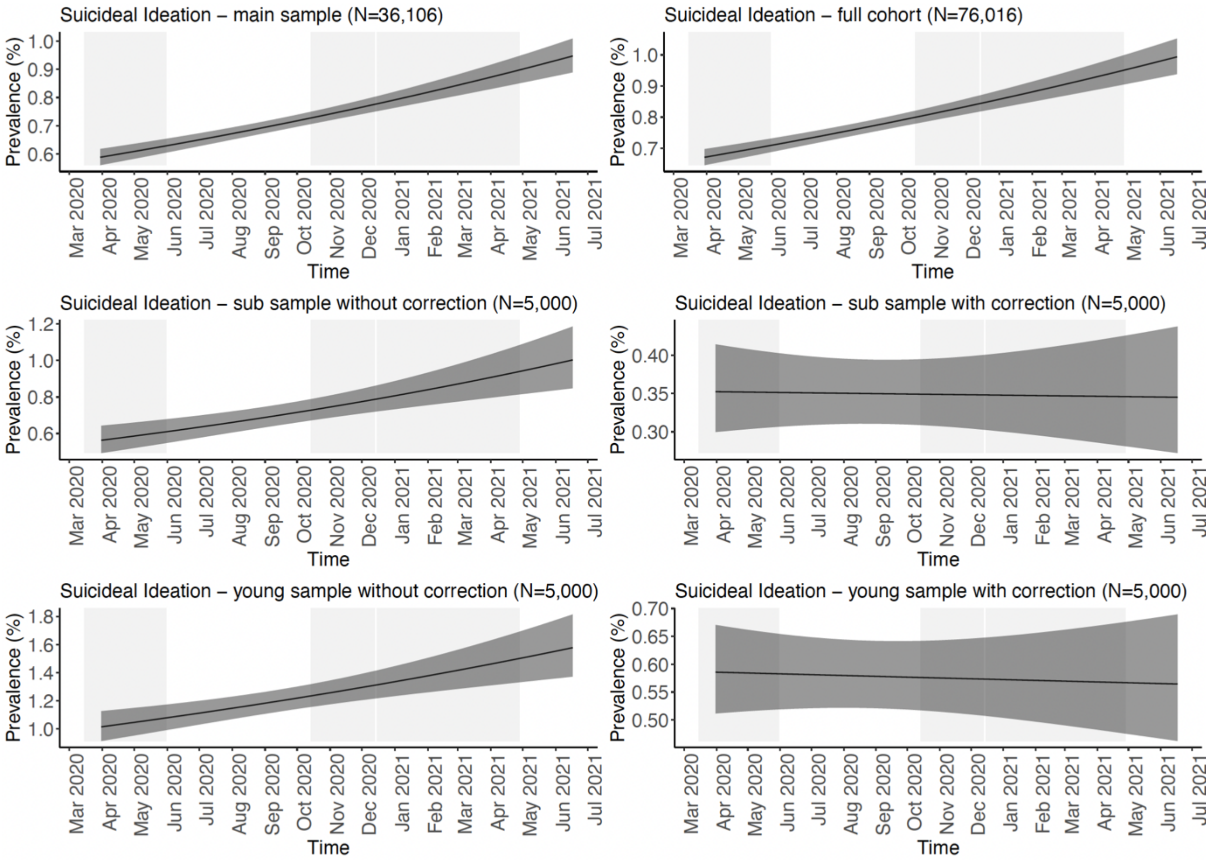

#### 3. Figure S9-15 - longitudinal trajectories of phenotypic outcomes by age

**Figure S9. The trajectory of depressive symptoms during the COVID-19 pandemic by time and age in the Lifelines cohort.** Shown are contour plots that visualize the two-dimensional space of the tensor product interaction between time and age on depressive symptoms. Results of GAM analyses are shown for the following analyses: 1) main analysis sample of N=36,106 participants (left upper panel), 2) full cohort of N=76,016 participants (right upper panel), 3) a sub sample of 5,000 participants selected from analysis (1) without correction for random effects (left mid panel), 4) the same sub sample of 5,000 participants as (3) with correction for random effects (right mid panel), 5) a sub sample of the youngest 5,000 participants without correction for random effects (left lower panel), and 6) a sub sample of the youngest 5,000 participants with correction for random effects (right lower panel). The y-axis shows age in years and the x-axis the time in months. The standardized interaction effect is color-coded with green showing more symptoms and purple showing less symptoms.

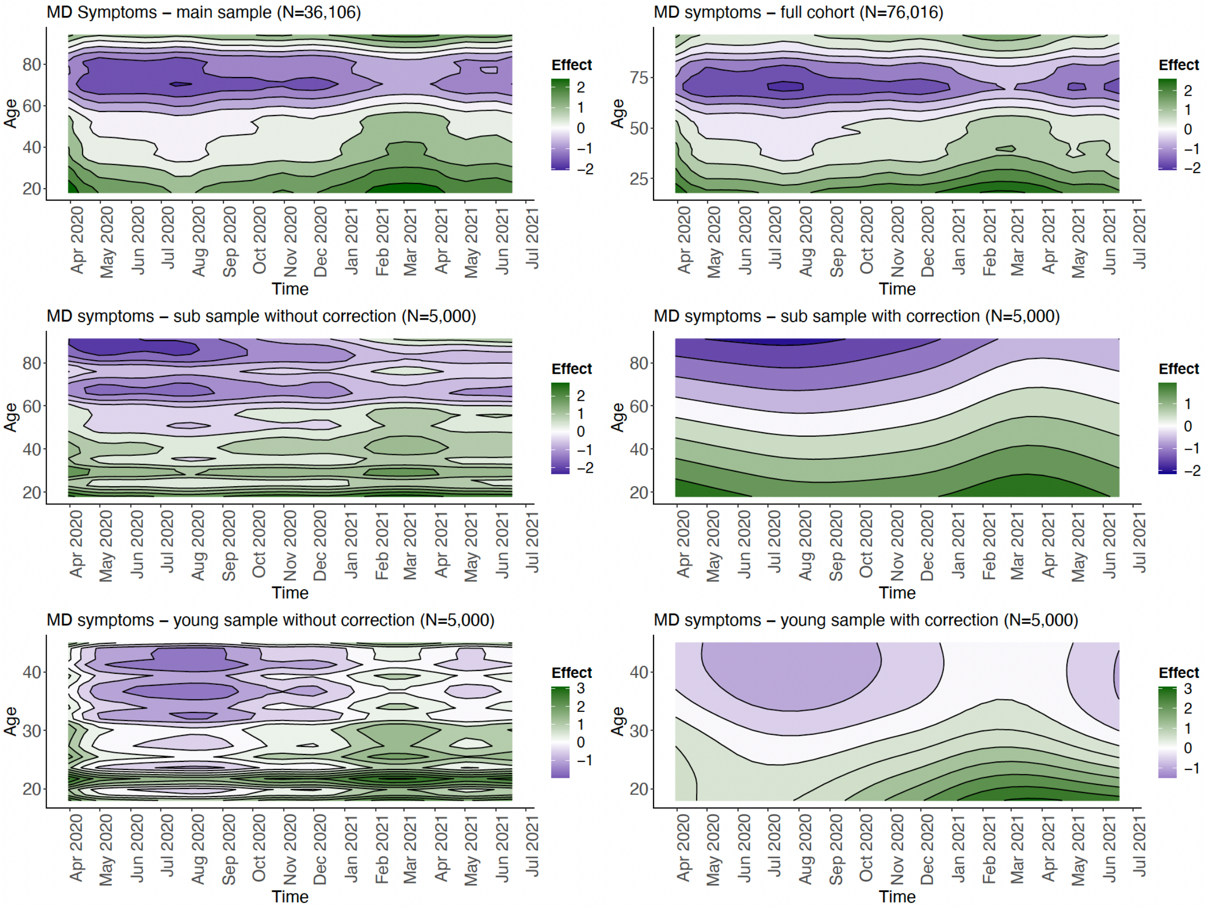

**Figure S10. The trajectory of MDD during the COVID-19 pandemic by time and age in the Lifelines cohort.** Shown are contour plots that visualize the two-dimensional space of the tensor product interaction between time and age on the prevalence of MDD. Results of GAM analyses are shown for the following analyses: 1) main analysis sample of N=36,106 participants (left upper panel), 2) full cohort of N=76,016 participants (right upper panel), 3) a sub sample of 5,000 participants selected from analysis (1) without correction for random effects (left mid panel), 4) the same sub sample of 5,000 participants as (3) with correction for random effects (right mid panel), 5) a sub sample of the youngest 5,000 participants without correction for random effects (left lower panel), and 6) a sub sample of the youngest 5,000 participants with correction for random effects (right lower panel). The y-axis shows age in years and the x-axis the time in months. The standardized interaction effect is color-coded with green showing a higher prevalence and purple showing a lower prevalence.

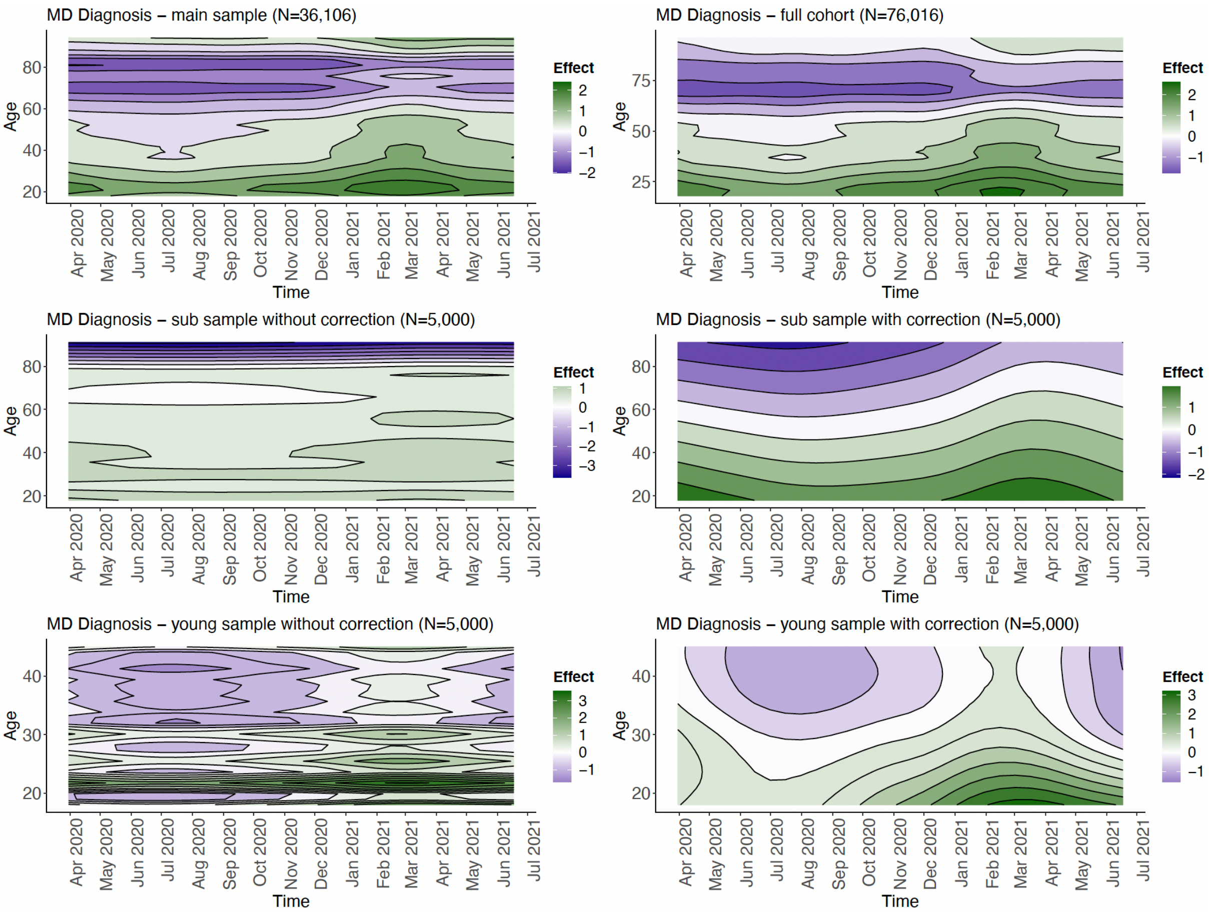

**Figure S11. The trajectory of anxiety symptoms during the COVID-19 pandemic by time and age in the Lifelines cohort.** Shown are contour plots that visualize the two-dimensional space of the tensor product interaction between time and age on anxiety symptoms. Results of GAM analyses are shown for the following analyses: 1) main analysis sample of N=36,106 participants (left upper panel), 2) full cohort of N=76,016 participants (right upper panel), 3) a sub sample of 5,000 participants selected from analysis (1) <u>without correction</u> for random effects (left mid panel), 4) the same sub sample of 5,000 participants as (3) <u>with correction</u> for random effects (right mid panel), 5) a sub sample of the youngest 5,000 participants <u>without correction</u> for random effects (left lower panel), and 6) a sub sample of the youngest 5,000 participants <u>with correction</u> for random effects (right lower panel). The y-axis shows age in years and the x-axis the time in months. The standardized interaction effect is color-coded with green showing more symptoms and purple showing less symptoms.

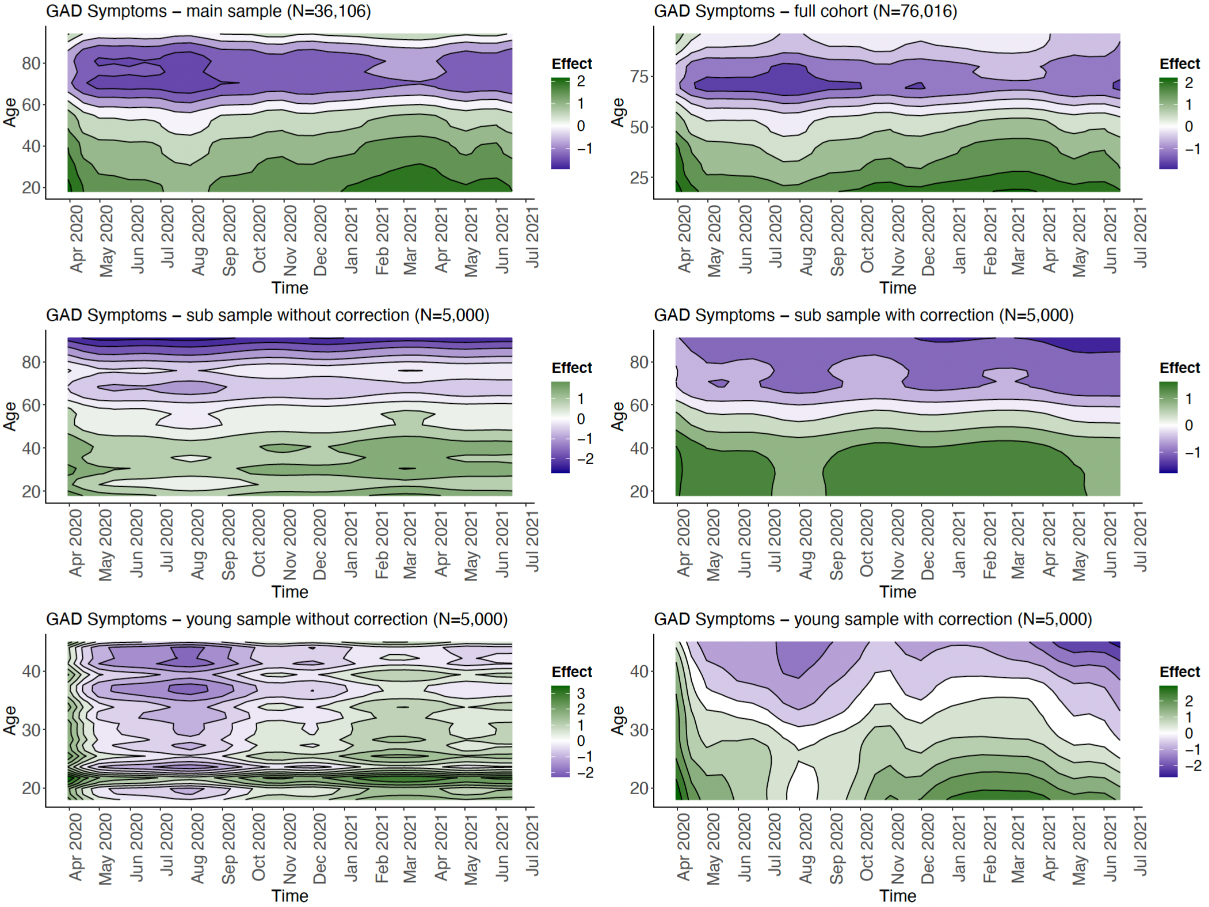

**Figure S12. The trajectory of GAD during the COVID-19 pandemic by time and age in the Lifelines cohort.** Shown are contour plots that visualize the two-dimensional space of the tensor product interaction between time and age on the prevalence of GAD. Results of GAM analyses are shown for the following analyses: 1) main analysis sample of N=36,106 participants (left upper panel), 2) full cohort of N=76,016 participants (right upper panel), 3) a sub sample of 5,000 participants selected from analysis (1) without correction for random effects (left mid panel), 4) the same sub sample of 5,000 participants as (3) with correction for random effects (right mid panel), 5) a sub sample of the youngest 5,000 participants without correction for random effects (left lower panel), and 6) a sub sample of the youngest 5,000 participants with correction for random effects (right lower panel). The y-axis shows age in years and the x-axis the time in months. The standardized interaction effect is color-coded with green showing a higher prevalence and purple showing a lower prevalence.

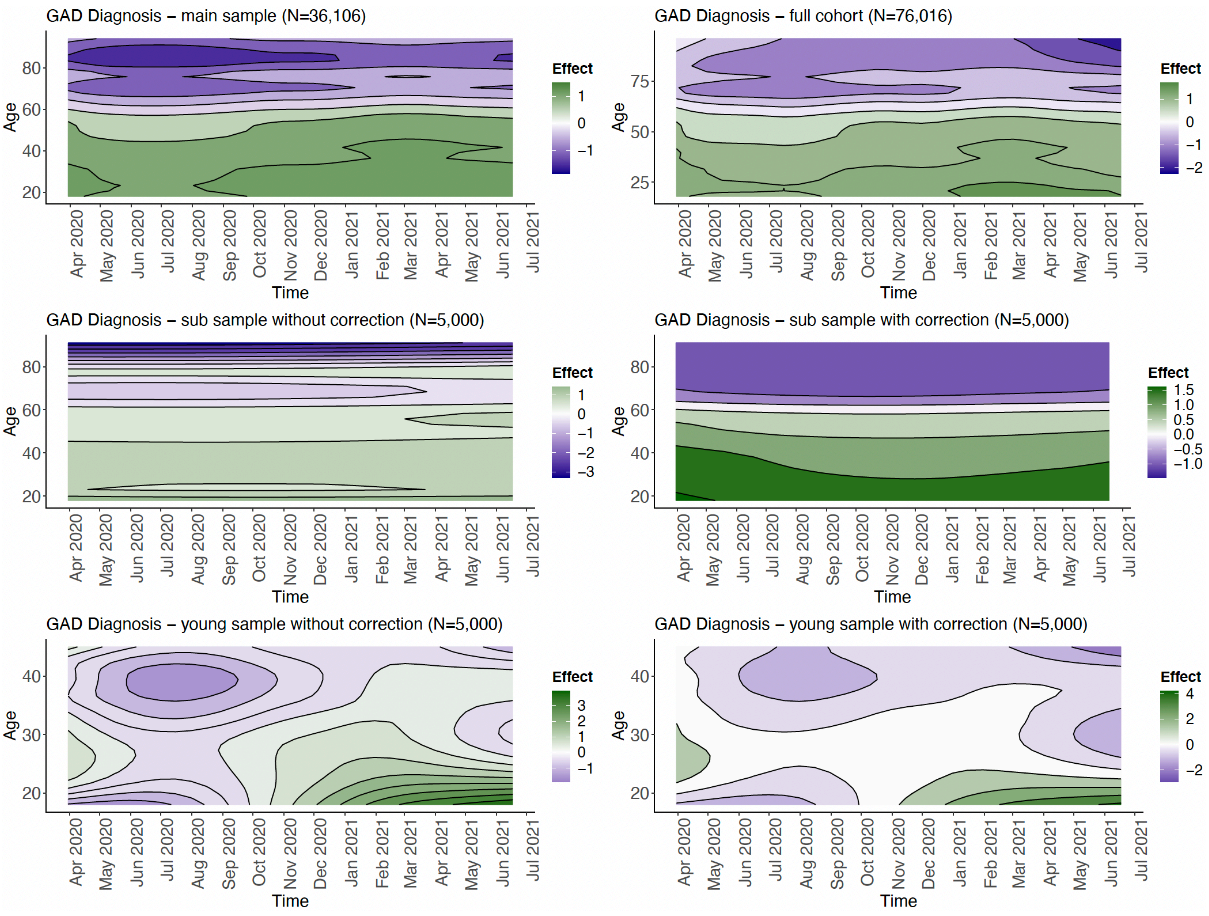

**Figure S13. The trajectory of suicidal ideation during the COVID-19 pandemic by time and age in the Lifelines cohort.** Shown are contour plots that visualize the two-dimensional space of the tensor product interaction between time and age on the prevalence of suicidal ideation. Results of GAM analyses are shown for the following analyses: 1) main analysis sample of N=36,106 participants (left upper panel), 2) full cohort of N=76,016 participants (right upper panel), 3) a sub sample of 5,000 participants selected from analysis (1) without correction for random effects (left mid panel), 4) the same sub sample of 5,000 participants as (3) with correction for random effects (right mid panel), 5) a sub sample of the youngest 5,000 participants without correction for random effects (left lower panel), and 6) a sub sample of the youngest 5,000 participants with correction for random effects (right lower panel). The y-axis shows age in years and the x-axis the time in months. The standardized interaction effect is color-coded with green showing a higher prevalence and purple showing a lower prevalence.

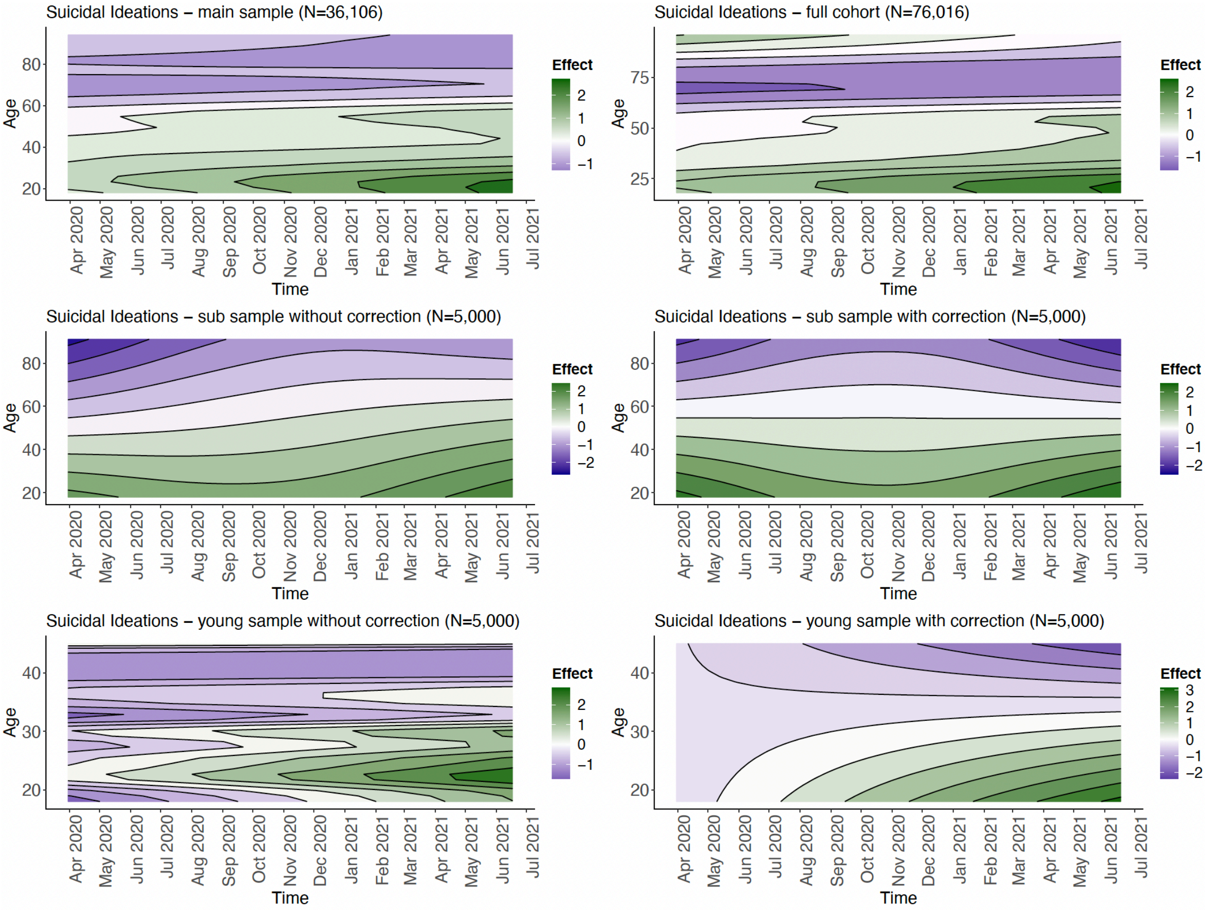

**Figure S14. The trajectory of suicidal ideation during the COVID-19 pandemic by specific age groups in the Lifelines cohort.** Shown are the prevalence of suicidal ideation over time as estimated by the GAM tensor product interaction between time and age per specific age groups. Results of GAM analyses are shown for the following analyses: 1) main analysis sample of N=36,106 participants (left upper panel), 2) full cohort of N=76,016 participants (right upper panel), 3) a sub sample of 5,000 participants selected from analysis (1) without correction for random effects (left mid panel), 4) the same sub sample of 5,000 participants as (3) with correction for random effects (right mid panel), 5) a sub sample of the youngest 5,000 participants without correction for random effects (left lower panel), and 6) a sub sample of the youngest 5,000 participants with correction for random effects (right lower panel). The y-axis shows the prevalence in percentages and the x-axis the time in months. The age groups are color-coded with dark red showing younger age groups and light red showing older age groups. For the mixed-effect models, the random effects were removed before plotting and thus yielded lower prevalence estimates. The grey rectangles denoted the three lockdowns in the Netherlands.

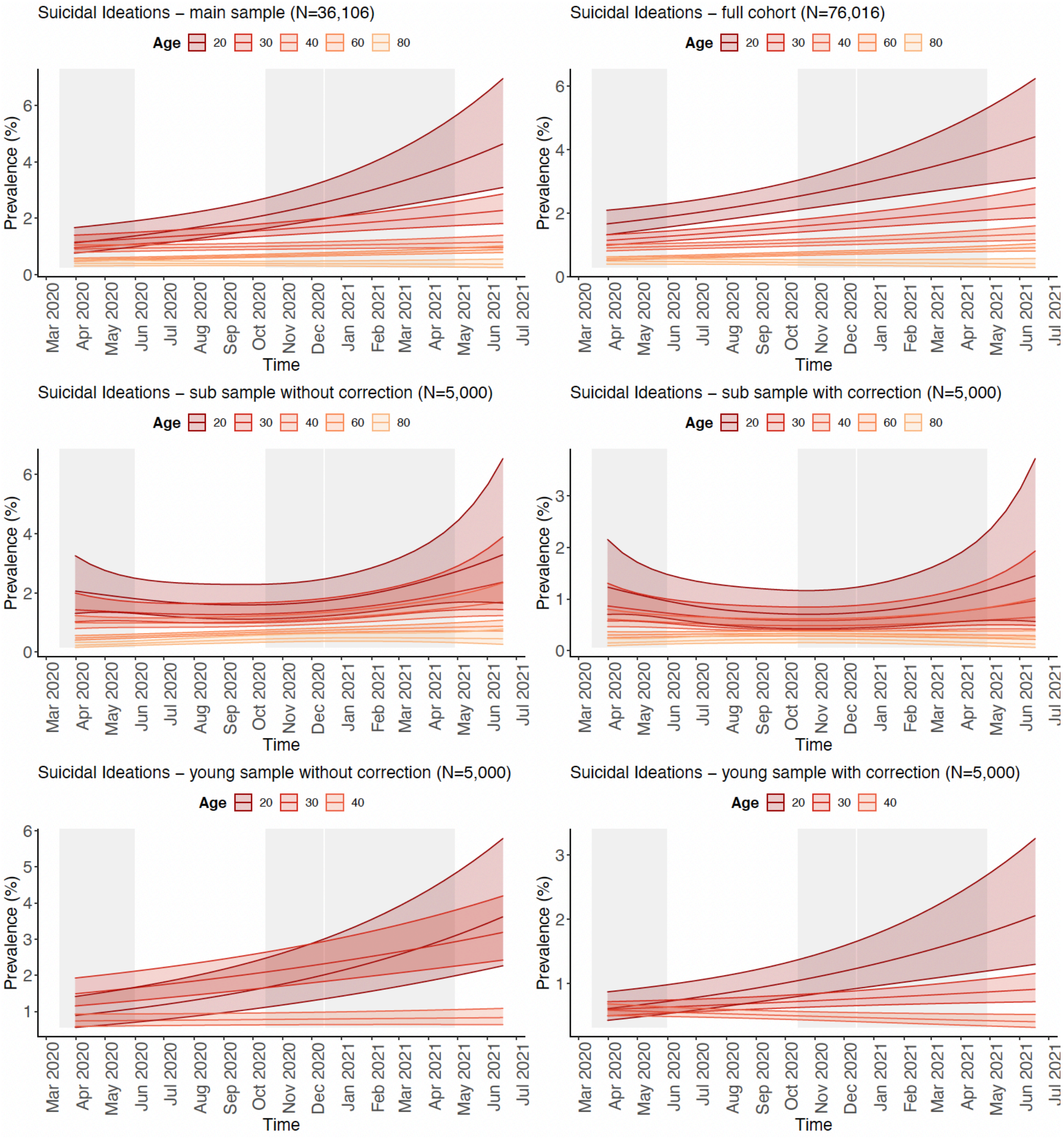

**Figure S15. The trajectory of suicidal ideation during the COVID-19 pandemic by specific time points in the Lifelines cohort.** Shown are the prevalence of suicidal ideation over time as estimated by the GAM tensor product interaction between time and age per specific time points. Results of GAM analyses are shown for the following analyses: 1) main analysis sample of N=36,106 participants (left upper panel), 2) full cohort of N=76,016 participants (right upper panel), 3) a sub sample of 5,000 participants selected from analysis (1) without correction for random effects (left mid panel), 4) the same sub sample of 5,000 participants as (3) with correction for random effects (right mid panel), 5) a sub sample of the youngest 5,000 participants without correction for random effects (left lower panel), and 6) a sub sample of the youngest 5,000 participants with correction for random effects (right lower panel). The y-axis shows the prevalence in percentages and the x-axis continuous age. The time points are color-coded with the specific date shown in the legend. For the mixed-effect models, the random effects were removed before plotting.

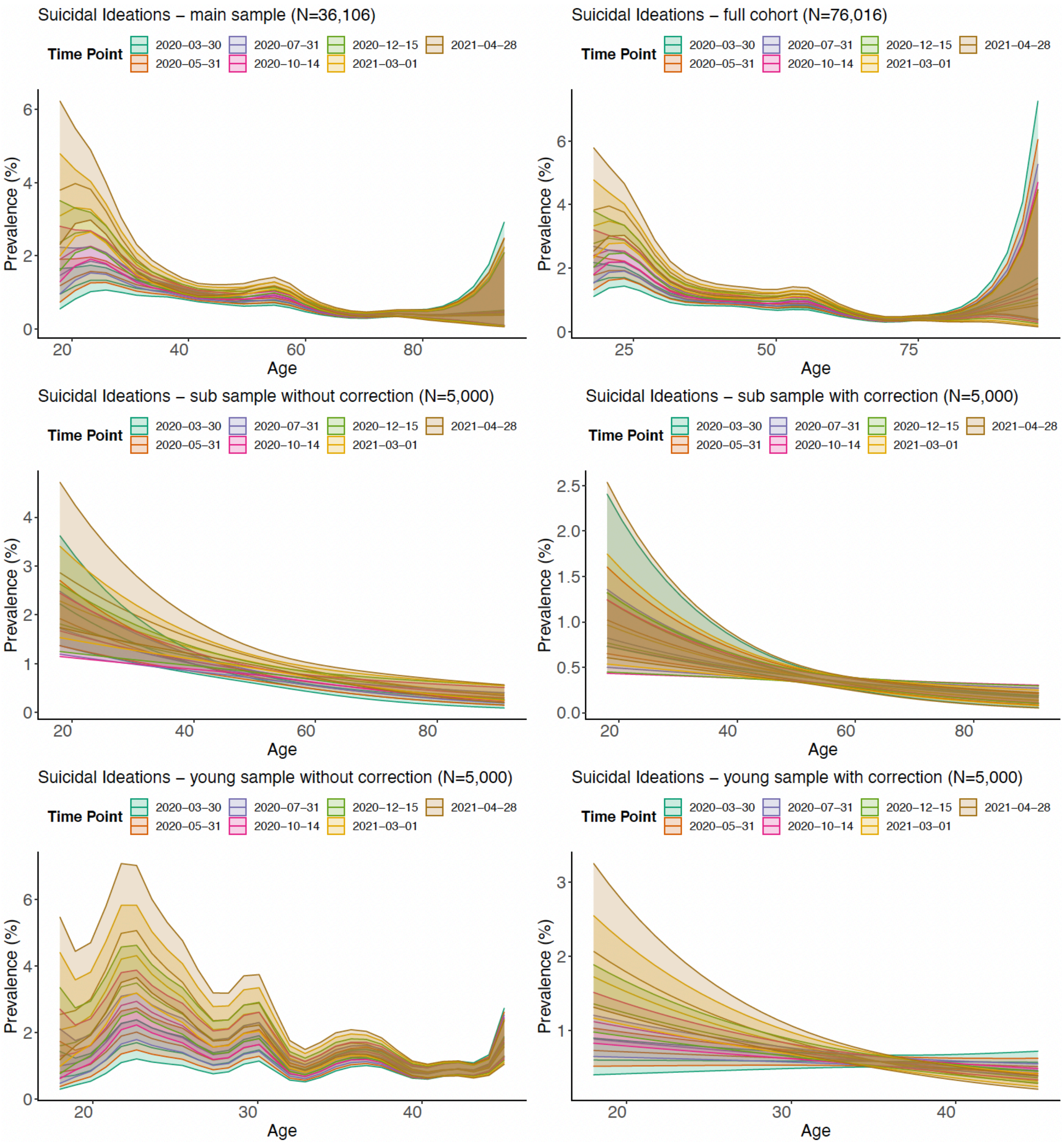

#### 4. Figure S16-20 - longitudinal trajectories of phenotypic outcomes by sex

**Figure S16. The longitudinal trajectory of depressive symptoms during the COVID-19 pandemic by sex in the Lifelines cohort.** Shown are the fitted smooths with standard errors of mean depressive symptom score over time for each sex. Results of GAM analyses are shown for the following analyses: 1) main analysis sample of N=36,106 participants (left upper panel), 2) full cohort of N=76,016 participants (right upper panel), 3) a sub sample of 5,000 participants selected from analysis (1) without correction for random effects (left mid panel), 4) the same sub sample of 5,000 participants as (3) with correction for random effects (right mid panel), 5) a sub sample of the youngest 5,000 participants without correction for random effects (left lower panel), and 6) a sub sample of the youngest 5,000 participants with correction for random effects (right lower panel). The y-axis shows the mean symptom sum score and the x-axis the time in months. For the mixed-effect models, the random effects were removed before plotting and thus yielded lower prevalence estimates. The grey rectangles denoted the three lockdowns in the Netherlands.

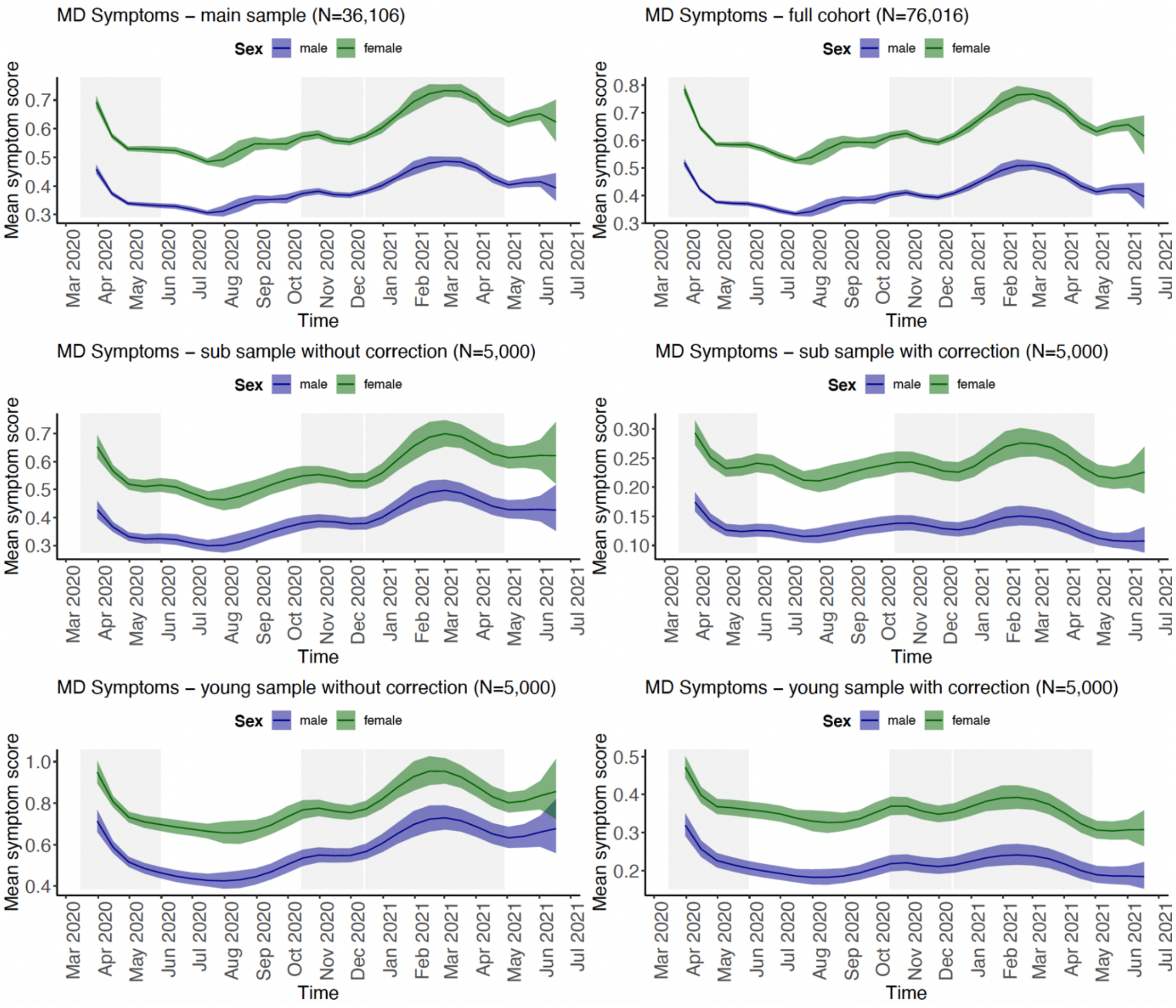

**Figure S17. The longitudinal trajectory of MDD during the COVID-19 pandemic by sex in the Lifelines cohort.** Shown are the fitted smooths with standard errors of MDD prevalence over time for each sex. Results of GAM analyses are shown for the following analyses: 1) main analysis sample of N=36,106 participants (left upper panel), 2) full cohort of N=76,016 participants (right upper panel), 3) a sub sample of 5,000 participants selected from analysis (1) <u>without correction</u> for random effects (left mid panel), 4) the same sub sample of 5,000 participants as (3) <u>with correction</u> for random effects (right mid panel), 5) a sub sample of the youngest 5,000 participants <u>without correction</u> for random effects (left lower panel), and 6) a sub sample of the youngest 5,000 participants <u>with correction</u> for random effects (right lower panel). The y-axis shows the prevalence in % and the x-axis the time in months. For the mixed-effect models, the random effects were removed before plotting and thus yielded lower prevalence estimates. The grey rectangles denoted the three lockdowns in the Netherlands.

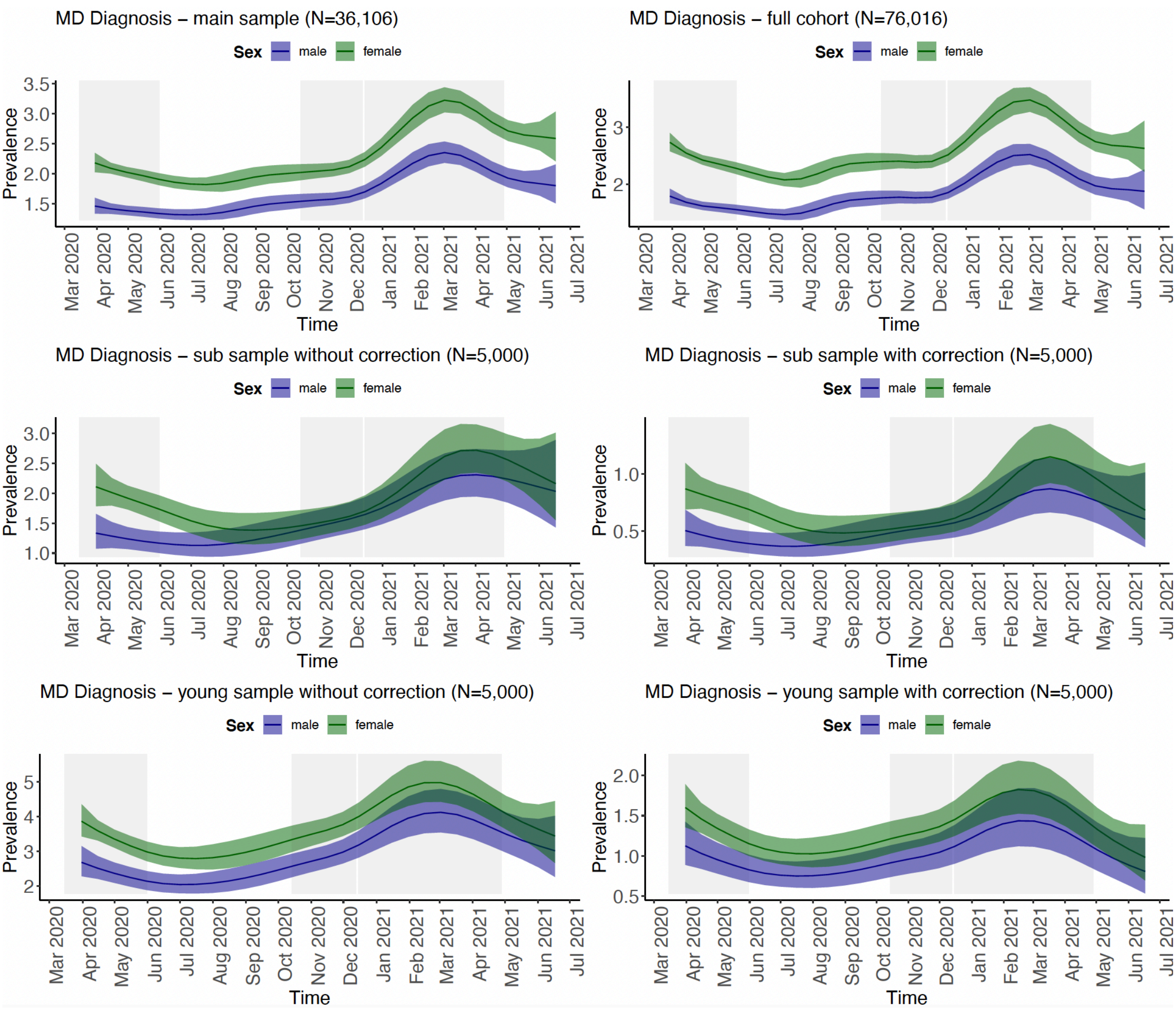

**Figure S18. The longitudinal trajectory of anxiety symptoms during the COVID-19 pandemic by sex in the Lifelines cohort.** Shown are the fitted smooths with standard errors of mean anxiety symptom score over time for each sex. Results of GAM analyses are shown for the following analyses: 1) main analysis sample of N=36,106 participants (left upper panel), 2) full cohort of N=76,016 participants (right upper panel), 3) a sub sample of 5,000 participants selected from analysis (1) without correction for random effects (left mid panel), 4) the same sub sample of 5,000 participants as (3) with correction for random effects (right mid panel), 5) a sub sample of the youngest 5,000 participants without correction for random effects (left lower panel), and 6) a sub sample of the youngest 5,000 participants with correction for random effects (right lower panel). The y-axis shows the mean anxiety symptom sum score and the x-axis the time in months. For the mixed-effect models, the random effects were removed before plotting and thus yielded lower prevalence estimates. The grey rectangles denoted the three lockdowns in the Netherlands.

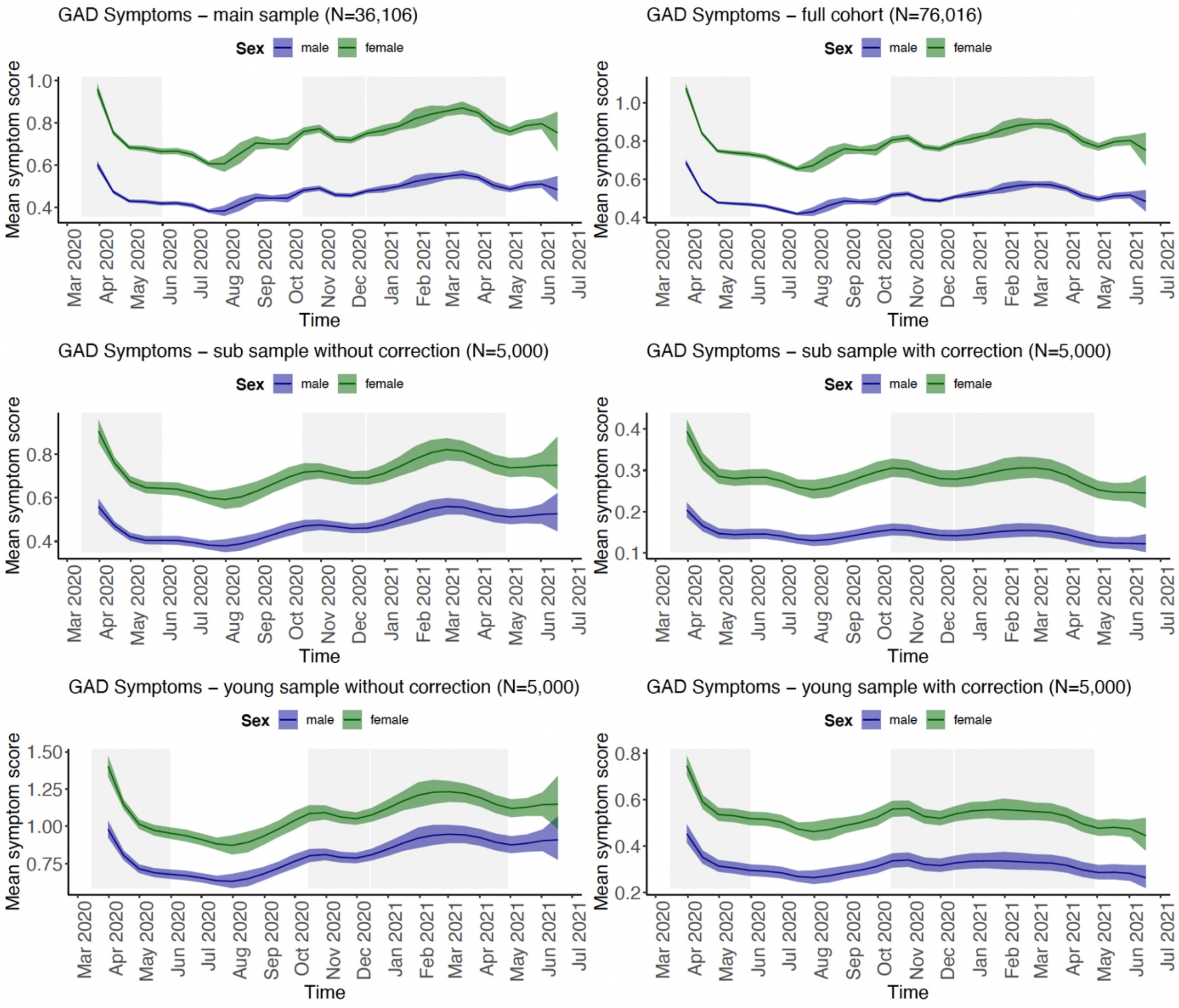

**Figure S19. The longitudinal trajectory of GAD during the COVID-19 pandemic in the Lifelines cohort by sex.** Shown are the fitted smooths with standard errors of GAD prevalence over time for each sex. Results of GAM analyses are shown for the following analyses: 1) main analysis sample of N=36,106 participants (left upper panel), 2) full cohort of N=76,016 participants (right upper panel), 3) a sub sample of 5,000 participants selected from analysis (1) without correction for random effects (left mid panel), 4) the same sub sample of 5,000 participants as (3) with correction for random effects (right mid panel), 5) a sub sample of the youngest 5,000 participants without correction for random effects (left lower panel), and 6) a sub sample of the youngest 5,000 participants with correction for random effects (right lower panel). The y-axis shows the prevalence in % and the x-axis the time in months. For the mixed-effect models, the random effects were removed before plotting and thus yielded lower prevalence estimates. The grey rectangles denoted the three lockdowns in the Netherlands.

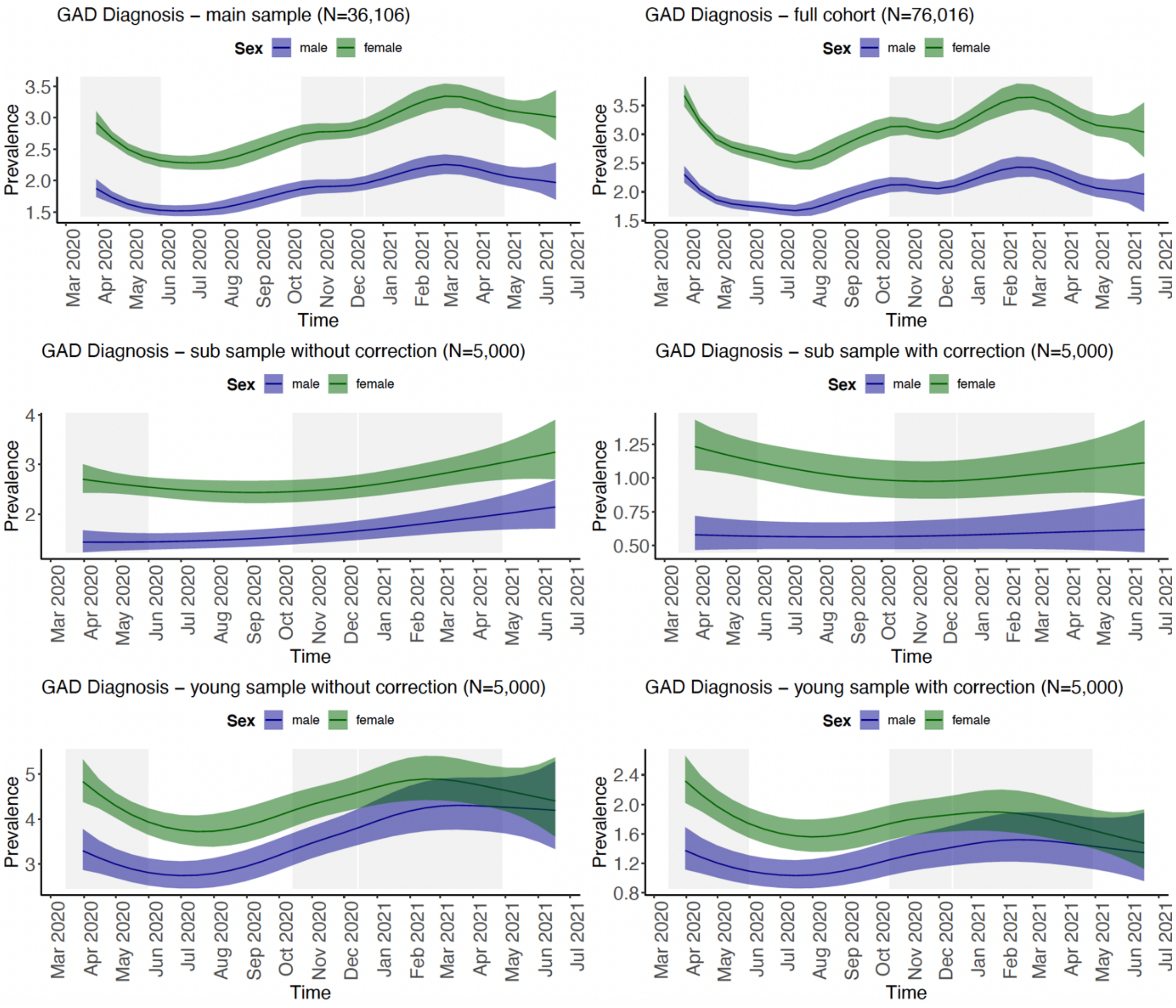

**Figure S20. The longitudinal trajectory of suicidal ideations during the COVID-19 pandemic by sex in the Lifelines cohort.** Shown are the fitted smooths with standard errors of suicidal ideation prevalence over time for each sex. Results of GAM analyses are shown for the following analyses: 1) main analysis sample of N=36,106 participants (left upper panel), 2) full cohort of N=76,016 participants (right upper panel), 3) a sub sample of 5,000 participants selected from analysis (1) without correction for random effects (left mid panel), 4) the same sub sample of 5,000 participants as (3) with correction for random effects (right mid panel), 5) a sub sample of the youngest 5,000 participants without correction for random effects (left lower panel), and 6) a sub sample of the youngest 5,000 participants with correction for random effects (right lower panel). The y-axis shows the prevalence in % and the x-axis the time in months. For the mixed-effect models, the random effects were removed before plotting and thus yielded lower prevalence estimates. The grey rectangles denoted the three lockdowns in the Netherlands.

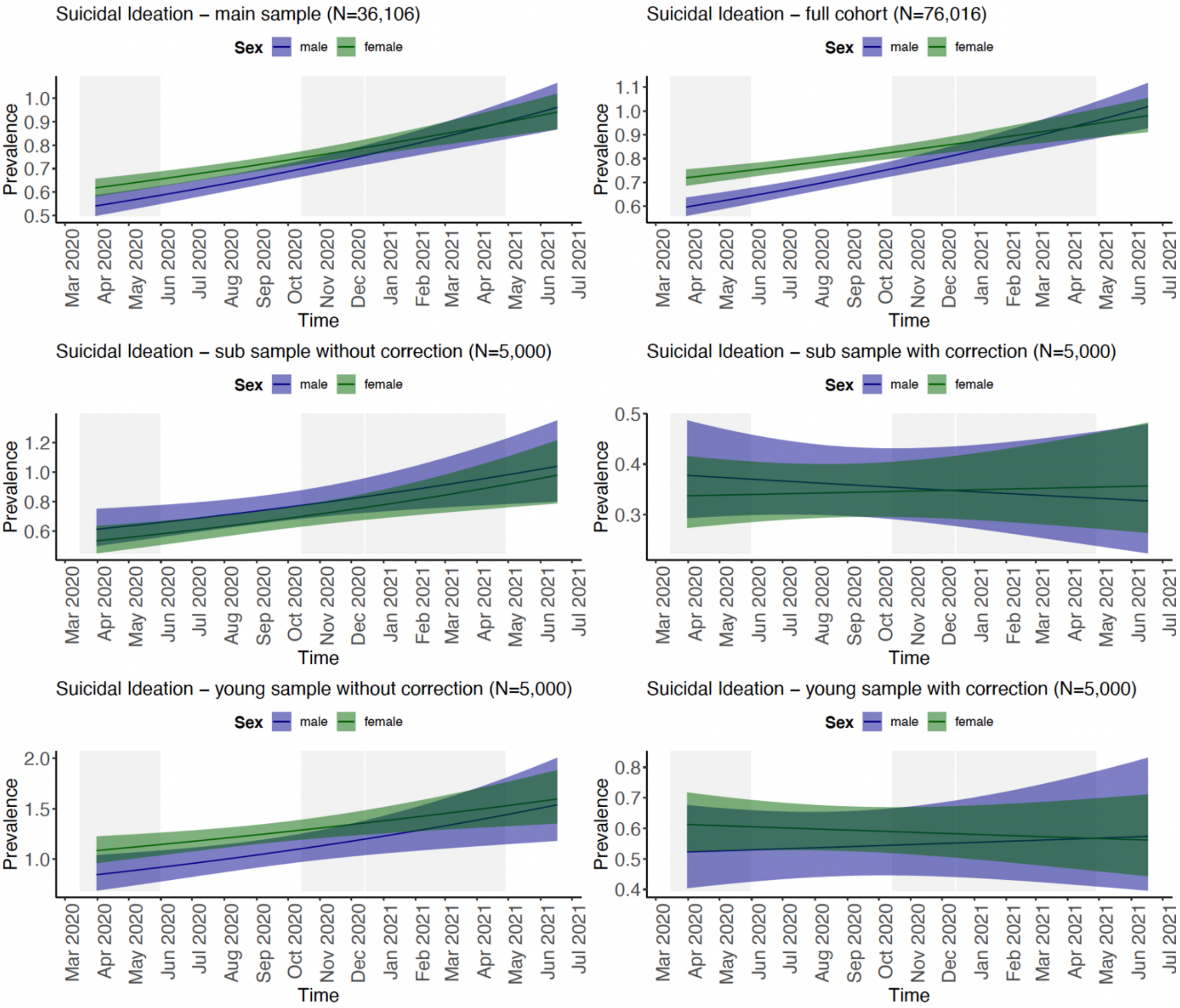

#### 5. Figure S21-25 - longitudinal trajectories of phenotypic outcomes by sex visualized using difference plots

**Figure S21. The difference between sexes in their trajectory of depressive symptoms during the COVID-19 pandemic in the Lifelines cohort.** Shown are the difference in fitted smooths of mean depressive symptom score between the sexes over time. Results of GAM analyses are shown for the following analyses: 1) main analysis sample of N=36,106 participants (left upper panel), 2) full cohort of N=76,016 participants (right upper panel), 3) a sub sample of 5,000 participants selected from analysis (1) without correction for random effects (left mid panel), 4) the same sub sample of 5,000 participants as (3) with correction for random effects (right mid panel), 5) a sub sample of the youngest 5,000 participants without correction for random effects (left lower panel), and 6) a sub sample of the youngest 5,000 participants with correction for random effects (right lower panel). The y-axis shows the mean difference in symptom sum score and the x-axis the time in months. The grey rectangles denoted the three lockdowns in the Netherlands.

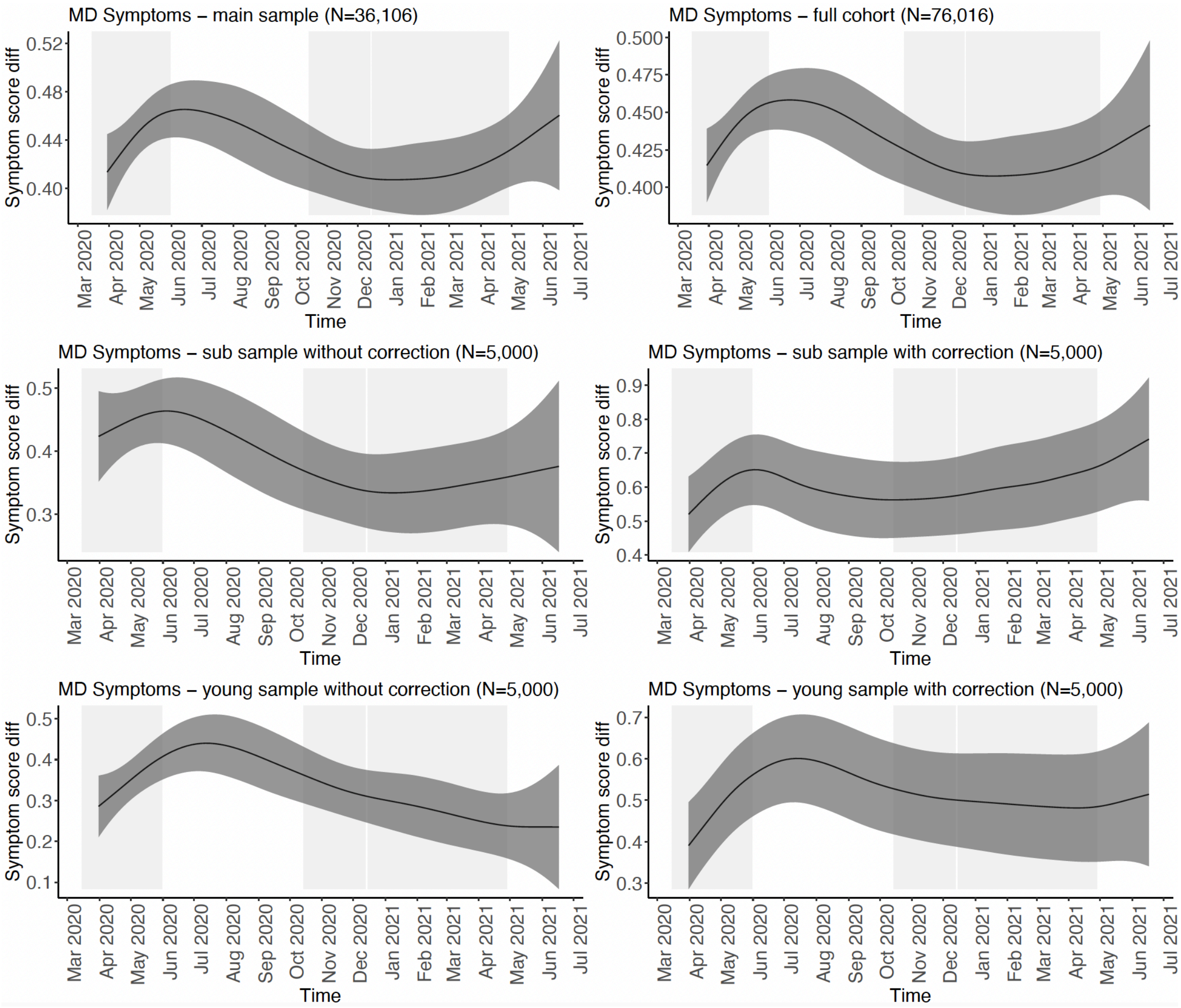

**Figure S22. The difference between sexes in their trajectory of MDD during the COVID-19 pandemic in the Lifelines cohort.** Shown are the difference in fitted smooths of the prevalence between the sexes over time. Results of GAM analyses are shown for the following analyses: 1) main analysis sample of N=36,106 participants (left upper panel), 2) full cohort of N=76,016 participants (right upper panel), 3) a sub sample of 5,000 participants selected from analysis (1) <u>without correction</u> for random effects (left mid panel), 4) the same sub sample of 5,000 participants as (3) <u>with correction</u> for random effects (right mid panel), 5) a sub sample of the youngest 5,000 participants <u>without correction</u> for random effects (left lower panel), and 6) a sub sample of the youngest 5,000 participants <u>with correction</u> for random effects (right lower panel). The y-axis shows the difference in prevalence in % and the x-axis the time in months. The grey rectangles denoted the three lockdowns in the Netherlands.

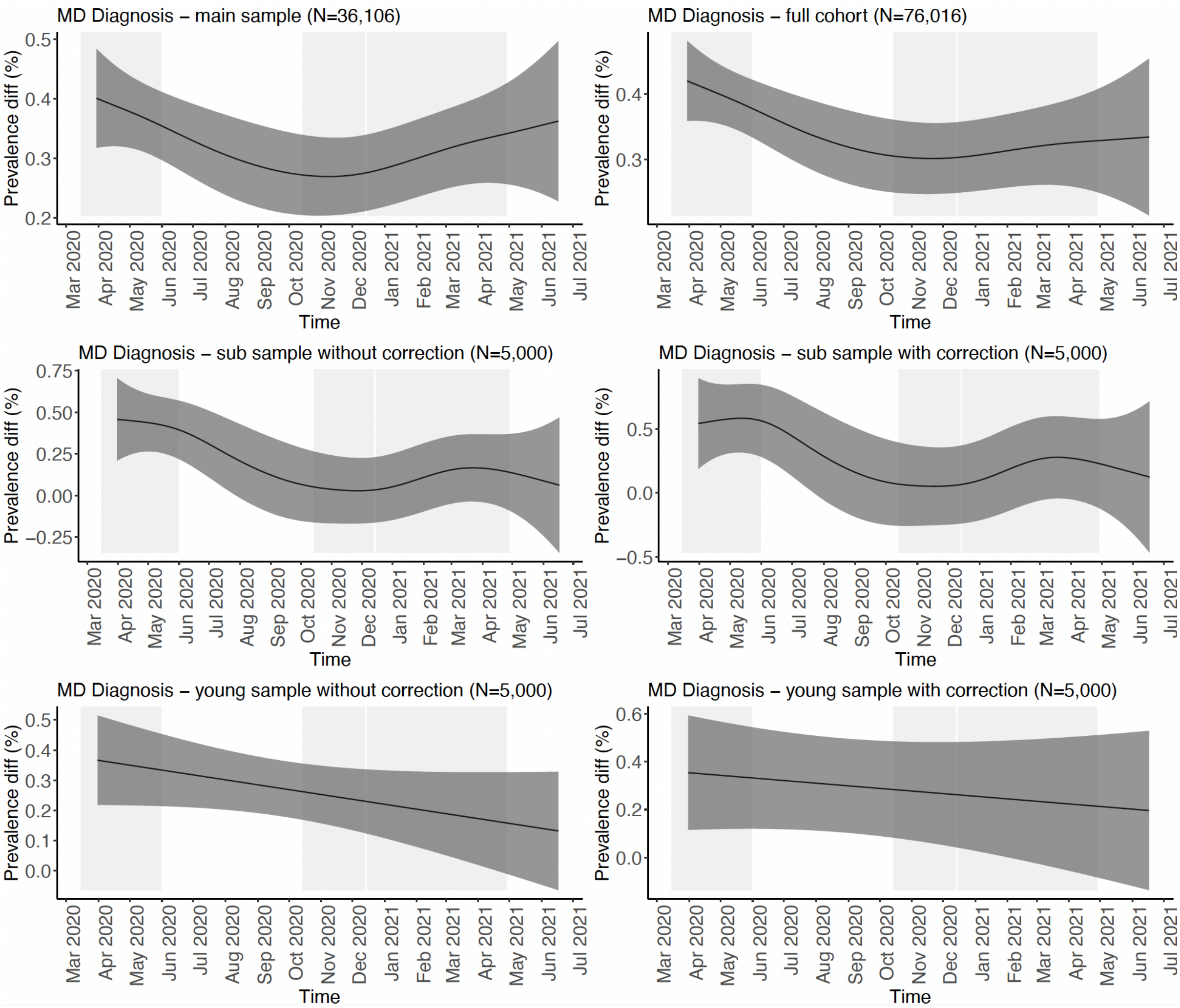

**Figure S23. The difference between sexes in their trajectory of mean anxiety score during the COVID-19 pandemic in the Lifelines cohort.** Shown are the difference in fitted smooths of mean anxiety symptom score between the sexes over time. Results of GAM analyses are shown for the following analyses: 1) main analysis sample of N=36,106 participants (left upper panel), 2) full cohort of N=76,016 participants (right upper panel), 3) a sub sample of 5,000 participants selected from analysis (1) without correction for random effects (left mid panel), 4) the same sub sample of 5,000 participants as (3) with correction for random effects (right mid panel), 5) a sub sample of the youngest 5,000 participants without correction for random effects (left lower panel), and 6) a sub sample of the youngest 5,000 participants with correction for random effects (right lower panel). The y-axis shows the difference in mean anxiety symptom sum score and the x-axis the time in months. The grey rectangles denoted the three lockdowns in the Netherlands.

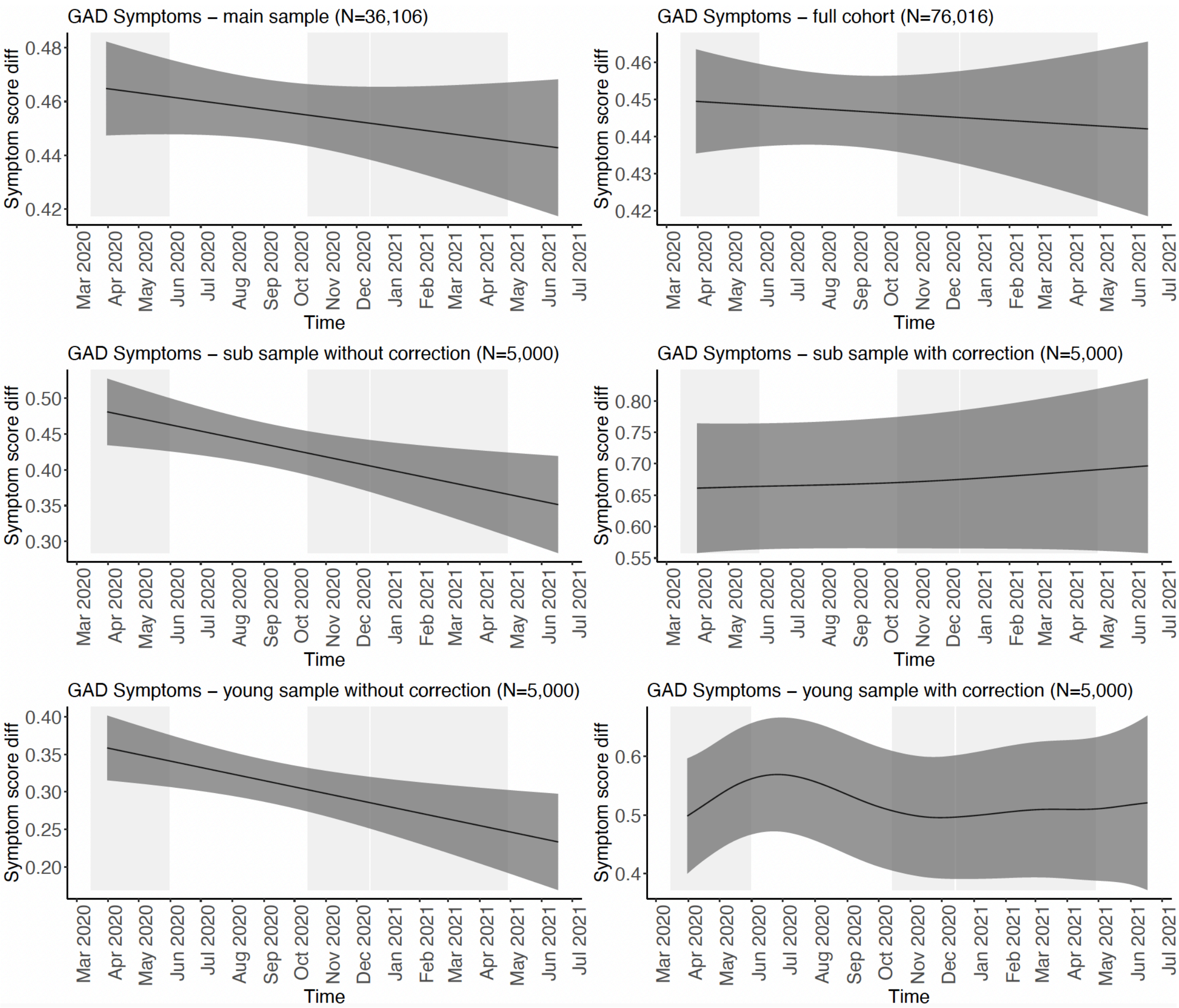

**Figure S24. The difference between sexes in their trajectory of GAD during the COVID-19 pandemic in the Lifelines cohort.** Shown are the difference in fitted smooths of the prevalence of GAD between the sexes over time. Results of GAM analyses are shown for the following analyses: 1) main analysis sample of N=36,106 participants (left upper panel), 2) full cohort of N=76,016 participants (right upper panel), 3) a sub sample of 5,000 participants selected from analysis (1) without correction for random effects (left mid panel), 4) the same sub sample of 5,000 participants as (3) with correction for random effects (right mid panel), 5) a sub sample of the youngest 5,000 participants without correction for random effects (left lower panel), and 6) a sub sample of the youngest 5,000 participants with correction for random effects (right lower panel). The y-axis shows the difference in prevalence in % and the x-axis the time in months. The grey rectangles denoted the three lockdowns in the Netherlands.

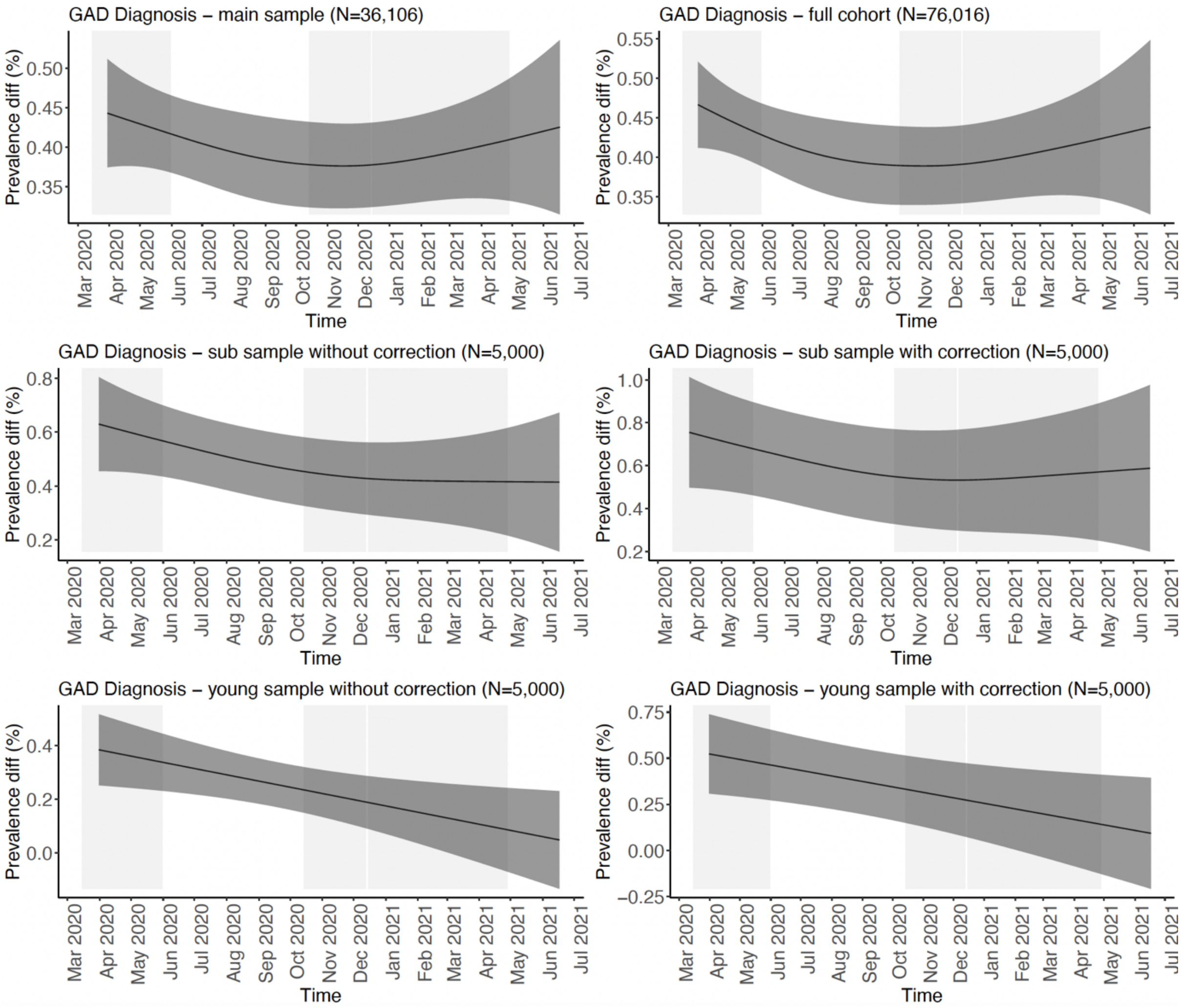

**Figure S25. The difference between sexes in their trajectory of suicidal ideation during the COVID-19 pandemic in the Lifelines cohort.** Shown are the difference in fitted smooths of the prevalence of suicidal ideation between the sexes over time. Results of GAM analyses are shown for the following analyses: 1) main analysis sample of N=36,106 participants (left upper panel), 2) full cohort of N=76,016 participants (right upper panel), 3) a sub sample of 5,000 participants selected from analysis (1) <u>without correction</u> for random effects (left mid panel), 4) the same sub sample of 5,000 participants as (3) <u>with correction</u> for random effects (right mid panel), 5) a sub sample of the youngest 5,000 participants <u>without correction</u> for random effects (left lower panel), and 6) a sub sample of the youngest 5,000 participants <u>with correction</u> for random effects (right lower panel). The y-axis shows the difference in prevalence in % and the x-axis the time in months. The grey rectangles denoted the three lockdowns in the Netherlands.

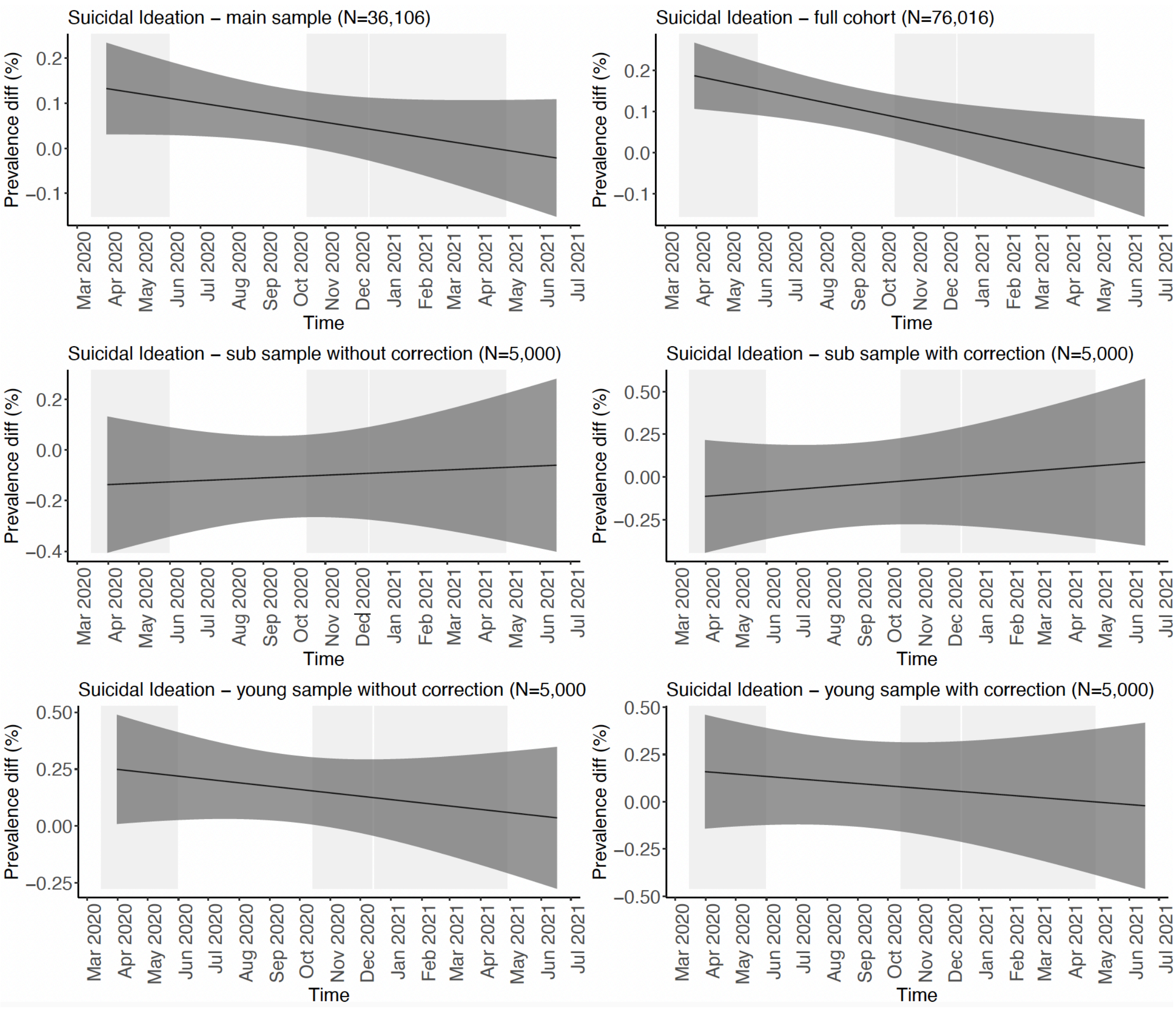

#### 6. Figure S26-30 - longitudinal trajectories of phenotypic outcomes by lifetime history of MDD/GAD

**Figure S26. The longitudinal trajectory of depressive symptoms during the COVID-19 pandemic by lifetime history of MDD in the Lifelines cohort.** Shown are the fitted smooths with standard errors of mean depressive symptom score over time for study participants with and without MDD lifetime history. Results of GAM analyses are shown for the following analyses: 1) main analysis sample of N=36,106 participants (left upper panel), 2) full cohort of N=76,016 participants (right upper panel), 3) a sub sample of 5,000 participants selected from analysis (1) without correction for random effects (left mid panel), 4) the same sub sample of 5,000 participants as (3) with correction for random effects (right mid panel), 5) a sub sample of the youngest 5,000 participants without correction for random effects (left lower panel), and 6) a sub sample of the youngest 5,000 participants with correction for random effects (right lower panel). The y-axis shows the mean symptom sum score and the x-axis the time in months. For the mixed-effect models, the random effects were removed before plotting and thus yielded lower prevalence estimates. The grey rectangles denoted the three lockdowns in the Netherlands.

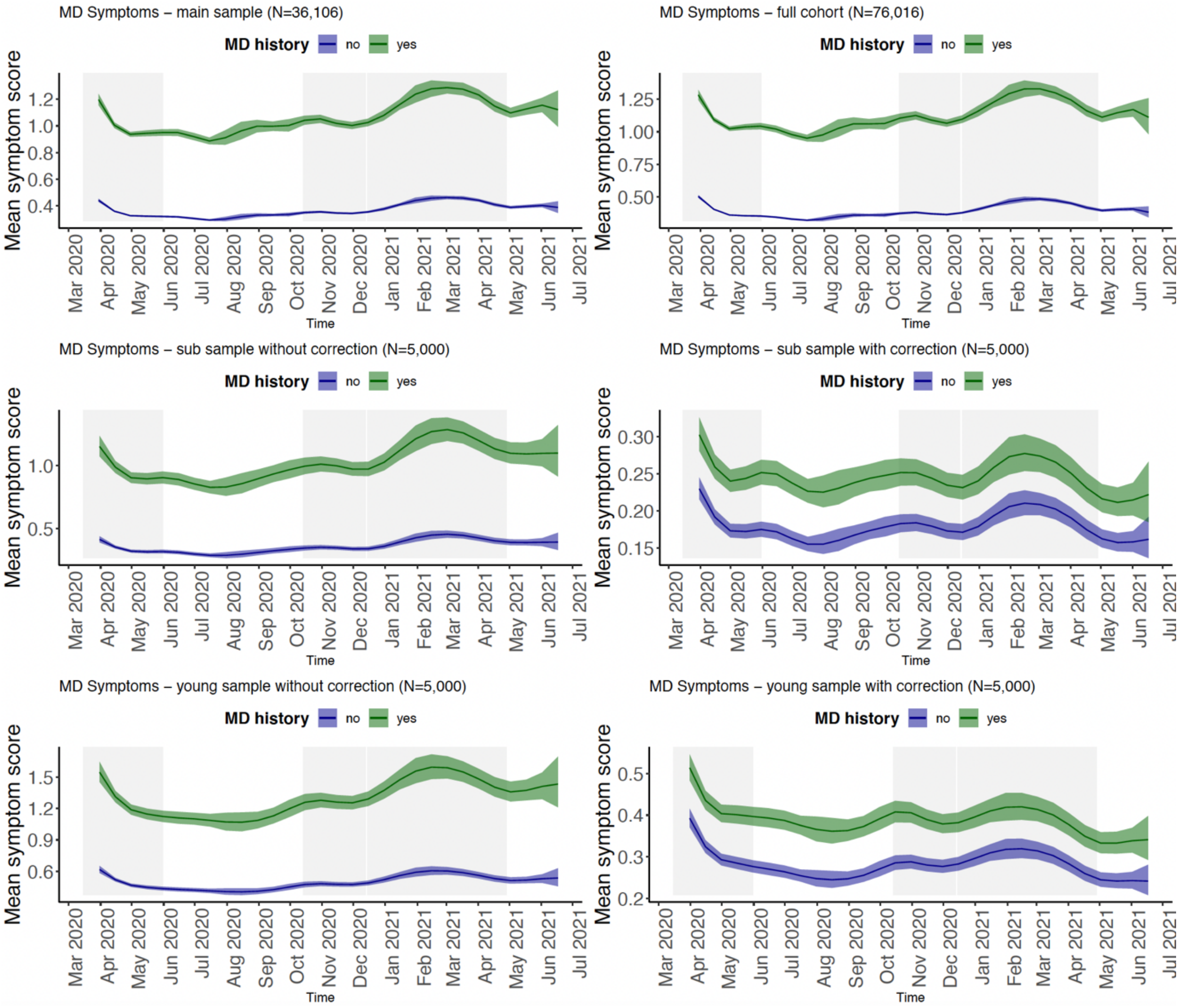

**Figure S27. The longitudinal trajectory of MDD during the COVID-19 pandemic by lifetime history of MDD in the Lifelines cohort.** Shown are the fitted smooths with standard errors of MDD prevalence over time for study participants with and without MDD lifetime history. Results of GAM analyses are shown for the following analyses: 1) main analysis sample of N=36,106 participants (left upper panel), 2) full cohort of N=76,016 participants (right upper panel), 3) a sub sample of 5,000 participants selected from analysis (1) without correction for random effects (left mid panel), 4) the same sub sample of 5,000 participants as (3) with correction for random effects (right mid panel), 5) a sub sample of the youngest 5,000 participants without correction for random effects (left lower panel), and 6) a sub sample of the youngest 5,000 participants with correction for random effects (right lower panel). The y-axis shows the prevalence in % and the x-axis the time in months. For the mixed-effect models, the random effects were removed before plotting and thus yielded lower prevalence estimates. The grey rectangles denoted the three lockdowns in the Netherlands.

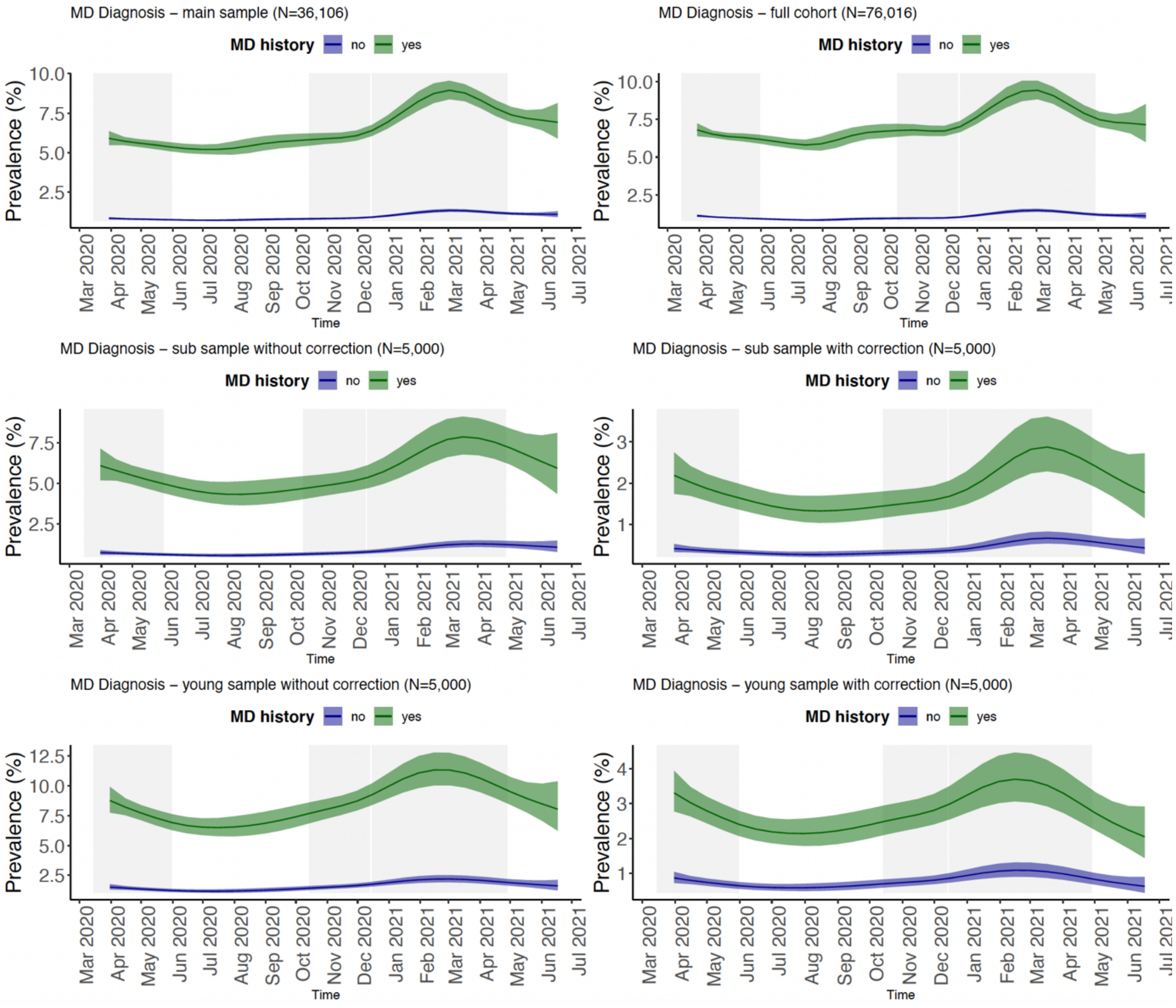

**Figure S28. The longitudinal trajectory of anxiety symptom score during the COVID-19 pandemic by lifetime history of GAD in the Lifelines cohort.** Shown are the fitted smooths with standard errors of mean anxiety symptom score over time for study participants with and without GAD lifetime history. Results of GAM analyses are shown for the following analyses: 1) main analysis sample of N=36,106 participants (left upper panel), 2) full cohort of N=76,016 participants (right upper panel), 3) a sub sample of 5,000 participants selected from analysis (1) <u>without correction</u> for random effects (left mid panel), 4) the same sub sample of 5,000 participants as (3) <u>with correction</u> for random effects (right mid panel), 5) a sub sample of the youngest 5,000 participants <u>without correction</u> for random effects (left lower panel), and 6) a sub sample of the youngest 5,000 participants <u>with correction</u> for random effects (right lower panel). The y-axis shows the mean anxiety symptom sum score and the x-axis the time in months. For the mixed-effect models, the random effects were removed before plotting and thus yielded lower prevalence estimates. The grey rectangles denoted the three lockdowns in the Netherlands.

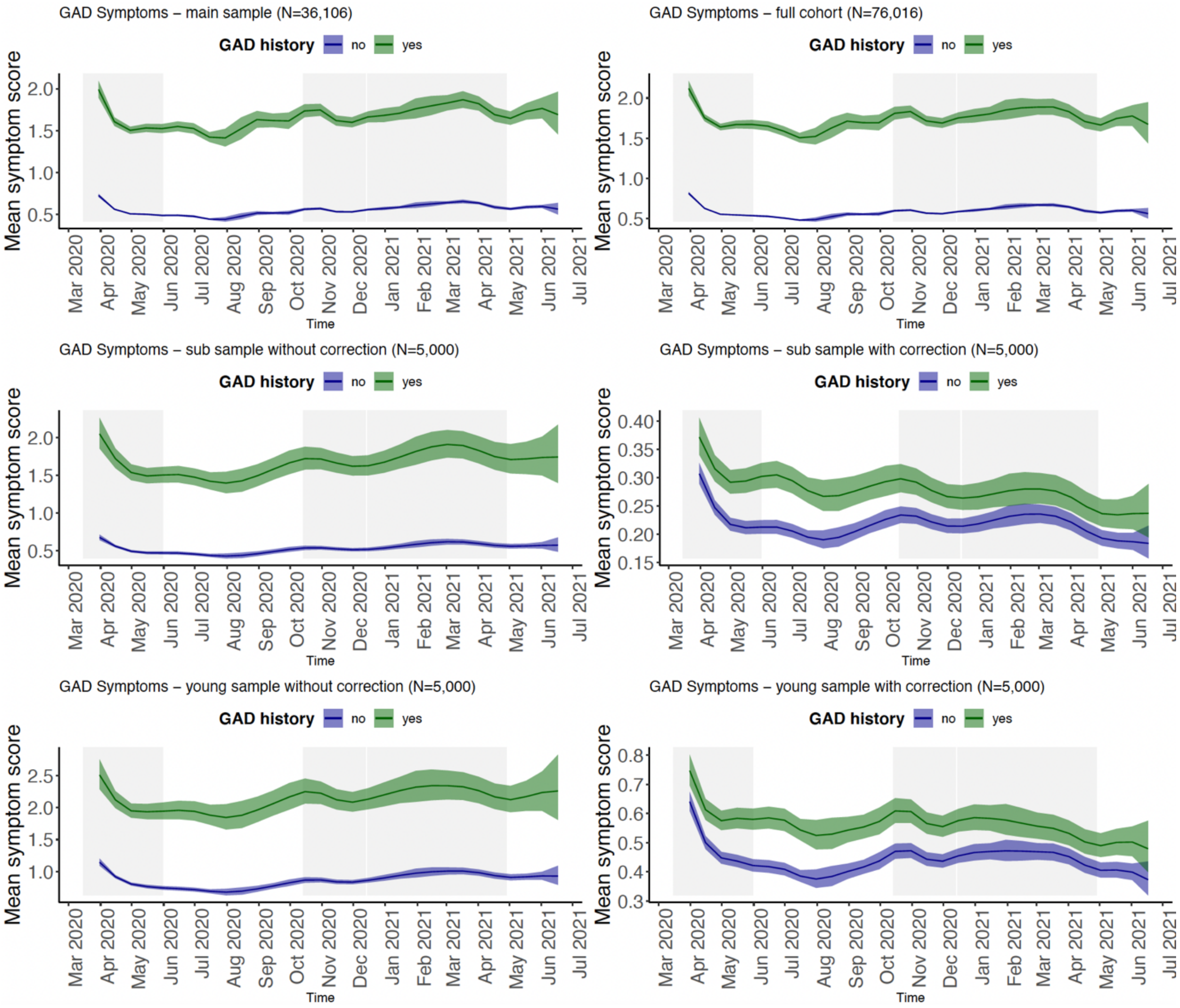

**Figure S29. The longitudinal trajectory of GAD during the COVID-19 pandemic by lifetime history of GAD in the Lifelines cohort.** Shown are the fitted smooths with standard errors of GAD prevalence over time for study participants with and without GAD lifetime history. Results of GAM analyses are shown for the following analyses: 1) main analysis sample of N=36,106 participants (left upper panel), 2) full cohort of N=76,016 participants (right upper panel), 3) a sub sample of 5,000 participants selected from analysis (1) without correction for random effects (left mid panel), 4) the same sub sample of 5,000 participants as (3) with correction for random effects (right mid panel), 5) a sub sample of the youngest 5,000 participants without correction for random effects (left lower panel), and 6) a sub sample of the youngest 5,000 participants with correction for random effects (right lower panel). The y-axis shows the prevalence in % and the x-axis the time in months. For the mixed-effect models, the random effects were removed before plotting and thus yielded lower prevalence estimates. The grey rectangles denoted the three lockdowns in the Netherlands.

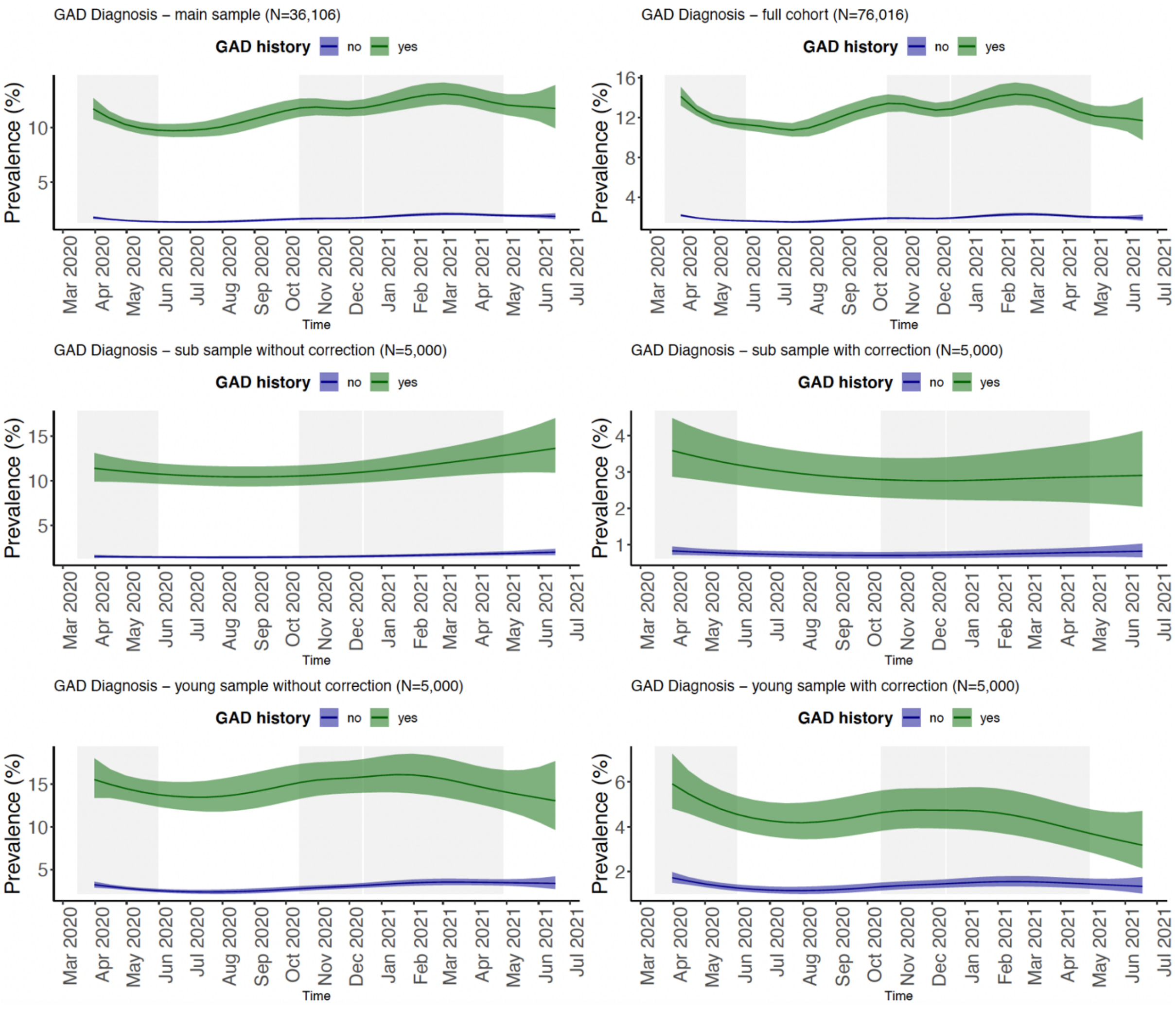

**Figure S30. The longitudinal trajectory of suicidal ideations during the COVID-19 pandemic by lifetime history of MDD in the Lifelines cohort.** Shown are the fitted smooths with standard errors of suicidal ideation prevalence over time for study participants with and without MDD lifetime history. Results of GAM analyses are shown for the following analyses: 1) main analysis sample of N=36,106 participants (left upper panel), 2) full cohort of N=76,016 participants (right upper panel), 3) a sub sample of 5,000 participants selected from analysis (1) without correction for random effects (left mid panel), 4) the same sub sample of 5,000 participants as (3) with correction for random effects (right mid panel), 5) a sub sample of the youngest 5,000 participants without correction for random effects (left lower panel), and 6) a sub sample of the youngest 5,000 participants with correction for random effects (right lower panel). The y-axis shows the prevalence in % and the x-axis the time in months. For the mixed-effect models, the random effects were removed before plotting and thus yielded lower prevalence estimates. The grey rectangles denoted the three lockdowns in the Netherlands.

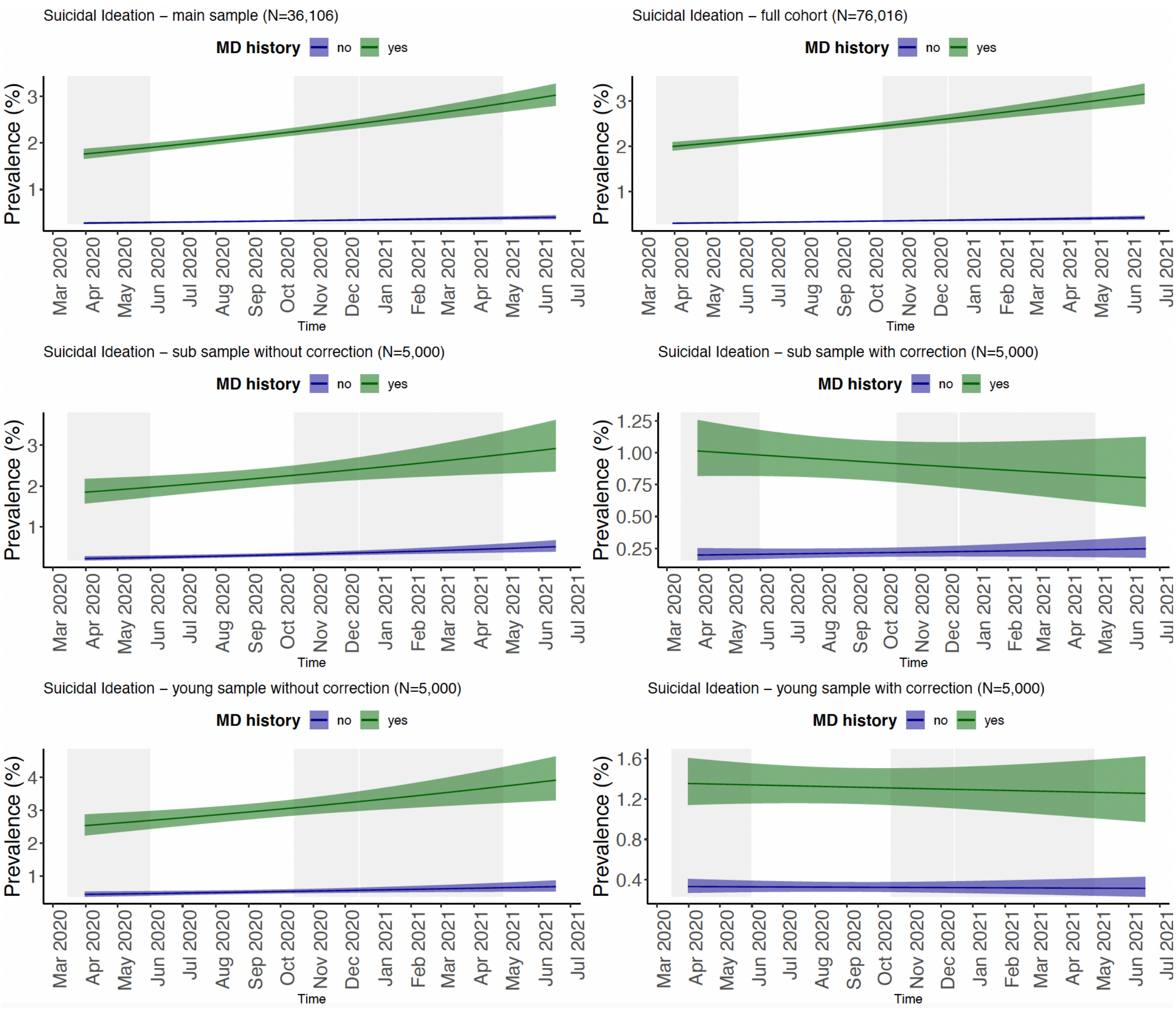

#### 7. Figure S31-35 - longitudinal trajectories of phenotypic outcomes by lifetime history of MD/GAD visualized using difference plots

**Figure S31. The difference in trajectory of depressive symptoms between study participants with and without a history of MDD during the COVID-19 pandemic in the Lifelines cohort.** Shown are the difference in fitted smooths of mean depressive symptom score between study participants with and without MDD lifetime history. Results of GAM analyses are shown for the following analyses: 1) main analysis sample of N=36,106 participants (left upper panel), 2) full cohort of N=76,016 participants (right upper panel), 3) a sub sample of 5,000 participants selected from analysis (1) <u>without correction</u> for random effects (left mid panel), 4) the same sub sample of 5,000 participants as (3) <u>with correction</u> for random effects (right mid panel), 5) a sub sample of the youngest 5,000 participants <u>without correction</u> for random effects (left lower panel), and 6) a sub sample of the youngest 5,000 participants <u>with correction</u> for random effects (right lower panel). The y-axis shows the mean difference in symptom sum score and the x-axis the time in months. The grey rectangles denoted the three lockdowns in the Netherlands.

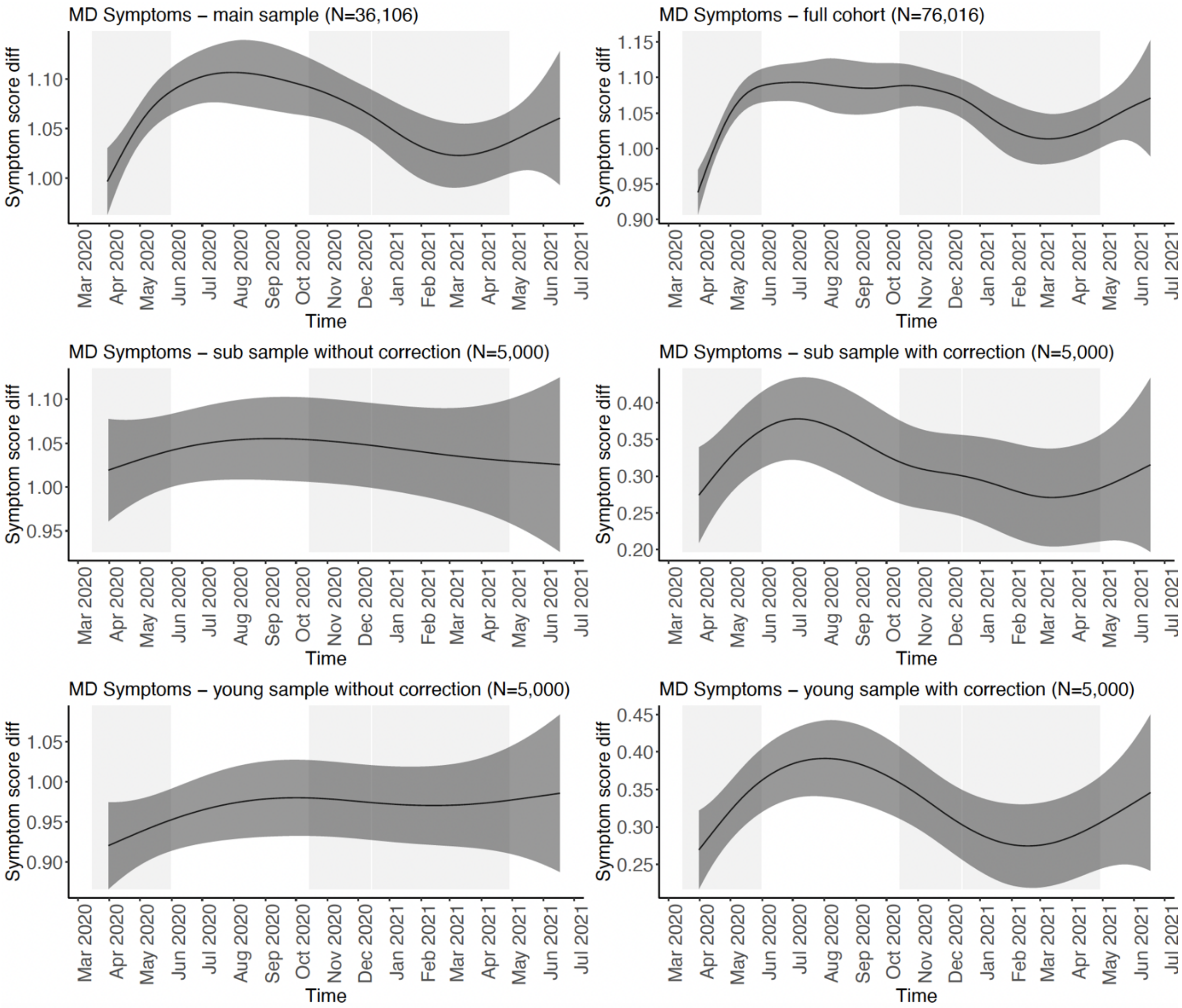

**Figure S32. The difference in trajectory of MDD between study participants with and without a history of MDD during the COVID-19 pandemic in the Lifelines cohort.** Shown are the difference in fitted smooths of the prevalence between study participants with and without MDD lifetime history. Results of GAM analyses are shown for the following analyses: 1) main analysis sample of N=36,106 participants (left upper panel), 2) full cohort of N=76,016 participants (right upper panel), 3) a sub sample of 5,000 participants selected from analysis (1) without correction for random effects (left mid panel), 4) the same sub sample of 5,000 participants as (3) with correction for random effects (right mid panel), 5) a sub sample of the youngest 5,000 participants without correction for random effects (left lower panel), and 6) a sub sample of the youngest 5,000 participants with correction for random effects (right lower panel). The y-axis shows the difference in prevalence in % and the x-axis the time in months. The grey rectangles denoted the three lockdowns in the Netherlands.

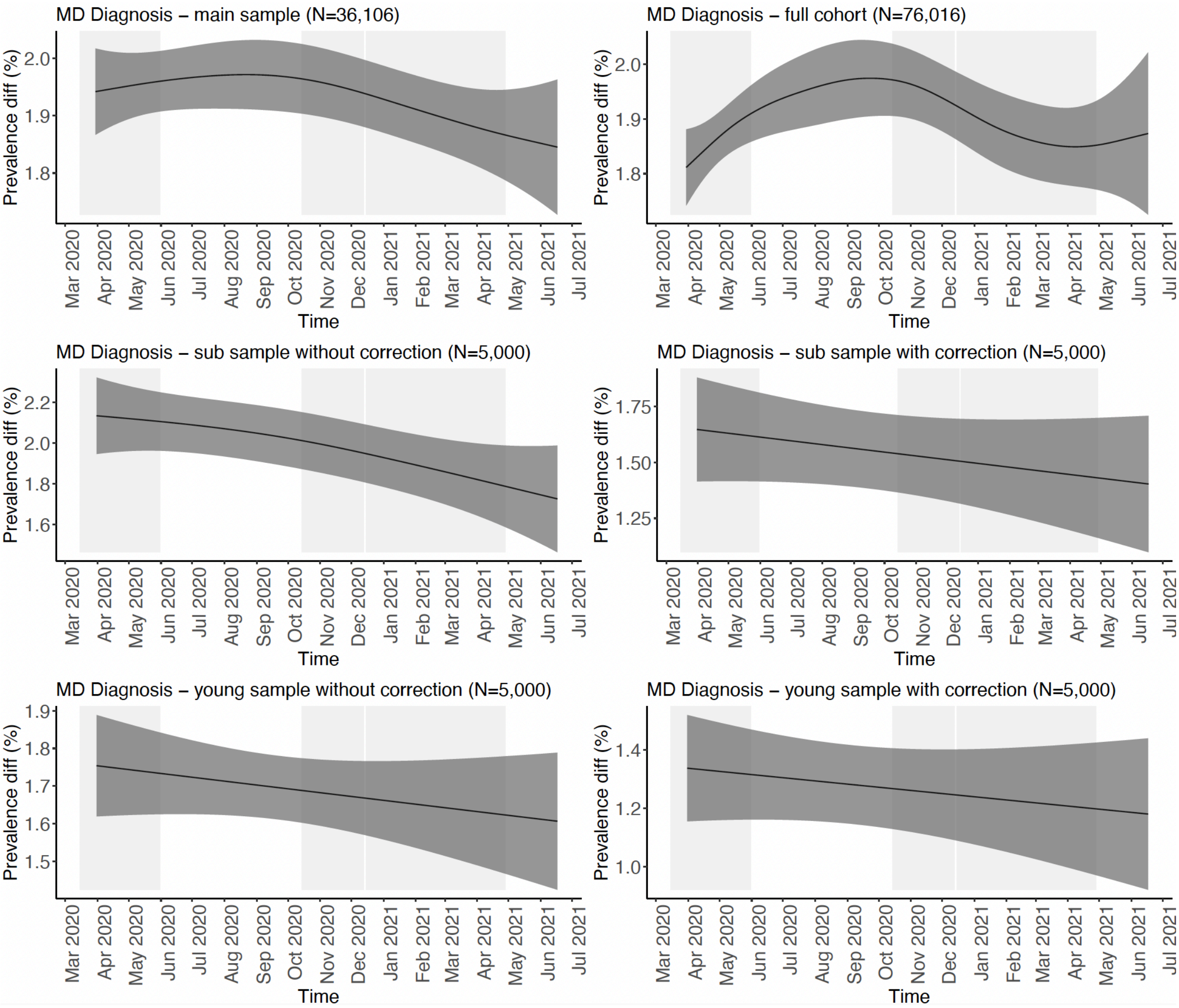

**Figure S33. The difference in trajectory of anxiety symptoms between study participants with and without a history of GAD during the COVID-19 pandemic in the Lifelines cohort.** Shown are the difference in fitted smooths of mean anxiety symptom score between study participants with and without GAD lifetime history. Results of GAM analyses are shown for the following analyses: 1) main analysis sample of N=36,106 participants (left upper panel), 2) full cohort of N=76,016 participants (right upper panel), 3) a sub sample of 5,000 participants selected from analysis (1) without correction for random effects (left mid panel), 4) the same sub sample of 5,000 participants as (3) with correction for random effects (right mid panel), 5) a sub sample of the youngest 5,000 participants without correction for random effects (left lower panel), and 6) a sub sample of the youngest 5,000 participants with correction for random effects (right lower panel). The y-axis shows the difference in mean anxiety symptom sum score and the x-axis the time in months. The grey rectangles denoted the three lockdowns in the Netherlands.

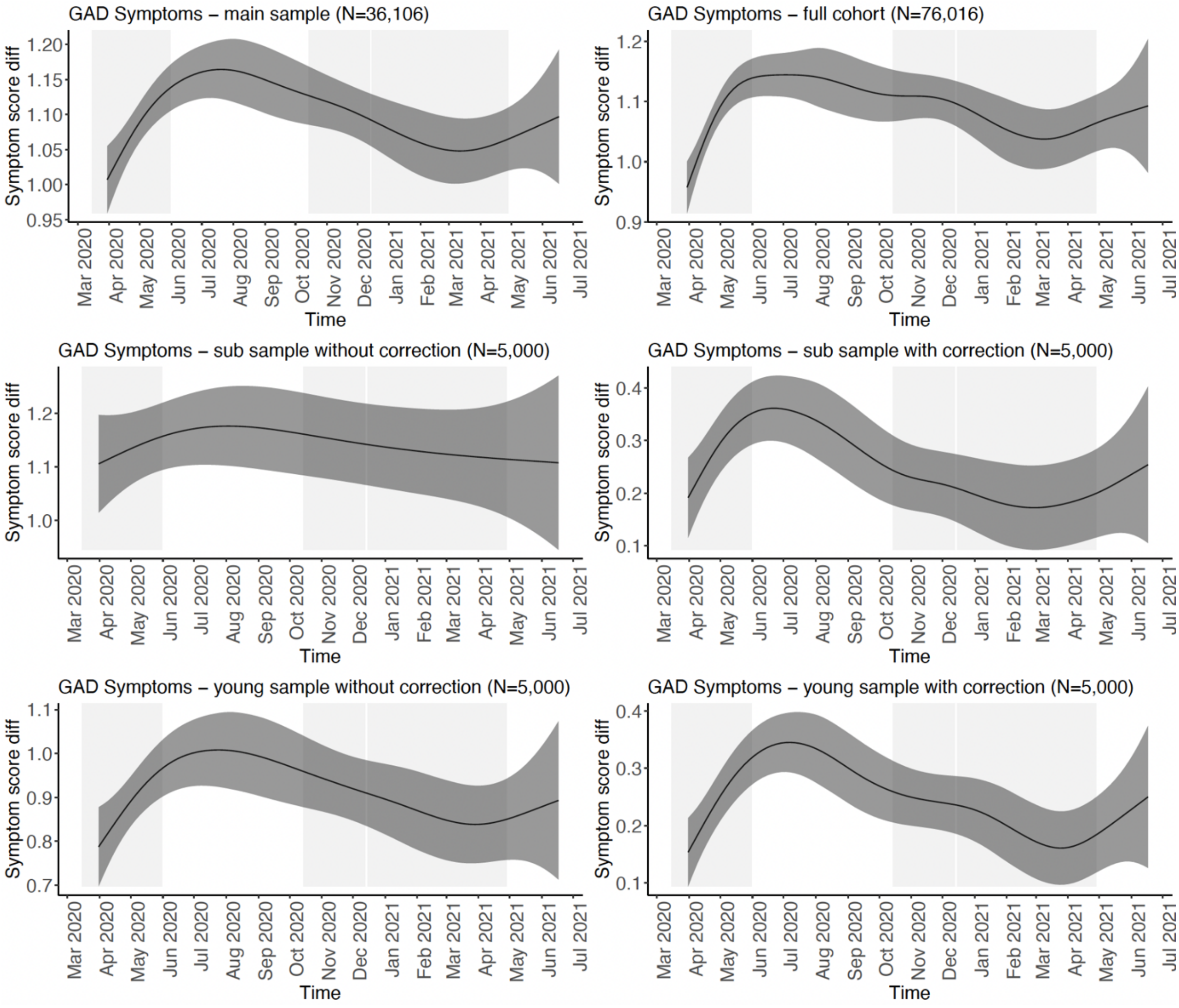

**Figure S34. The difference in trajectory of GAD between study participants with and without a history of GAD during the COVID-19 pandemic in the Lifelines cohort.** Shown are the difference in fitted smooths of the prevalence between study participants with and without GAD lifetime history. Results of GAM analyses are shown for the following analyses: 1) main analysis sample of N=36,106 participants (left upper panel), 2) full cohort of N=76,016 participants (right upper panel), 3) a sub sample of 5,000 participants selected from analysis (1) without correction for random effects (left mid panel), 4) the same sub sample of 5,000 participants as (3) with correction for random effects (right mid panel), 5) a sub sample of the youngest 5,000 participants without correction for random effects (left lower panel), and 6) a sub sample of the youngest 5,000 participants with correction for random effects (right lower panel). The y-axis shows the difference in prevalence in % and the x-axis the time in months. The grey rectangles denoted the three lockdowns in the Netherlands.

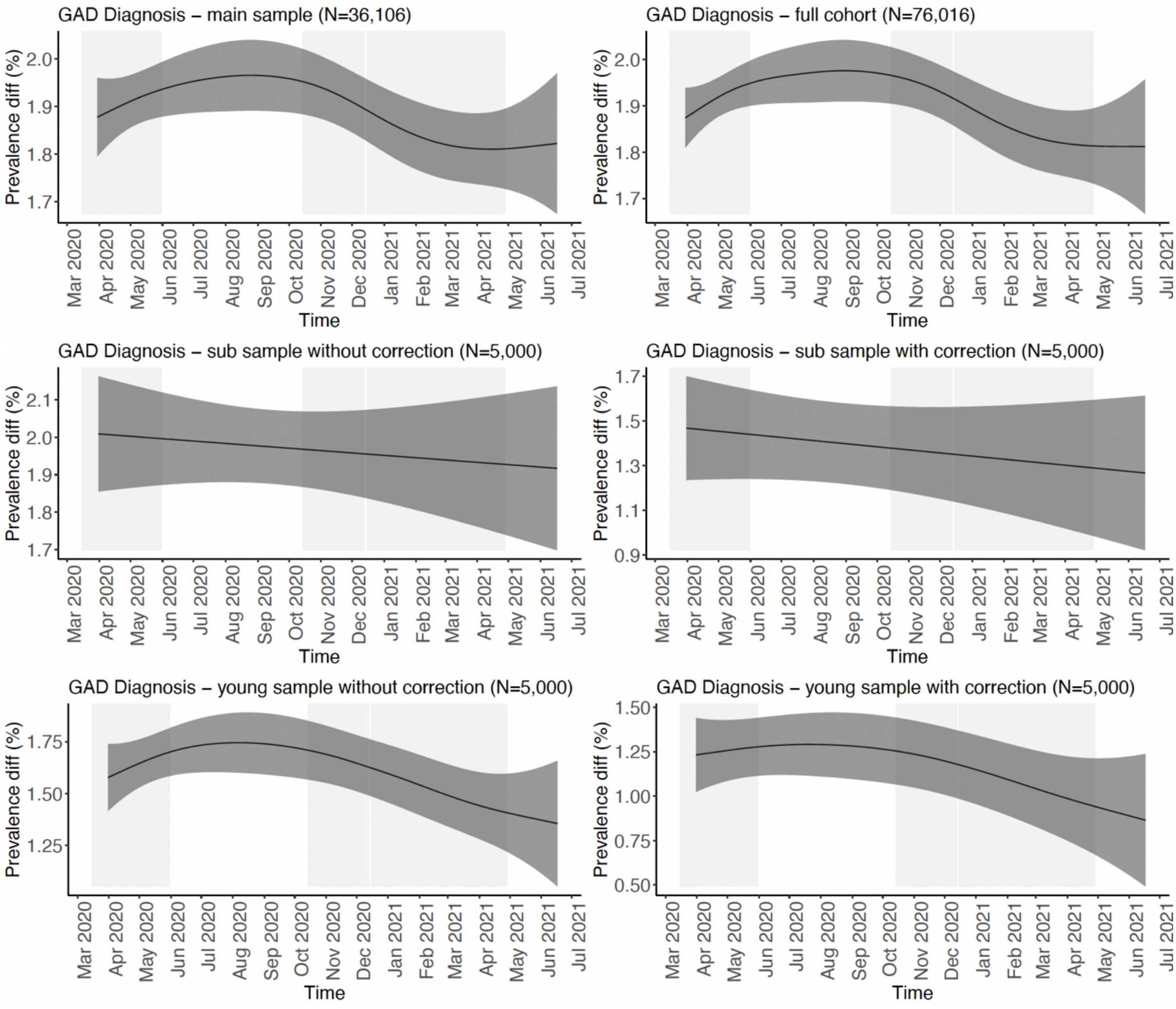

**Figure S35. The difference in trajectory of suicidal ideations between study participants with and without a history of MDD during the COVID-19 pandemic in the Lifelines cohort.** Shown are the difference in fitted smooths of the prevalence between study participants with and without MDD lifetime history. Results of GAM analyses are shown for the following analyses: 1) main analysis sample of N=36,106 participants (left upper panel), 2) full cohort of N=76,016 participants (right upper panel), 3) a sub sample of 5,000 participants selected from analysis (1) without correction for random effects (left mid panel), 4) the same sub sample of 5,000 participants as (3) with correction for random effects (right mid panel), 5) a sub sample of the youngest 5,000 participants without correction for random effects (left lower panel), and 6) a sub sample of the youngest 5,000 participants with correction for random effects (right lower panel). The y-axis shows the difference in prevalence in % and the x-axis the time in months. The grey rectangles denoted the three lockdowns in the Netherlands.

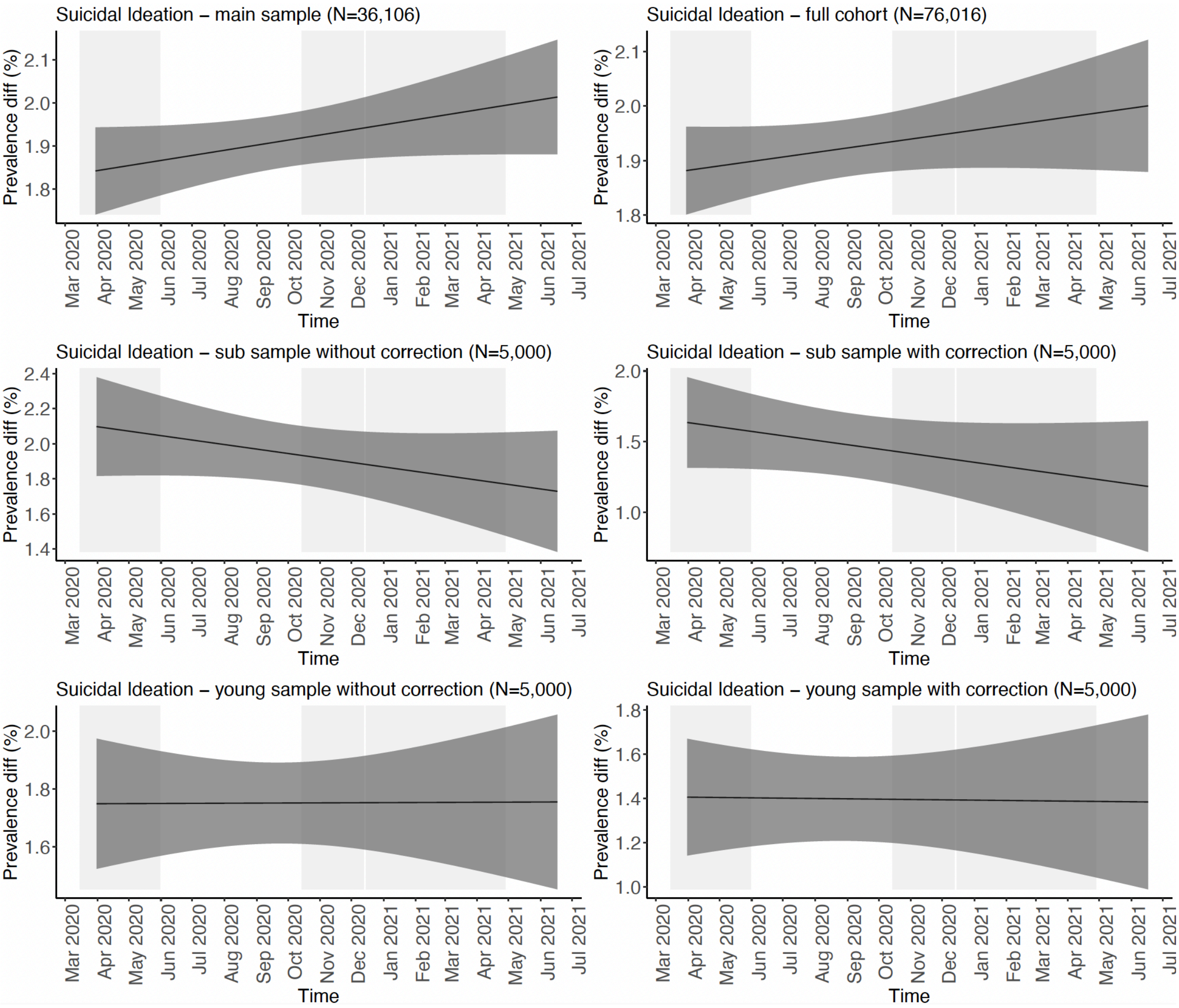
